# Hands are frequently contaminated with fecal bacteria and enteric pathogens globally: A systematic review and meta-analysis

**DOI:** 10.1101/2022.07.11.22277510

**Authors:** Molly E. Cantrell, Émile Sylvestre, Hannah Wharton, Rahel Scheidegger, Lou Curchod, David M. Gute, Jeffrey Griffiths, Timothy R Julian, Amy J. Pickering

## Abstract

Enteric pathogen infections are a leading cause of morbidity and mortality globally, with the highest disease burden in low-income countries. Hands act as intermediaries in enteric pathogen transmission, transferring enteric pathogens between people and the environment through contact with fomites, food, water, and soil. In this study, we conducted a systematic review of prevalence and concentrations of fecal indicator microorganisms (i.e., *E. coli*, fecal coliform) and enteric pathogens on hands. We identified eighty-four studies, reporting 35,440 observations of hand contamination of people in community or household settings. The studies investigated 44 unique microorganisms, of which the most commonly reported indicators were *E. coli* and fecal coliforms. Hand contamination with 12 unique enteric pathogens was reported, with adenovirus and norovirus as the most frequent. Mean *E. coli* prevalence on hands was 62% [95% CI 40%-82%] and mean fecal coliform prevalence was 66% [95% CI 22%-100%]. Hands were more likely to be contaminated with *E. coli* in low/lower-middle-income countries (prevalence: 69% [95% CI 48% - 88%]) than in upper-middle/high-income countries (6% [95% CI 2% - 12%]). The review also highlighted the importance of standardizing hand sampling methods, as hand rinsing was associated with greater fecal contamination compared to other sampling methods.

## Introduction

Hands are an important route of enteric pathogen transmission. The routes enteric pathogens can take from contaminated feces to a susceptible host are complex and include fomites, food, drinking water, and soil (see the fecal-diagram, Figure S1). Hands can facilitate direct exposure to pathogens (hand-to-mouth contacts), as well as indirect exposure through contact with drinking water, food, and fomites (1).

Several studies have implicated contaminated hands in child diarrheal disease risk (2,3). A study of handwashing interventions at day care centers in the United States showed significant reductions in child diarrheal disease (4). Similarly, in rural Bangladesh, a study of households measured fecal contamination in hands, soil, water, flies, and food, and found that hands were the most strongly associated with increased subsequent risk of diarrheal illness among children under five years old (3). In an exposure study in India, infants mouthing their own hands posed the second highest daily risk of enteric infection after soil ingestion (5). A fecal exposure assessment model showed that children in Tanzania ingest a significantly greater amount of feces each day from hand-to-mouth contacts (0.93 mg) than from drinking water (0.098 mg) (2). Another Tanzanian study found that viral pathogens were more frequently found on hands than in drinking water (6). Further evidence on the role of contaminated hands in diarrheal disease risks includes evidence of the efficacy of hand hygiene interventions on reducing diarrheal disease risks in communities globally (4,7–9). The goal of this study is to determine the extent to which hands are contaminated with enteric pathogens and other fecal indicators. We further aimed to identify factors that influence the prevalence and concentrations of fecal and enteric pathogen contamination on hands, including age, gender, country income, urbanicity, climate, and hand sampling method.

## Materials and Methods

We conducted a systematic review of fecal indicator microorganisms and enteric pathogen detection on hands following the PRISMA-P guidelines (see Supplemental Information for PRISMA-P Checklist and search terms). Studies were identified in an electronic search of PubMed, Embase, and Web of Science databases, first in March 2018, updated in June 2020, and updated again in September 2022. The following search terms were used: (((fecal OR pathogenic OR enteric) AND bacteria) OR e. coli OR enterococci OR helminth OR protozoa OR virus OR phage) AND hand AND contamination. Two independent reviewers completed initial title and abstract screening and a third reviewer resolved discrepancies. Data extraction from each study was completed by one reviewer and checked by a second; data extraction included type of organism, prevalence (defined as percentage of hand samples positive for the organism), the concentration of the organism measured on hands, methods used, and detection limit. Given wide variation in the units used to report hand contamination, mean concentration values were synthesized and reported using units of log_10_CFU/hand (see Supplemental Information).

We included peer-reviewed published studies of all designs that measured fecal indicators or enteric pathogens on human hands for people in non-occupational settings, such as in households and communities. Studies were excluded for the following reasons: microorganisms measured were not enteric pathogens or fecal indicators; hands were artificially contaminated with bacteria; studies did not present primary data; studies were conducted in occupational settings such as food handling, farm, clinical, or laboratory settings; and/or dealt with food and animal contamination.

We conducted a meta-analysis to analyze the subset of studies with estimates of the prevalence rates (defined as the proportion of hands with detectable contamination) for *E. coli* and fecal coliforms. *E. coli* and fecal coliforms were chosen as the fecal indicators to use for a meta-analysis because they were reported by the largest number of studies. Within the meta-analysis, we combined the prevalence rates of hand contamination using the inverse variance method to obtain a weighted average across individual studies. We modelled the mean proportion 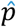 (the number of positive samples *n*_*s*_ divided by the total number of samples *n*), as a Bernoulli trial process. For the *i*^th^ study, the variance of the estimator of 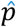 can be approximated with a normal distribution as follows:

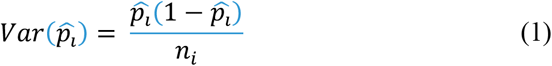

Using the inverse variance method, the weight for the *i*th study is then 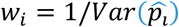. We used a double arcsine function to transform the prevalence 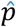 to a value that is not constrained to the 0-1 range and back-transformed to 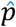 after pooling (10). To calculate pooled sample prevalences, we chose an inverse variance heterogeneity (IVhet) model rather than the more conventional random-effects model. The advantage of the IVhet model is that it can maintain the individual study weights despite substantial between-study heterogeneity (11). We evaluated publication bias with Luis Furuya-Kanamori (LFK) index and Doi plots (12). We performed analyses with the MetaXL add-in version 5.3 in Microsoft Excel and the *metafor* package in R (13).

We compared *E. coli* and fecal coliform prevalence between i) children and adults, ii) country income groups (low and lower-middle vs. upper-middle, and high-income), iii) urban and rural areas, iv) climate classifications (Köppen-Geiger), and v) types of hand sampling methods. We tested for differences between the mean pooled prevalence of two groups using a two-sample z-test allowing unequal variance. Country income levels were based on World Bank country income classifications (available via the *World Bank Data Help Desk*, https://datahelpdesk.worldbank.org/) for the year the study was conducted; if that information was not reported, then the year the study was published was used (see Supplemental Information for how fiscal year 2023 classifications are defined). The climate classifications were based on Köppen-Geiger Climate Classifications where zone A is tropical or equatorial, zone B is arid or dry, zone C is warm/mild temperate, zone D is continental, and zone E is polar (14).

## Results and Discussion

### Fecal indicator bacteria and enteric pathogens

We identified 84 studies, which reported forty-four unique fecal indicator bacteria and enteric pathogens found on hands (n=35,440 observations). The paper search selection process, including records identified and excluded is reported in Figure S2. Of the 44 unique microorganisms reported, 12 were pathogens (inclusive of enteric and opportunistic pathogens) and the other 32 were fecal indicator bacteria. The most common indicators were *E. coli* (56 studies, or 67%), fecal coliforms (24 studies, or 29%), and enterococci (9 studies, or 11%). The most commonly measured pathogens were adenovirus (5 studies) and norovirus (5 studies). All indicators reported are summarized in Table 1. We note that not including specific names of enteric pathogens in our search terms may have limited the number of papers identified reporting enteric pathogens.

**Table 1:** Pathogens and indicators measured on hands as reported in the included studies

| Type of Micro-organisms | Microorganism | Total Number of Studies | Prevalence |  |  |  | Concentration |  |  |  | Countries, References |
| --- | --- | --- | --- | --- | --- | --- | --- | --- | --- | --- | --- |
|  |  |  | Number of Studies | Number of Samples | Percent Positive (% average)<sup>1</sup> | Range (%) | Number of Studies | Number of Samples | Mean Concentration (log<sub>10</sub>CFU/hand, simple weighted average by sample size) | Range |  |
| Bacteria | Aerobic Plate Count<sup>2</sup> | 2 | 2 | 99 | 92 | 84 – 100 | 1 | 74 | 4.38 | - | Ireland(15)<br>U.S.(16) |
|  | *Bacillus* spp | 2 | 2 | 370 | 5.2 | 3 – 7.5 | 0 | - | - | - | Mauritius(17)<br>Mauritius(18) |
|  | *Bacteroidales* Cow<sup>2</sup> | 2 | 2 | 1185 | 93.3 | 89.6 – 97.5 | 1 | 914 | 3.80 log<sub>10</sub> gene copies / hand | 3.67 – 3.84 log<sub>10</sub> gene copies / hand | Bangladesh(19)<br>India(20) |
|  | *Bacteroidales* General | 5 | 5 | 1333 | 78.9 | 50 – 98.8 | 2 | 935 | 2.53 log<sub>10</sub> gene copies / hand | 2.44-2.8 log<sub>10</sub> gene copies / hand | Bangladesh(21)<br>India(22)<br>Tanzania(23)<br>Tanzania(24)<br>India(20) |
|  | *Bacteroidales* Human<sup>2</sup> | 5 | 5 | 2039 | 21 | 3 – 32.2 | 1 | 271 | 1.93 log<sub>10</sub> gene copies / hand | 1.91-1.94 log<sub>10</sub> gene copies / hand | Bangladesh(19)<br>Tanzania(6)<br>India(22)<br>Tanzania(24)<br>India(20) |
|  | *Campylobacter jejuni* | 2 | 2 | 155 | 10.0 | 2.2 - 15 | 2 | -110 | 1.46 log<sub>10</sub> gene copies / hand | 1.17 – 1.94 log<sub>10</sub> gene copies / hand | Kenya(25)<br>Uganda(26) |

<sup>1</sup> For indicators with 2-9 studies, percent positive is calculated as the unweighted arithmetic mean of the percent positive values reported by the individual studies. For indicators with at least 10 studies (only *E. coli* and fecal coliforms), percent positive was calculated using the explained meta-analysis and the 95% confidence interval is reported.
<sup>2</sup> Commonly used fecal indicator bacteria

| Type of Micro-organisms | Microorganism | Total Number of Studies | Prevalence |  |  |  | Concentration |  |  |  | Countries, References |
| --- | --- | --- | --- | --- | --- | --- | --- | --- | --- | --- | --- |
|  |  |  | Number of Studies | Number of Samples | Percent Positive (% average)<sup>1</sup> | Range (%) | Number of Studies | Number of Samples | Mean Concentration (log<sub>10</sub>CFU/hand, simple weighted average by sample size) | Range |  |
|  | *Clostridium perfringens* | 1 | 0 | - | - | - | 1 | 900 | 1.54 | - | Bangladesh(27) |
|  | Coagulase negative *Staphylococcus spp.* | 3 | 3 | 770 | 70.2 | 45 – 95.5 | 0 | - | - | - | Mauritius(17)<br>Mauritius(18)<br>India(28) |
|  | *Commensal flora* | 1 | 1 | 133 | 31.5 | - | 0 | - | - | - | India(29) |
|  | Diarrheagenic *E. coli* (DEC)<sup>3</sup> | 3 | 3 | 1006 | 7.52 | 0.3 – 19.6 | 1 | 222 | 1.25 | - | Bangladesh(19)<br>Tanzania(6)<br>Tanzania(30) |
|  | *E. coli*<sup>2</sup> | 56 | 47 | 20288 | 62 [40, 82] | 0.5 – 100 | 35 | 17999 | 1.16 | -0.1 – 6.7 | Pakistan(31)<br>Kenya(25)<br>India(32)<br>India(33)<br>Bangladesh(21)<br>Bangladesh(34)<br>Croatia(35)<br>Tanzania(36)<br>Netherlands(37)<br>India,<br>Mozambique(38)<br>Great Britain(39)<br>India(40)<br>Bangladesh(41)<br>Zimbabwe(42)<br>Bangladesh(19)<br>Democratic Republic of Congo(43) |

<sup>3</sup> Diarrheagenic *E. coli* includes Enterohemorrhagic *E. coli* (EHEC, stx1, stx2, eaeA), Enteroinvasive *E. coli* (EIEC, ipaH), Enteropathogenic *E. coli* (EPEC), Enterotoxigenic *E. coli* (ETEC, stib/ltib), Shiga-toxin producing *E. coli* (STEC)

| Type of Micro-organisms | Microorganism | Total Number of Studies | Prevalence |  |  |  | Concentration |  |  |  | Countries, References |
| --- | --- | --- | --- | --- | --- | --- | --- | --- | --- | --- | --- |
|  |  |  | Number of Studies | Number of Samples | Percent Positive (% average)<sup>1</sup> | Range (%) | Number of Studies | Number of Samples | Mean Concentration (log<sub>10</sub>CFU/hand, simple weighted average by sample size) | Range |  |
|  |  |  |  |  |  |  |  |  |  |  | India(44)<br>Peru(45)<br>Ethiopia(46)<br>Kenya(47)<br>Malawi(48)<br>Bangladesh(49)<br>Tanzania(50)<br>Peru(51)<br>Peru(52)<br>Bangladesh(27)<br>Great Britain(53)<br>Tanzania(54)<br>U. S.(55)<br>Peru(56)<br>India(29)<br>Kenya(57)<br>India(58)<br>Indonesia(59)<br>U. S.(16)<br>Tanzania(60)<br>Mexico(61)<br>Tanzania(30)<br>Tanzania(62)<br>Bangladesh(63)<br>Zimbabwe(64)<br>Zimbabwe(65)<br>Indonesia(66)<br>Mauritius(17)<br>Tanzania(67)<br>Tanzania(23)<br>Tanzania(24)<br>Bangladesh(68)<br>Thailand(69)<br>Bangladesh(70)<br>India(71)<br>Peru(72) |
|  |  |  | Number of Studies | Number of Samples | Percent Positive (% average)<sup>1</sup> | Range (%) | Number of Studies | Number of Samples | Mean Concentration (log<sub>10</sub>CFU/hand, simple weighted average by sample size) | Range |  |
|  |  |  |  |  |  |  |  |  |  |  | Kenya(73)<br>Mauritius(18)<br>India(28)<br>Zambia(74) |
|  | Enteroaggregative *E. coli* (EAEC)<sup>4</sup> | 4 | 4 | 1278 | 11.3 | 2.0 – 19.6 | 2 | 272 | 1.34 | 1.25 – 1.73 | India(33)<br>Bangladesh(19)<br>Tanzania(6)<br>Tanzania(30) |
|  | *Enterobacter* spp. | 2 | 2 | 804 | 0.48 | 0.2 – 0.75 | 0 | - | - | - | Great Britain(53)<br>India(28) |
|  | *Enterobacteriaceae* | 1 | 1 | 80 | 37.5 | 20 – 45 | 1 | 30 | 2.69 | 2.28 – 3.08 | Netherlands(37) |
|  | *Enterococci*<sup>2</sup><br>(includes *Enterococcus* spp) | 9 | 8 | 2114 | 60.1 | 2 - 100 | 6 | 585 | 1.73 | 1.00 – 2.60 | India,<br>Mozambique(38)<br>Great Britain(39)<br>Bangladesh(49)<br>Great Britain(53)<br>Tanzania(54)<br>Tanzania(30)<br>Tanzania(62)<br>Tanzania(24)<br>India(28) |
|  | *Enterococcus faecalis*<sup>2</sup> | 1 | 1 | 133 | 11 | - | 0 | - | - | - | India(29) |
|  | Fecal bacteria<sup>2</sup><br>(defined as total of | 1 | 1 | 72 | 31 | - | 1 | 72 | 2.22 | 1.72 – 2.58 | U. S.(75) |

<sup>4</sup> Three studies reported AggR

| Type of Micro-organisms | Microorganism | Total Number of Studies | Prevalence |  |  |  | Concentration |  |  |  | Countries, References |
| --- | --- | --- | --- | --- | --- | --- | --- | --- | --- | --- | --- |
|  |  |  | Number of Studies | Number of Samples | Percent Positive (% average)<sup>1</sup> | Range (%) | Number of Studies | Number of Samples | Mean Concentration (log<sub>10</sub>CFU/hand, simple weighted average by sample size) | Range |  |
|  | fecal coliforms and enterococci) |  |  |  |  |  |  |  |  |  |  |
|  | Fecal coliform<sup>2</sup> | 24 | 20 | 11794 | 66<br>[22, 100] | 6 – 100 | 11 | 8162 | 1.25 | 0.12 – 8.29 | Bangladesh(21)<br>Ethiopia(76)<br>Canada(77)<br>Canada(78)<br>U. S.(79)<br>Bangladesh(34)<br>Great Britain(39)<br>U. S.(80)<br>Zimbabwe(42)<br>U.S.(81)<br>Bangladesh(82)<br>Bangladesh(83)<br>U. S.(55)<br>Botswana(84)<br>U.S.(85)<br>Pakistan(86)<br>Mexico(61)<br>India(22)<br>U. S.(87)<br>Peru(70)<br>India(71)<br>India(20)<br>U. S.(88)<br>Botswana(89) |
|  | *Fecal Streptococci*<sup>2</sup> | 6 | 4 | 4445 | 52.9 | 7 – 98 | 5 | 5454 | 1.85 | 0.16 – 4.40 | Bangladesh(83)<br>Botswana(90)<br>Greece(91)<br>Tanzania(23)<br>Tanzania(67)<br>Thailand(69) |
|  | *Klebsiella* spp. | 6 | 6 | 1427 | 11.2 | 0.3 – 33.3 | 1 | 3 | 4.65 | 4.37 – 4.93 | Great Britain(39)<br>Great Britain(53) |
|  |  |  | Number of Studies | Number of Samples | Percent Positive (% average)<sup>1</sup> | Range (%) | Number of Studies | Number of Samples | Mean Concentration (log<sub>10</sub>CFU/hand, simple weighted average by sample size) | Range |  |
|  |  |  |  |  |  |  |  |  |  |  | India(29)<br>Tanzania(61)<br>Mauritius(18)<br>India(28) |
|  | Methicillin resistant staphylococcus aureus | 1 | 1 |  | 206 | 0.5 | 0 | - | - | - | U. S.(92) |
|  | Micrococcus spp. | 3 | 3 | 576 | 22.2 | 3.0 – 42.7 | 0 | - | - | - | U. S.(92)<br>Mauritius(17)<br>Mauritius(18) |
|  | Pathogenic *E. coli* | 4 | 4 | 1258 | 34.6 | 22.1 – 44.1 | 1 | 222 | 1.25 | - | Bangladesh(19)<br>Tanzania(6)<br>Tanzania(30)<br>Tanzania(24) |
|  | *Proteus* spp. | 1 | 1 | 200 | 1.5 | - | 0 | - | - | - | Mauritius(17) |
|  | *Pseudomonas* spp. | 3 | 3 | 806 | 3.7 | 1.5 – 5.5 | 0 | - | - | - | U. S.(92)<br>Mauritius(17)<br>India(28) |
|  | *Salmonella* spp. | 1 | 1 | 12 | 41.6 | 33.3 – 50 | 0 | - | - | - | Mexico(61) |
|  | *Serratia* spp. | 1 | 1 | 12 | 83.3 | - | 0 | - | - | - | Mexico(61) |
|  | *Shigella* spp. | 1 | 1 | 12 | 0 | - | 0 | - | - | - | Mexico(61) |
|  | *Shigella* spp. / EIEC | 2 | 2 | 155 | 10.8 | 2.2 – 19 | 2 | 155 | 3.44 log<sub>10</sub> gene copies / hand | -2.35 – 4.56 log<sub>10</sub> gene | Kenya(25)<br>Uganda(26) |
|  |  |  | Number of Studies | Number of Samples | Percent Positive (% average)<sup>1</sup> | Range (%) | Number of Studies | Number of Samples | Mean Concentration (log<sub>10</sub>CFU/hand, simple weighted average by sample size) | Range |  |
|  |  |  |  |  |  |  |  |  |  | copies / hand |  |
|  | *Staphylococcus aureus* | 3 | 3 | 565 | 59.3 | 19 – 92 | 1 | 7 | 5.93 | 5.74 – 6.11 | Benin(93)<br>India(29)<br>Mexico(61) |
|  | *Streptococcus Faecalis*<sup>2</sup> | 1 | 1 | 18 | 50 | - | 0 | - | - | - | Croatia(35) |
|  | *Streptococcus* spp. | 3 | 3 | 388 | 15.8 | 0-37 | 1 | 6 | 4.9 | - | Mauritius(18)<br>Mexico(61)<br>U. S.(92) |
|  | Total coliform<sup>2</sup> | 8 | 5 | 1266 | 60.8 | 27.5 – 97.8 | 4 | 1096 | 1.96 | 1.24 – 2.45 | Peru(45)<br>Peru(52)<br>U. S.(94)<br>Bangladesh(27)<br>Peru(53)<br>U.S.(16)<br>Tanzania(60)<br>Peru(72) |
|  | *Vibrio cholerae* | 1 | 1 | 70 | 14 | 12 – 16 | 0 | - | - | - | Kenya(25) |
| Viruses | Adenovirus | 5 | 5 | 576 | 22.1 | 4.5 – 50 | 2 | 155 | 3.92 log<sub>10</sub> gene copies / hand | 2.62 – 5.33 log<sub>10</sub> gene copies / hand | Kenya(25)<br>Uganda(26)<br>Tanzania(6)<br>Tanzania(30)<br>Finland(95) |
|  | Enterovirus | 3 | 3 | 390 | 8.8 | 6.3 – 11.1 | 0 | - | - | - | Tanzania(6)<br>Tanzania(30)<br>Tanzania(24) |
|  | Hepatitis A virus | 1 | 1 | 12 | 17 | - | 0 | - | - | - | Mexico(61) |
|  |  |  | Number of Studies | Number of Samples | Percent Positive (% average)<sup>1</sup> | Range (%) | Number of Studies | Number of Samples | Mean Concentration (log<sub>10</sub>CFU/hand, simple weighted average by sample size) | Range |  |
|  | Norovirus<sup>5</sup> | 5 | 5 | 1285 | 3.1 | 0 – 10 | 1 (1) | 50 (88) | 1.73 (0.2 log<sub>10</sub> gene copies / hand) | - | India(33)<br>Bangladesh(19)<br>U. S.(16)<br>Tanzania(62)<br>Finland(95) |
|  | Rotavirus | 4 | 4 | 325 | 23.4 | 0 – 100 | 0 | - | - | - | U.S.(96)<br>Mexico(61)<br>Tanzania(6)<br>Tanzania(30) |
| Protozoa | *Giardia Lamblia* | 2 | 2 | 926 | 35.3 | 2.3 – 67 | 0 | - | - | - | Bangladesh(19)<br>Mexico(61) |
| Multicellular animal parasites | Ascaris lumbricoides | 1 | 1 | 336 | 10 | - | 0 | - | - | - | Tanzania(60) |
|  | Hookworm | 1 | 1 | 336 | 2 | - | - | - | - | - | Tanzania(60) |
|  | Soil-transmitted helminths | 2 | 2 | 466 | 8.7 | 0.87 - 16.6 | 2 | 466 | 1.68 eggs/hand | 1 – 2.35 eggs/hand | South Africa(97)<br>Kenya(98) |

<sup>5</sup> Norovirus includes GI and GII

### Prevalence of pathogen and fecal indicator contamination on hands

Of the 84 studies identified, 76 (90%) reported contamination prevalence rates for the target pathogen or indicator. The average prevalence rate across all 44 indicators was 37.9% (median: 29.5%), ranging from 0% (no detectable contamination) to 100% (detectable contamination on all study participants). *E. coli* and fecal coliform contamination on hands were common among the study populations. Overall, mean *E. coli* prevalence was 62% (range: 0.5 - 100%) and mean fecal coliform prevalence was 66% (range: 6 - 100%). Our findings confirm that hands are frequently contaminated with fecal bacteria, but contamination prevalence is highly variable between studies. Our finding of frequent contamination of hands is supported by previous work demonstrating the role of hands in connecting sources of fecal contamination (e.g. soil, floor, drain) with opportunities for subsequent exposures (e.g. mouth, bath, handwashing) (99).

### Country income

Country income level, as defined by the World Bank’s four income groups – low, lower-middle, upper-middle and high, was significantly correlated with hand contamination levels. Hands of people in low-income countries had significantly higher prevalence of *E. coli* and fecal coliforms than those in high-income countries. There were 28 countries included in this review (Figure S3), including 11 upper-middle/high income and 17 low/lower-middle income. Sub-Saharan Africa, South Asia, and North America were overrepresented (30, 22, and 17 studies, respectively) while South America and the Middle East/North Africa were underrepresented (5 and 2 studies, respectively). There were no studies from East Asia. The mean [95% Confidence Interval] estimate was 69% [48% - 58%] *E. coli* prevalence in low-income versus 6% [2% - 12%] in high-income (p = 5.74×10^−10^) and 85% [49% - 100%] fecal coliform prevalence in low-income versus 18% [10% - 28%] in high-income (p = 3.23×10^−13^) (Figure 1, Figure S7). We noted substantial between-study heterogeneities within each subgroup, as indicated by the high I^2^ values, which is an estimate of the proportion of observed variance attributable to study heterogeneity (*E. coli:* low-income: I^2^ = 91.3%; high-income: I^2^ = 99.6%; Fecal coliforms: low-income: I^2^ = 100%; high-income: I^2^ = 96%;). High heterogeneity suggests hand contamination prevalence is largely study or site-specific. The publication bias analysis showed major asymmetry to the right (Figure S5); there are more low-precision studies measuring low prevalence than low-precision studies measuring high prevalence.

**Figure 1.**
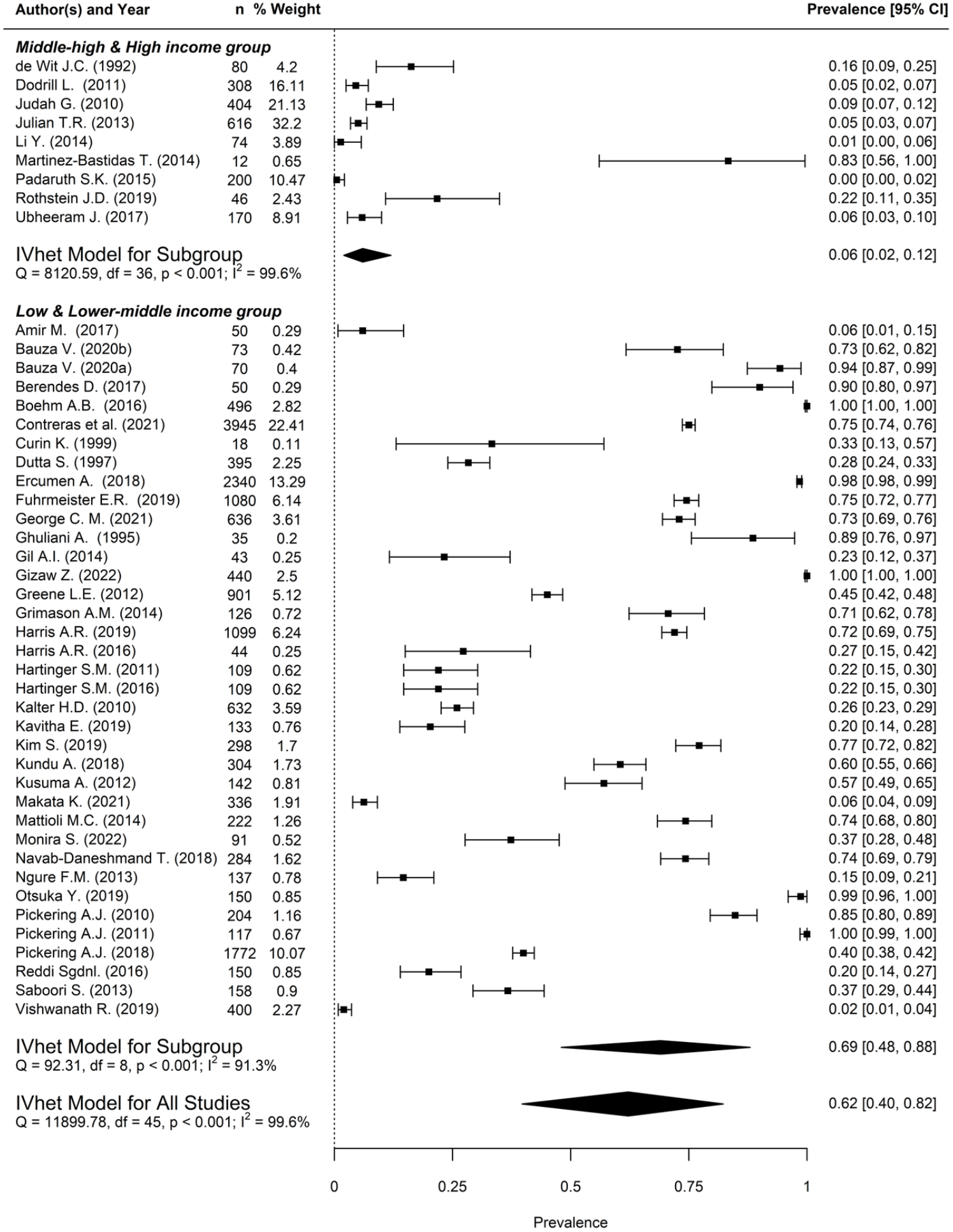
*E. coli* prevalence on hands by country income level. Pooled estimates for all studies and subgroups were evaluated with an inverse variance heterogeneity (IVhet) model.

The observed trend that hand contamination is generally more prevalent in low-income countries than high-income countries may be attributable to differences in access to water, sanitation, and hygiene, including hand washing facilities with soap and water. Access to handwashing facilities is correlated with piped water access and with sociodemographic index (SDI; a composite measure including income per capita, education, and fertility) (100). Previous work has also demonstrated that hands are quickly re-contaminated after handwashing in many tropical low-income settings (within minutes), potentially due to high levels of fecal contamination on surfaces and in soils in the domestic environment (24,70).

### Urban compared to rural areas

There was no significant difference in *E. coli* prevalence between urban and rural areas (p = 0.110; Figure S9, Figure 2). The publication bias analysis (Figure S6) for the low/lower-middle income subgroup showed minor asymmetry between the urban and rural groups. Notably, comparisons of hand contamination between urban and rural settings could only be conducted for *E. coli* prevalence in low/lower-middle income countries because there were no studies that measured *E. coli* in rural areas in upper-middle/high income countries and too few studies available to assess differences in fecal coliform levels (low/lower middle income: 2 studies in urban, 7 studies in rural; upper-middle/high income: 10 studies in urban, 0 studies in rural). Although WASH coverage levels are typically higher in urban than in rural areas, there are substantial intra-urban access inequalities (101). This could explain why we observed no difference between urban versus rural hand contamination prevalence levels(102). Many studies included in this review from urban low-income settings were conducted in informal settlements in contrast to more established, wealthier urban communities. Populations living in densely populated, low-income communities often share sanitation facilities. Shared sanitation facilities have been shown to be less likely to be clean (in terms of presence of fecal matter, number of flies, and smell) than private facilities (103). Another potential explanation is that population density (persons per km^2^) is correlated with higher rates of environmental fecal contamination, as shown in Egypt where *E. coli* drinking water contamination was increased in areas with the highest population densities (104).

**Figure 2:**
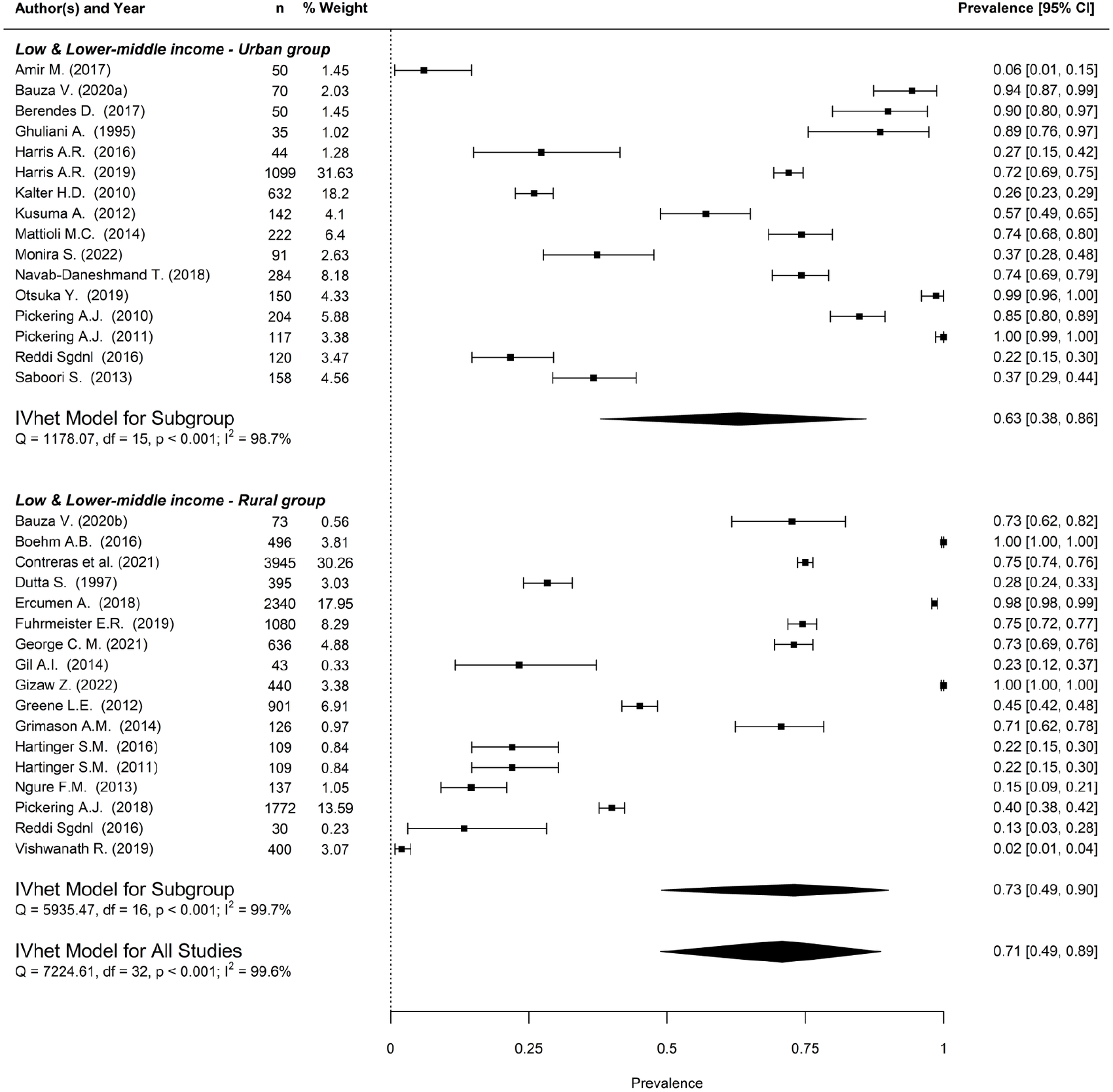
*E. coli* prevalence Urban vs. Rural in low/lower-middle income countries

### Adults compared to children

There were no clear differences in *E. coli* or fecal coliform prevalence between adults (16 years old and above) and children (birth to 15 years old) even when subgrouping by income level (Figures S27, S29, S31, S33, S35) (p = 0.21 (E. coli all), 0.09 (E. coli low-income only), 0.78 (fecal coliform all), 0.45 (fecal coliforms low-income only), 0.52 (fecal coliforms high income only)). There were also no significant differences in prevalence between the hands of children under 5 years old and adult hands (p = 0.23 (E. coli all), 0.47 (E. coli low-income only), 0.66 (fecal coliform all), 0.56 (fecal coliforms high-income only)). A potential explanation is that hand hygiene behavior between adults and children is highly correlated (105). Hand contamination is also strongly influenced by environmental contamination. Similar levels of contamination between adults and children could be a result of how difficult it is to keep hands free of fecal contamination in high-disease burden settings lacking safely managed sanitation (24,54,70). The results align with other studies that have found no difference in contamination levels with direct comparisons between adults and children (25,45).

### Climate classifications

*E. coli* prevalence was higher in tropical areas (classification A; 0.69 [95% CI 0.41, 0.93]) than in temperate areas (classification C; 0.28 [95% CI 0.09, 0.51]) (p = 0.015). Two fecal coliform comparisons had significant differences (tropical areas, A vs. temperate areas, C; tropical areas, A vs. dry areas, B), but country income was a confounding factor for both comparisons. A recent study in Kenya found that high 7-day temperature was associated with decreased *E. coli* levels on hands (106). A review on the associations between ambient temperature and enteric infections found increased risks in bacterial enteric infections and decreased risks in viral infections for every 1ºC temperature rise (107). Our results also suggest that temperature may influence hand contamination levels. This analysis could be improved by taking into account the time of year a study was conducted (rainy season, dry season, hot, or cold months), but this information was not readily available.

### Gender

Although analysis by gender was intended, very few of the identified studies reported gender. Three studies reported stratified prevalence values for males and females, while 5 additional studies reported a comparison between genders. Within 7 of these studies, which all enrolled school children, 5 found that male students had higher levels of fecal indicator bacteria, one found that females had higher levels of fecal indicator bacteria, and one found no statistical difference between genders. Potential reasons for higher rates of fecal contamination on male hands relative to female hands include differences in hygiene and/or interactions with the environment (playing sports and/or in soil) (108–110).

### Sampling methods

Among the 84 studies, 51 (61%) used hand rinse samples, 19 (23%) used swab samples, 13 (15%) used impressions, and 1 study (1%) did not report the method used (see SI for method details). Most studies that employed a culture-based method used membrane filtration or IDEXX Quanti-Tray; however, the lower detection limit ranged from 1 CFU/hand to 175 CFU/hand (when reported). Rinse samples had a higher mean prevalence for *E. coli* (0.69 [95% CI 0.46, 0.89]) than swab samples (0.24 [95% CI 0.00, 0.60]). Similarly, rinse samples had a higher mean prevalence for fecal coliform (0.84 [95% CI 0.35, 1.00]) than impression samples (0.27 [95% CI 0.08, 0.48]) (*E. coli* rinse vs. swab: p = 0.03; Fecal coliform rinse vs. impression: p = 2.32 ×10^−05^; Figures S37-S43). These findings show that hand rinse sampling may be more sensitive at recovering fecal indicator bacteria from hands than swabs and possibly impression plates, in line with prior research and what has been reported for hand microbiome studies (111,112). Standardization of sampling methods could improve the sensitivity of studies and facilitate the identification and assessment of risk factors for hand contamination.

## Conclusion

Hand contamination was highest in low/lower-middle income countries where diarrheal and enteric infections are also more common. In low/middle income countries, hand contamination was equally prevalent in rural and urban communities, emphasizing the need for hand hygiene across both settings. Given the role of hands in transferring fecal microbes between people and the environment, we suggest hands can be viewed as sentinel indicators of human exposure to enteric pathogens. However, there is a need to develop and implement standardized sampling and analysis methods, including molecular methods to detect specific pathogens on hands, as most studies have been limited to measuring fecal indicator bacteria with culture-based methods. Standardized molecular methods to multiplex detection of clinically relevant pathogens on hands would improve our understanding of the role of hands in human exposure to protozoan, bacteria, and viral pathogens and help to identify factors influencing hand contamination.

## Data Availability

All data included in the systematic review are publicly available in published manuscripts.

## Supporting Information

Detailed description of methods, all results, PRISMA-P checklist. table of all papers included. Data extracted from papers included in this review is publicly available on Open Science Framework at this link: https://osf.io/j6wb4/.

