## Supplemental Information for "Hands are frequently contaminated with fecal bacteria and enteric pathogens globally: A systematic review and meta-analysis"

Affiliations:
[1] Department of Civil and Environmental Engineering, Tufts University, Medford, MA, 02155
[2] Eawag, Swiss Federal Institute of Aquatic Science and Technology, 8600, Dubendorf, Switzerland
[3] ETH Zurich, Swiss Federal Institute of Technology, 8092, Zurich, Switzerland
[4] Department of Public Health and Family Medicine, Tufts Medical School, Tufts University, Boston, MA, 02111
[5] Friedman School of Nutrition Science and Policy, Tufts University, Boston, MA, 02111
[6] Swiss Tropical and Public Health Institute, 4051, Basel, Switzerland
[7] University of Basel, 4001, Basel, Switzerland
[8] Department of Civil and Environmental Engineering, University of California, Berkeley
[9] Chan Zuckerberg Biohub, San Francisco, CA

#Authors contributed equally

### Included in this supplemental information


Materials and Methods

- Additional details on search methods
- Figure S1: F-diagram
- Table S1 Hand sampling method

Limitations

Results

- Figure S2: Systematic review search selection process
- Figure S3: Map showing the number of studies per country included in the review
- Figure S4: Sample size of studies included in the review
- Table S2: Statistical Test (z-test) Results Summary
- Doi plots for manuscript figures
    - Figure S5: Doi plot of E. coli prevalence for all studies (for Figure 1 in the manuscript)
    - Figure S6: Doi plot of E. coli prevalence in low/lower-middle income countries (for Figure 2 in the manuscript)
- Country income
    - Figure S7: Fecal Coliform prevalence in low/lower-middle income vs. upper-middle/high income countries
    - Figure S8: Doi plot of Fecal Coliform prevalence in low/lower-middle income vs. upper-middle/high income countries
- Urban vs. rural
    - Figure S9: *E. coli* prevalence in urban v. rural area (all studies)
    - Figure S10: Doi plot of *E. coli* prevalence in urban v. rural area (all studies)
    - Figure S11: Fecal Coliform prevalence in rural vs. urban areas
    - Figure S12: Doi plot of Fecal Coliform prevalence in rural vs. urban areas
- Climate
    - Figure S13: *E. Coli* Prevalence in Climate Group A vs Group B
    - Figure S14: Doi plot for *E. Coli* Prevalence in Climate Group A vs Group B
    - Figure S15: *E. Coli* Prevalence Climate Group B vs Group C
    - Figure S16: Doi plot for *E. Coli* Prevalence Climate Group B vs Group C
    - Figure S17: *E. coli* Prevalence Climate Group A vs Group C
    - Figure S18: Doi plot for *E. coli* Prevalence Climate Group A vs Group C
    - Figure S19: *E. coli* Prevalence Climate Group A vs Group C – low and lower-middle income group
    - Figure S20: Doi plot of *E. coli* Prevalence Climate Group A vs Group C – low and lower-middle income group
    - Figure S21: Fecal Coliform Prevalence Climate Group A vs Group B

    - Figure S22: Doi plot of Fecal Coliform Prevalence Climate Group A vs Group B
    - Figure S23: Fecal Coliform Prevalence Climate Group A vs Group C
    - Figure S24: Doi plot of Fecal Coliform Prevalence Climate Group A vs Group C
    - Figure S25: Fecal Coliform Prevalence Climate Group B vs Group C
    - Figure S26: Doi plot for Fecal Coliform Prevalence Climate Group B vs Group C
- Adults vs. Children
    - Figure S27: *E. Coli* Prevalence for Adults vs. Children
    - Figure S28: Doi plot of *E. Coli* Prevalence for Adults vs. Children
    - Figure S29: *E. Coli* Prevalence Adults vs. Children in low/lower-middle income countries
    - Figure S30: Doi plot of *E. Coli* Prevalence Adults vs. Children in low/lower-middle income countries
    - Figure S31: Fecal Coliform Prevalence for Adults vs. Children
    - Figure S32: Doi plot of Fecal Coliform Prevalence for Adults vs. Children
    - Figure S33: Fecal Coliform Prevalence Adults vs. Children in low/lower-middle income countries
    - Figure S34: Doi plot of Fecal Coliform Prevalence Adults vs. Children in low/lower-middle income countries
    - Figure S35: Fecal Coliform Prevalence Adults vs. Children in middle-high/high income countries
    - Figure S36: Doi plot of Fecal Coliform Prevalence Adults vs. Children in middle-high/high income countries
    - Figure S37: *E. Coli* Prevalence Adults vs. Children Under 5
    - Figure S38: Doi plot of *E. Coli* Prevalence Adults vs. Children Under 5
    - Figure S39: *E. Coli* Prevalence Adults vs. Children Under 5 in low/lower-middle income countries
    - Figure S40: Doi plot of *E. Coli* Prevalence Adults vs. Children Under 5 in low/lower-middle income countries
    - Figure S41: Fecal Coliform Prevalence Adults vs. Children Under 5
    - Figure S42: Doi plot of Fecal Coliform Prevalence Adults vs. Children Under 5
    - Figure S43: Fecal Coliform Prevalence Adults vs. Children Under 5 in upper-middle/high income countries
    - Figure S44: Doi plot of Fecal Coliform Prevalence Adults vs. Children Under 5 in upper-middle/high income countries
- Methods
    - Figure S45: *E. Coli* prevalence in rinse vs swab methods
    - Figure S46: Doi plot of *E. Coli* prevalence in rinse vs swab methods
    - Figure S47: *E. Coli* prevalence in rinse vs swab methods in low/lower-middle income countries
    - Figure S48: Doi plot of *E. Coli* prevalence in rinse vs swab methods in low/lower-middle income countries

- Figure S49: *E. Coli* prevalence in rinse vs swab methods in high/upper-middle income countries
- Figure S50: Doi plot of *E. Coli* prevalence in rinse vs swab methods in high/upper-middle income countries
- Figure S51: Fecal Coliform prevalence in rinse vs impression methods
- Figure S52: Doi plot of Fecal Coliform Prevalence in rinse vs impression methods

- Limitations

PRISMA-P Checklist for Environmental Contamination

Table S3: Papers included in the systematic review

References

### Materials and Methods

Two independent reviewers completed initial title and abstract screening, RS and TRJ in March 2018, MEC and TRJ in June 2020, and HW and TRJ in September 2022. A third reviewer (AJP) resolved discrepancies. Full text review and data extraction was completed by one reviewer and checked by a second (up to March 2018: RS, checked by MEC; up to June 2020: MEC, checked by HW; up to September 2022: MEC and HW both reviewed/extracted and checked). Studies were included if a subset of the data met the inclusion criteria, with only the subset of relevant results included in the review. Two studies known to meet inclusion criteria by the author team were also included. Grey literature and white papers were excluded; we did not attempt to contact authors for missing data. Only papers in English and French were included.

Concentration values were synthesized to the unit $\log_{10}$CFU/hand. For this analysis, quantification of bacteria in units of most probable number (MPN) was treated as equivalent to quantification using units of colony forming unit (CFU). This assumption introduces some error because MPN is generally characterized by higher intra-sample variability and "somewhat higher" estimates than CFU (1). However, the analytical difference between methods is generally thought to be much lower than the intrinsic variability of the assay (1–3). Additionally, to standardize results for studies reporting concentration per unit area instead of "per hand", an adult hand was assumed to be 160 $cm^2$ (4). Notably, no studies on child hand contamination reported results per unit area other than "per hand", so a child's hand estimate was not needed. Concentration units that could not be synthesized and were, therefore, not included in the summary table include median CFU/ml, inter-quartile range MPN/hand, gene copies/hand, and eggs/hand. This included 20 observations from 10 studies.

For the majority of studies, urban and rural classifications were a result of the investigators of each study making a designation. If an urban or rural classification was not presented for a study, we inferred a designation based on population descriptions or locations described. Country-level income data was used in the absence of accessible data on incomes of specific communities studied.

For fiscal year 2023, country income classifications (available via the *World Bank Data Help Desk*, https://datahelpdesk.worldbank.org/) were defined as follows: low-income economies as those with a GNI per capita of $1,085 or less in 2021; lower middle-income economies are those with a GNI per capita between $1,086 and $4,255; upper middle-income economies are those with a GNI per capita between $4,256 and $13.205; high-income economies are those with a GNI per capita of more than $13,205.

Statistical significance was defined as p-value <0.05.

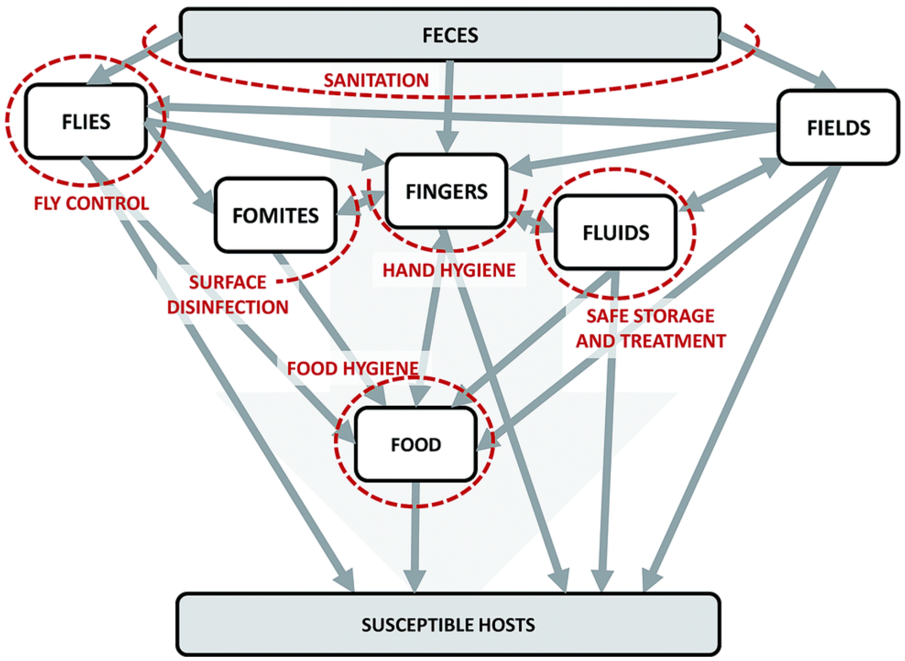


Figure S1: F-diagram (5)

Table S1: Hand Sampling methods (6)

| **Technique** | **Description** |
|---|---|
| Rinse | The whole hand or fingertips are placed in a glove or bag and rinsed and rubbed with a measured volume of sterile solution for a specific length of time |
| Swab | A swab is dipped in sterile solution and used to sample parts of the hand(s) |
| Impression | The fingertips are pressed directly onto selective agar media |

**Limitations**

The meta-analysis conclusions are impacted by limitations in both data collection and the underlying data set. First, the selection of keywords for the systematic review included both general keywords (e.g., “virus”, “helminth”) as well as specific keywords of known fecal indicators (“*E. coli*", “enterococci”). This selection may have been insufficient to capture all enteric pathogens and fecal indicators and may have biased the data set to overrepresent the historic fecal indicators and underrepresent novel or emerging pathogens and indicators. Second, heterogeneity in study designs, sample collection, and microbiological analysis amongst the identified studies may influence conclusions. For example, prevalence rates are likely biased by limits of detection such that studies using more sensitive methods may suggest higher rates of hand contamination. Unfortunately, limit of detection was not reported by a large proportion of studies, so formal analyses of these effects was not feasible. Additionally, quantification methods are characterized by variable levels of uncertainty, hindering interpretation of direct comparisons. For example, most probable number methods based on limited partitions are characterized by higher uncertainty than colony forming unit methods. Future studies of hand contamination should be encouraged to include methodological details sufficient to estimate methodological uncertainty and limit of detection, and corresponding impacts on prevalence rates and associated uncertainty.

**Results**

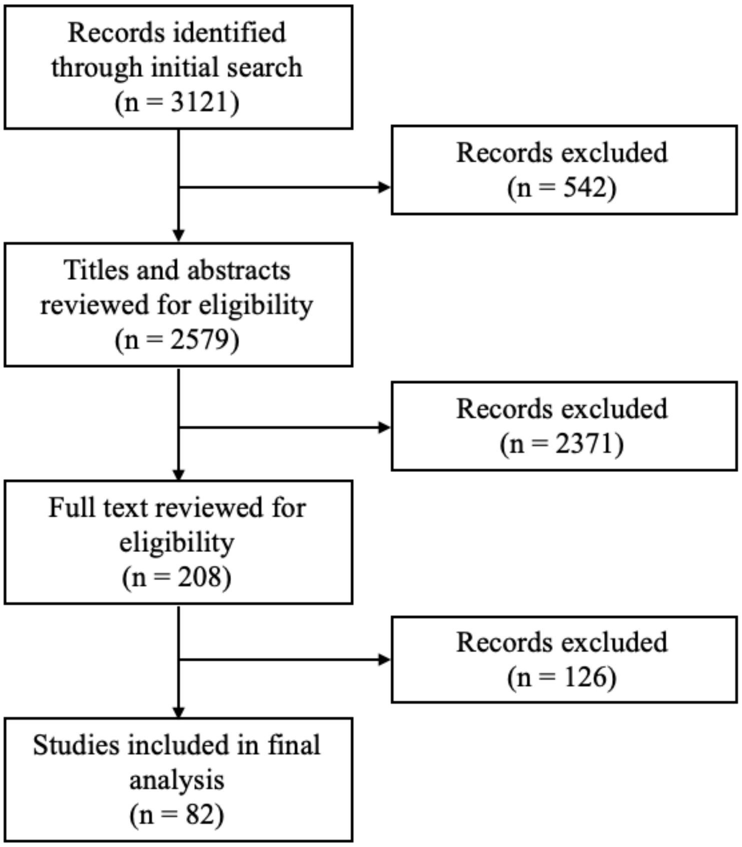


Figure S2: Systematic review search selection process (note that 2 additional studies were known and added by the authors. Therefore, final n = 84)

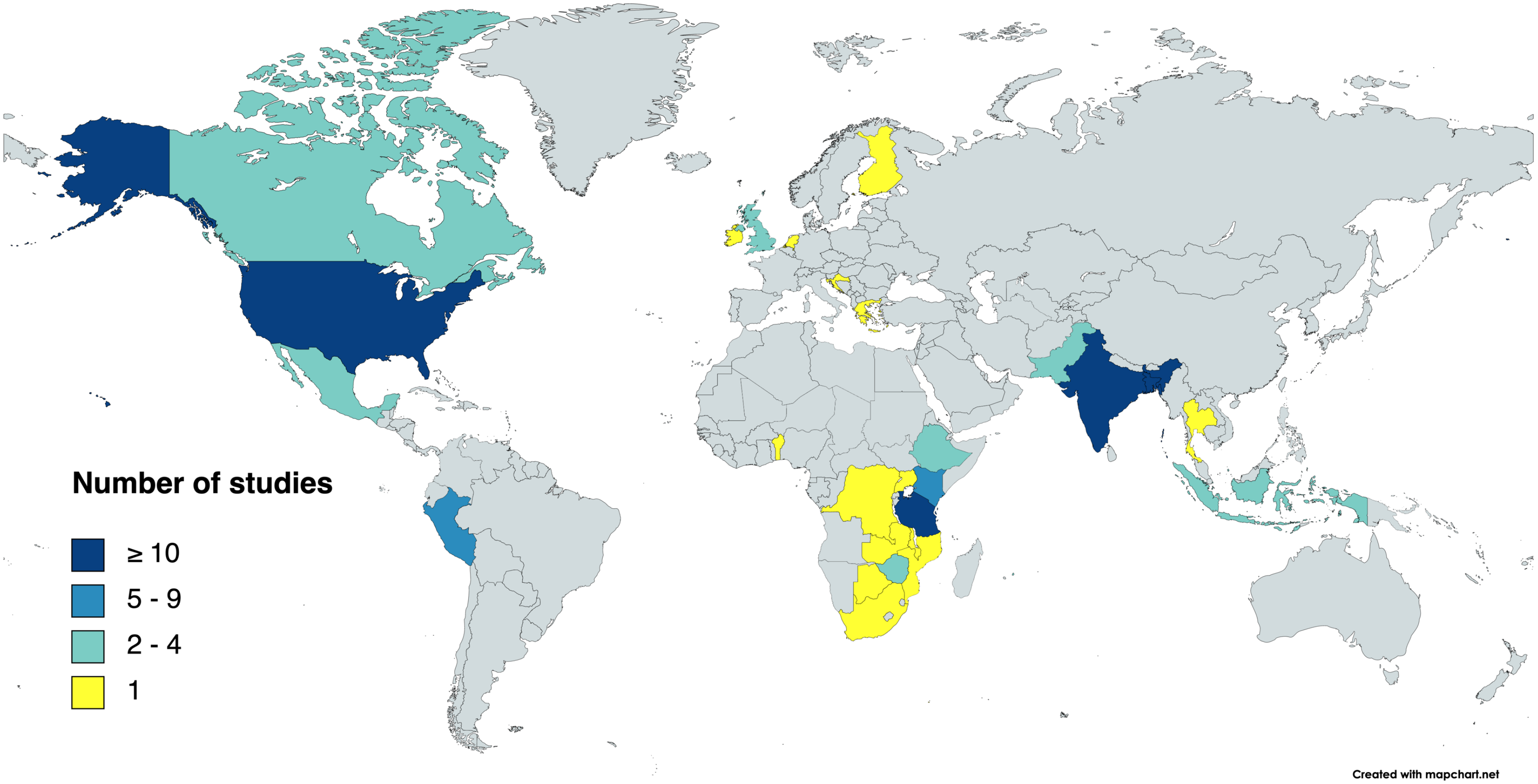


Figure S3: Map showing the number of studies per country included in the review

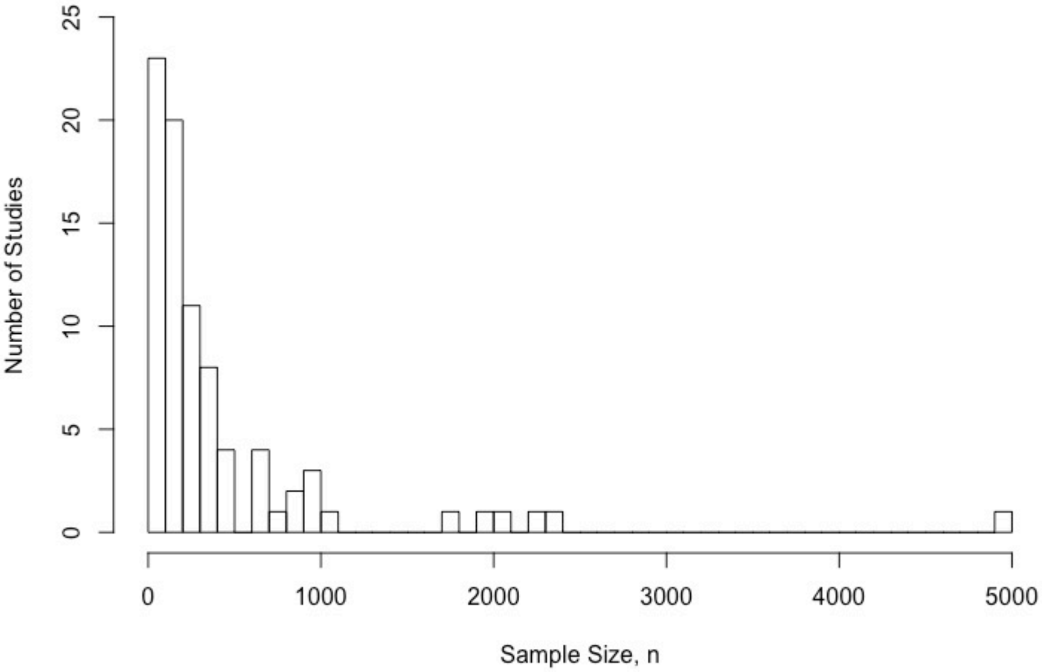


Figure S4: Sample size of studies included in the review (84 studies total)

For the interpretation of results from z-tests, it should be noted that the IVhet model accounts for between-study heterogeneity through a quasi-likelihood approach rather than a normal distribution. However, statistical methods to estimate the confidence interval of the pooled estimate of the IVhet model have not been proposed yet. The confidence interval of the IVhet estimator is generated assuming normality in MetaXL version 5.3.

Table S2: Statistical Test (z-test) Results Summary

| Subgroup | *E. coli* | | | Fecal Coliform | | |
|---|---|---|---|---|---|---|
| | All | Low income | High income | All | Low income | High income |
| Country Income | 5.74E-10 | na | na | 3.23E-13 | na | na |
| Urban v. Rural | 0.110 | 0.493 | * | 3.17E-11 | * | * |
| Adult v. Children | 0.209 | 0.096 | * | 0.775 | 0.455 | 0.521 |
| Adult v. Children under 5 | 0.235 | 0.468 | * | 0.663 | * | 0.564 |
| Climate A - B | 0.179 | * | * | 0.024 | * | * |
| Climate A - C | 0.016 | 0.204 | * | 0.000 | * | * |
| Climate B - C | 0.595 | * | * | 0.118 | * | * |
| Methods Rinse - Swab | 0.032 | 0.208 | 0.849 | * | * | * |
| Methods Rinse - Impression | * | * | * | 0.000 | * | * |
| Methods Swab - Impression | * | * | * | * | * | * |

*Note: yellow highlight means statistically significant (p-value < 0.05); orange highlight means significance was confounded by country income*

### Doi plots for manuscript figures

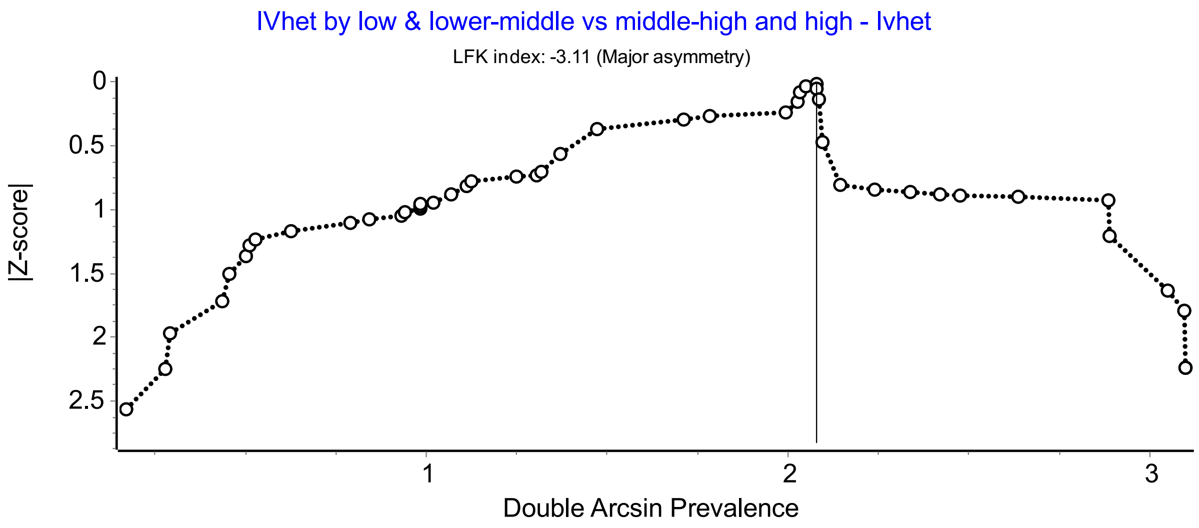


Figure S5: Doi plot of E. coli prevalence for all studies (for Figure 1 in the manuscript)

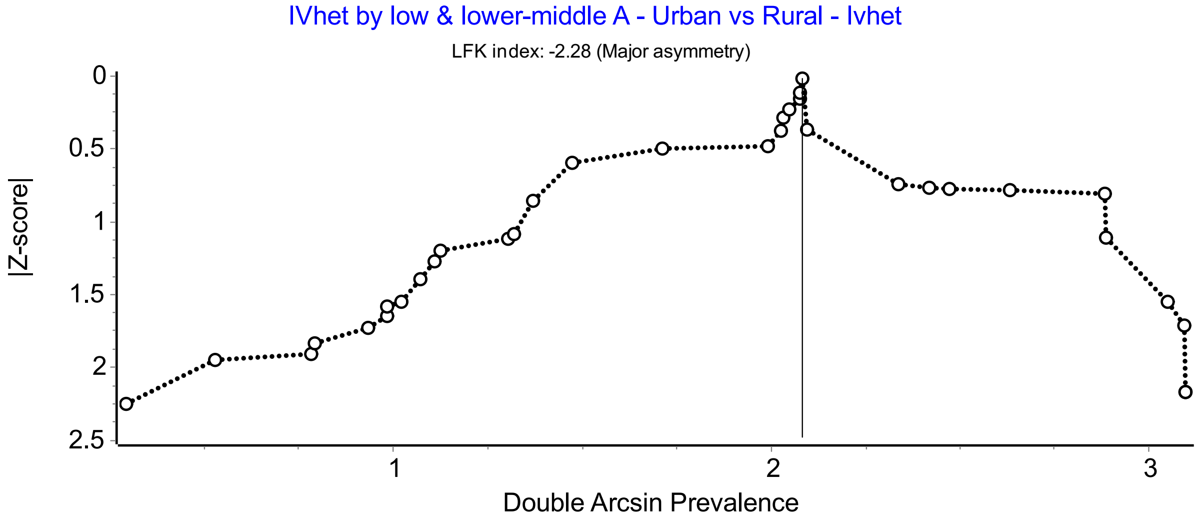


Figure S6: Doi plot of E. coli prevalence in low/lower-middle income countries (for Figure 2 in the manuscript)

### Country Income

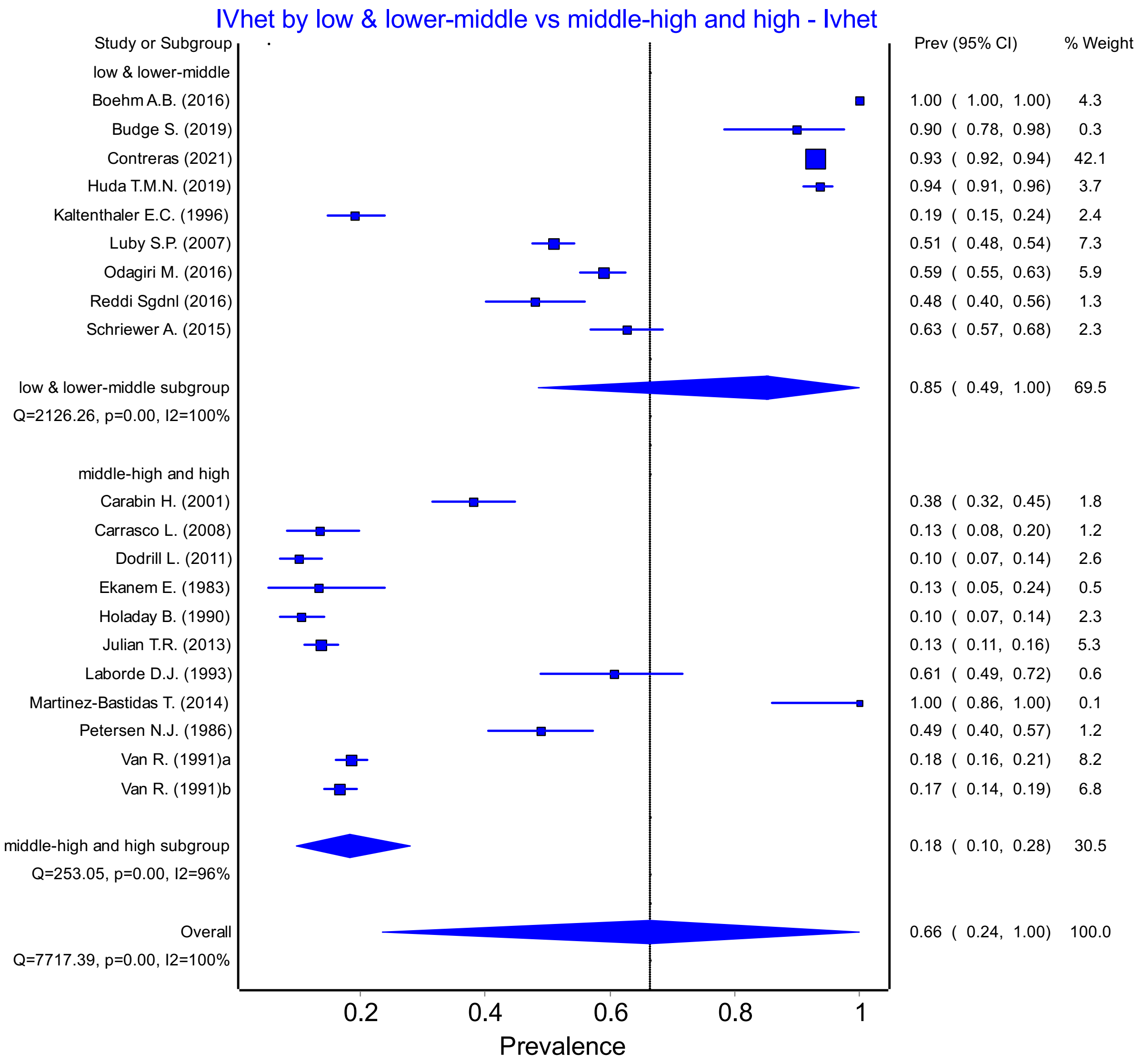


Figure S7: Fecal Coliform prevalence in low/lower-middle income vs. upper-middle/high income countries

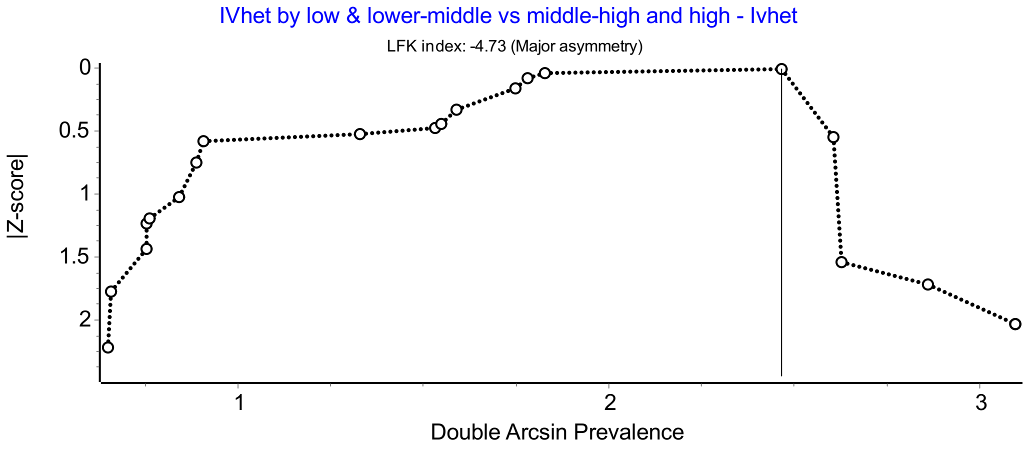


Figure S8: Doi plot of Fecal Coliform prevalence in low/lower-middle income vs. upper-middle/high income countries

### Urban vs. Rural

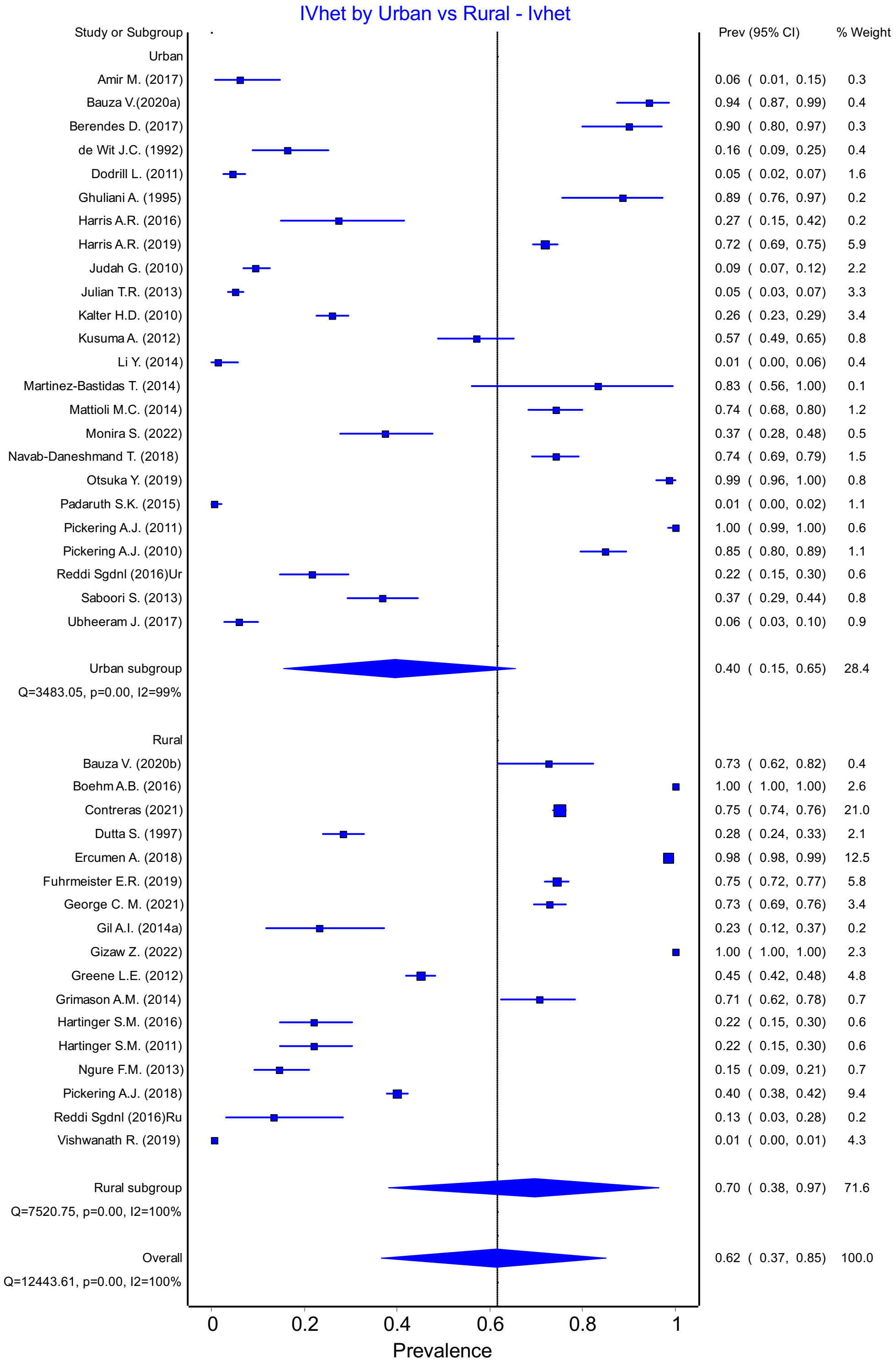


Figure S9: *E. coli* prevalence in urban v. rural area (all studies)

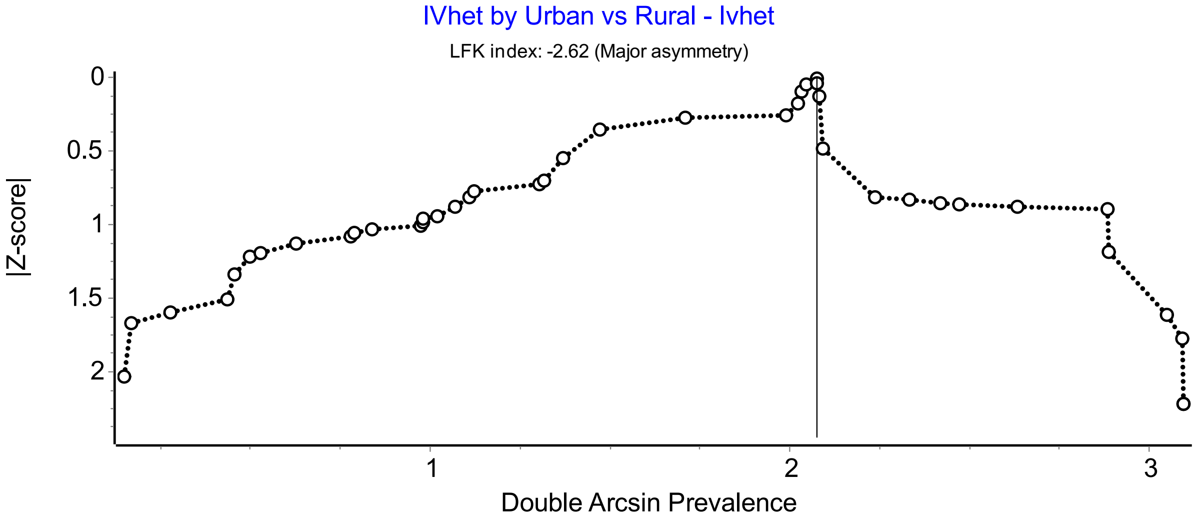

Figure S10: Doi plot of *E. coli* prevalence in urban v. rural area (all studies)

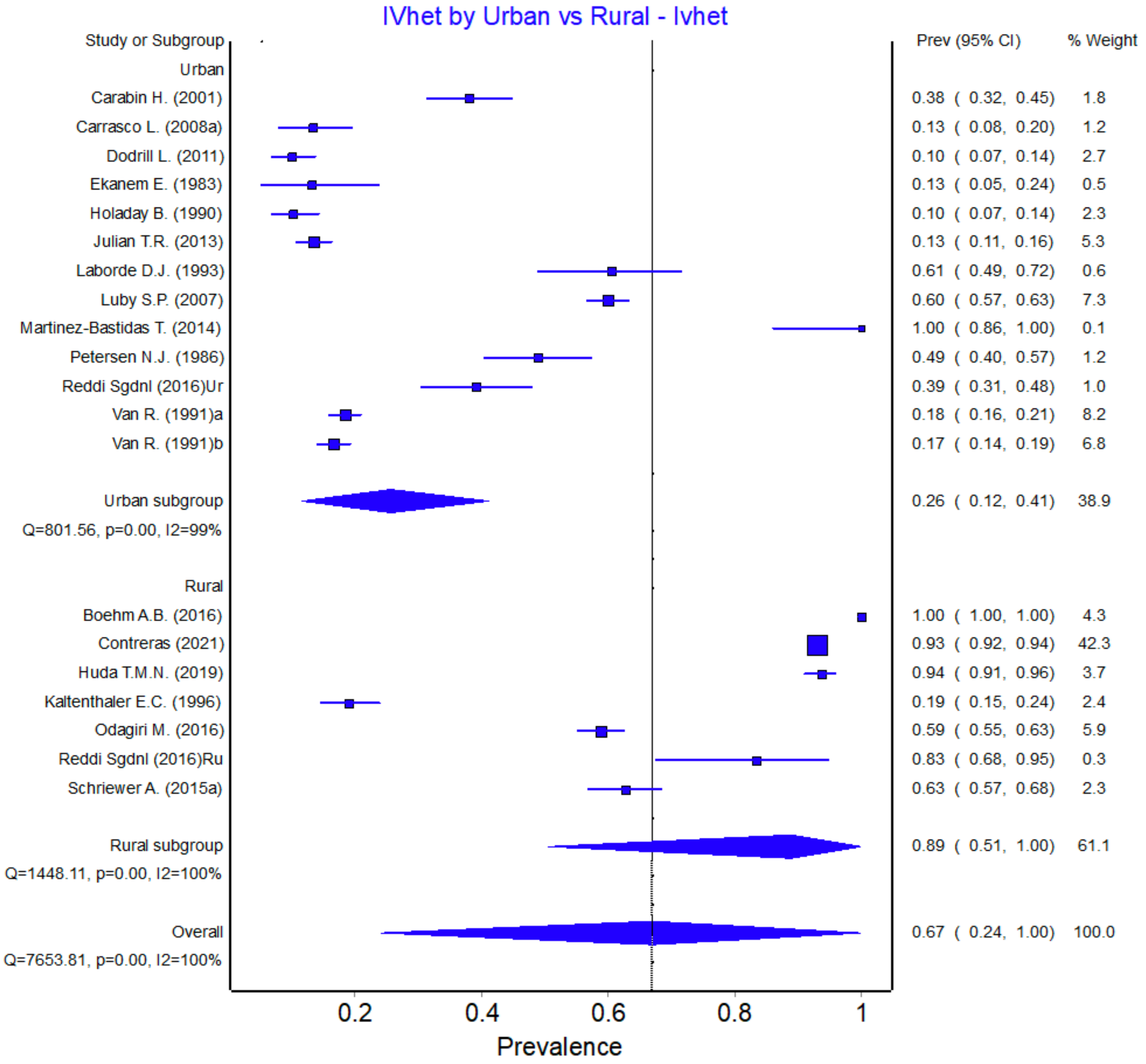


Figure S11: Fecal Coliform prevalence in rural vs. urban areas

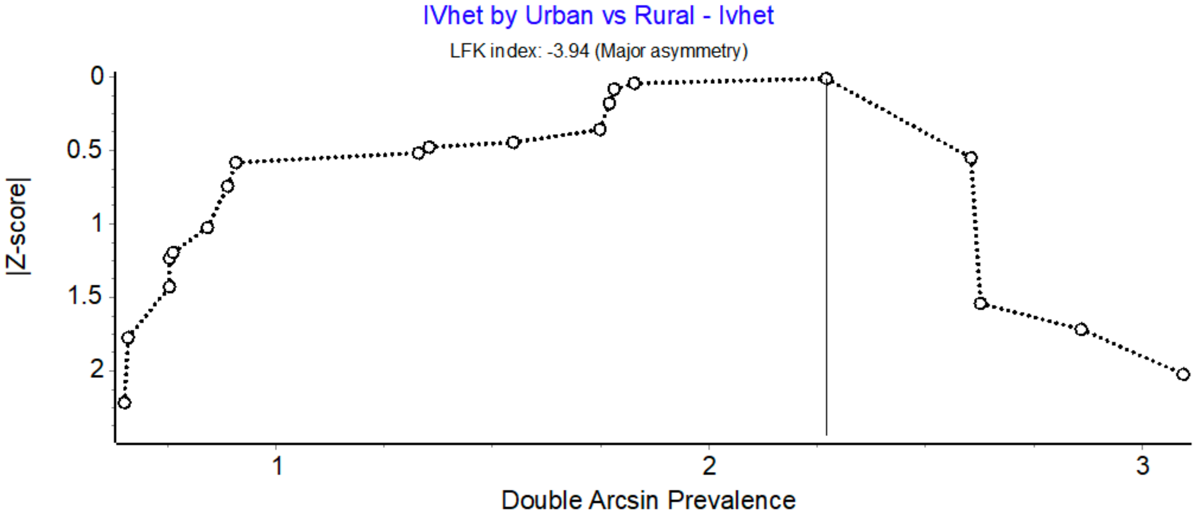


Figure S12: Doi plot of Fecal Coliform prevalence in rural vs. urban areas

### Climate

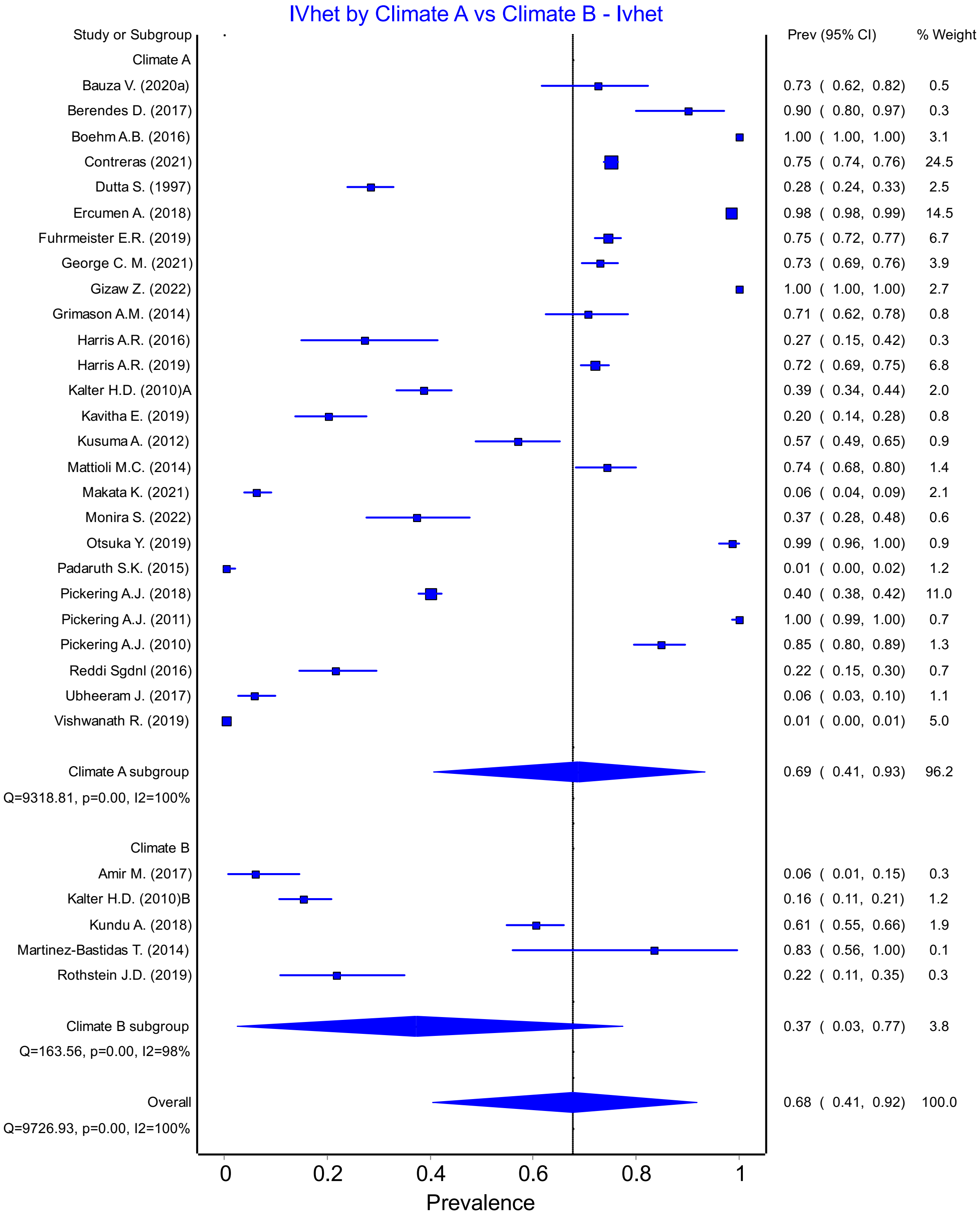


Figure S13: *E. Coli* Prevalence in Climate Group A vs Group B

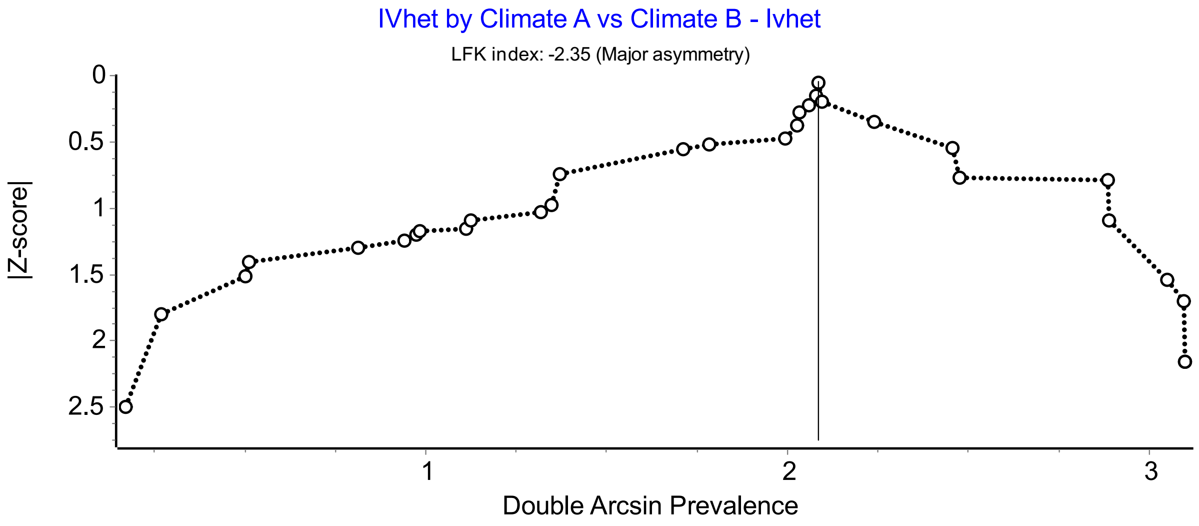


Figure S14: Doi plot for *E. coli* Prevalence Climate Group A vs Group B

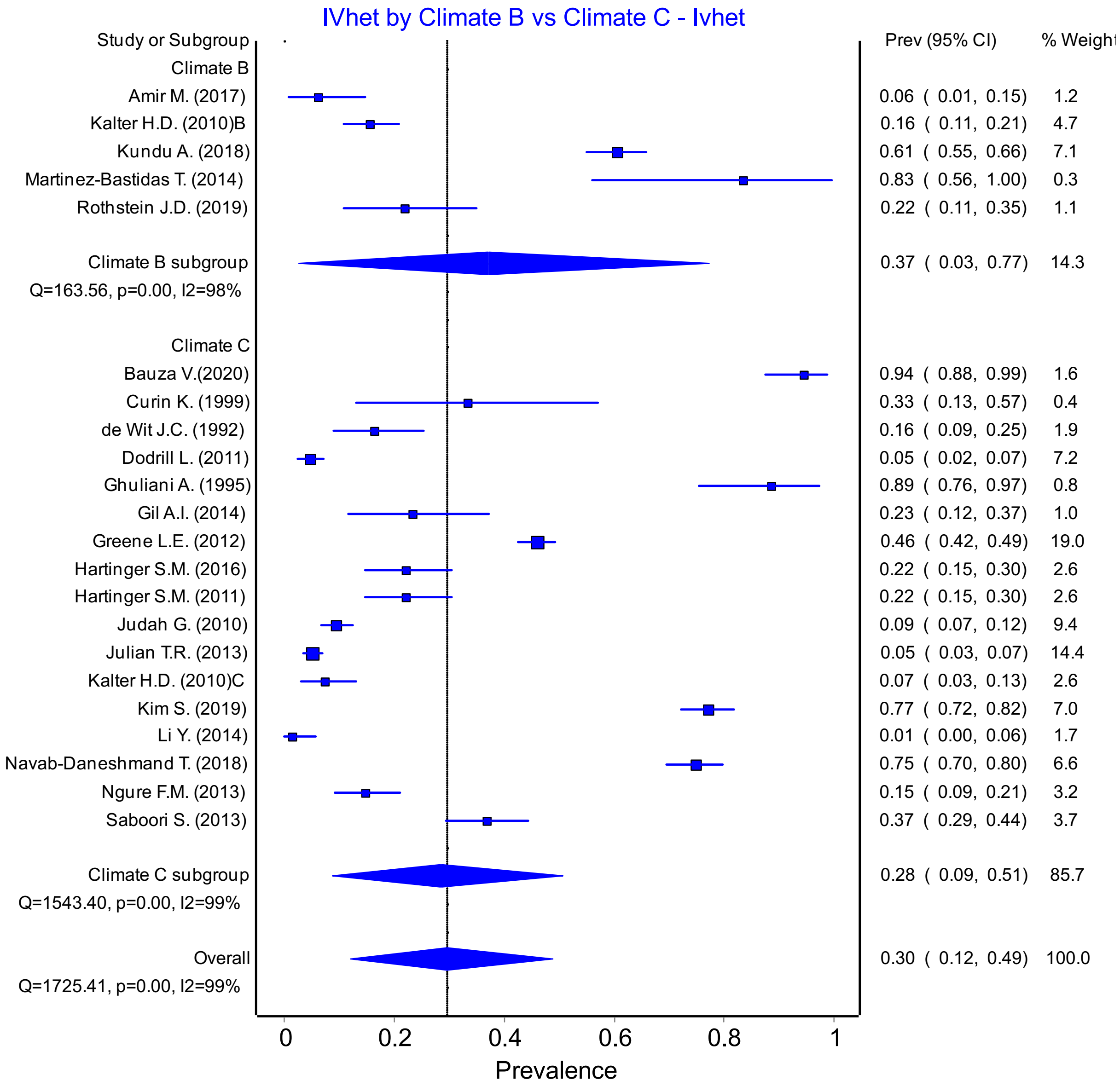


Figure S15: *E. Coli* Prevalence Climate Group B vs Group C

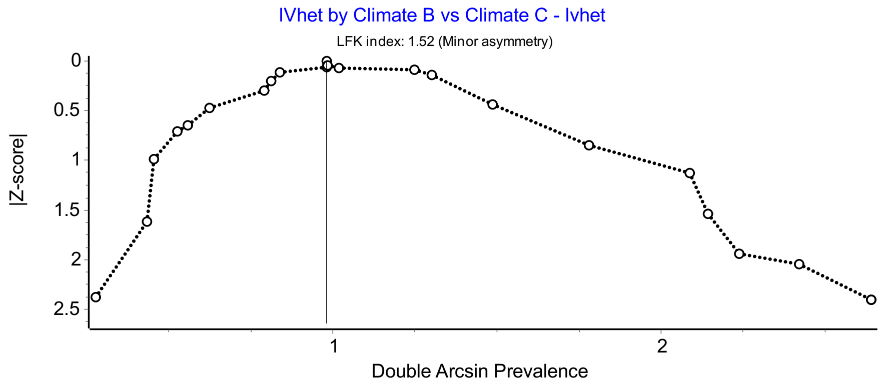


Figure S16: Doi plot for *E. coli* Prevalence Climate Group B vs Group C

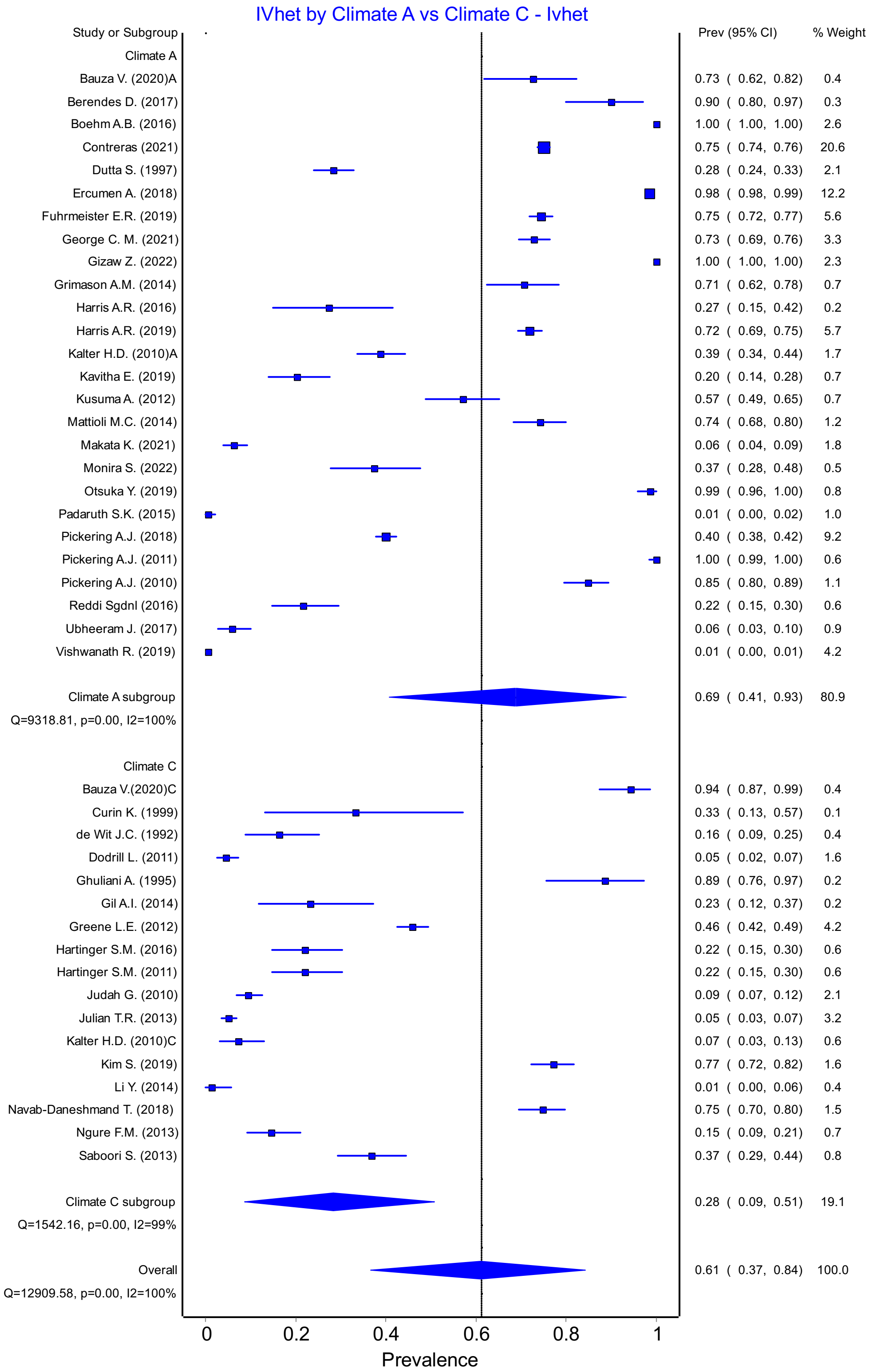


Figure S17: *E. coli* Prevalence Climate Group A vs Group C

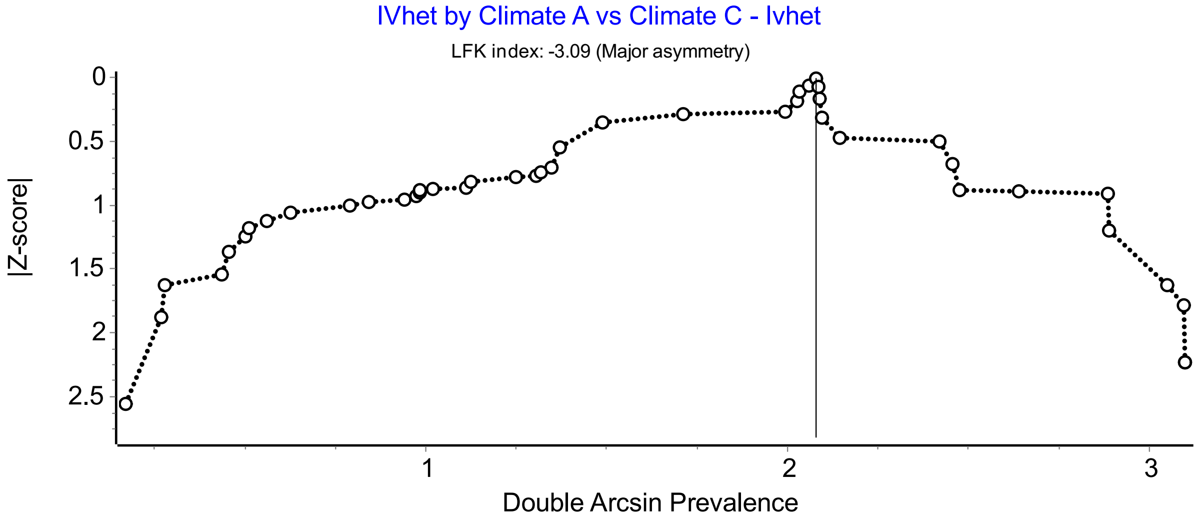


Figure S18: Doi plot for *E. coli* Prevalence Climate Group A vs Group C

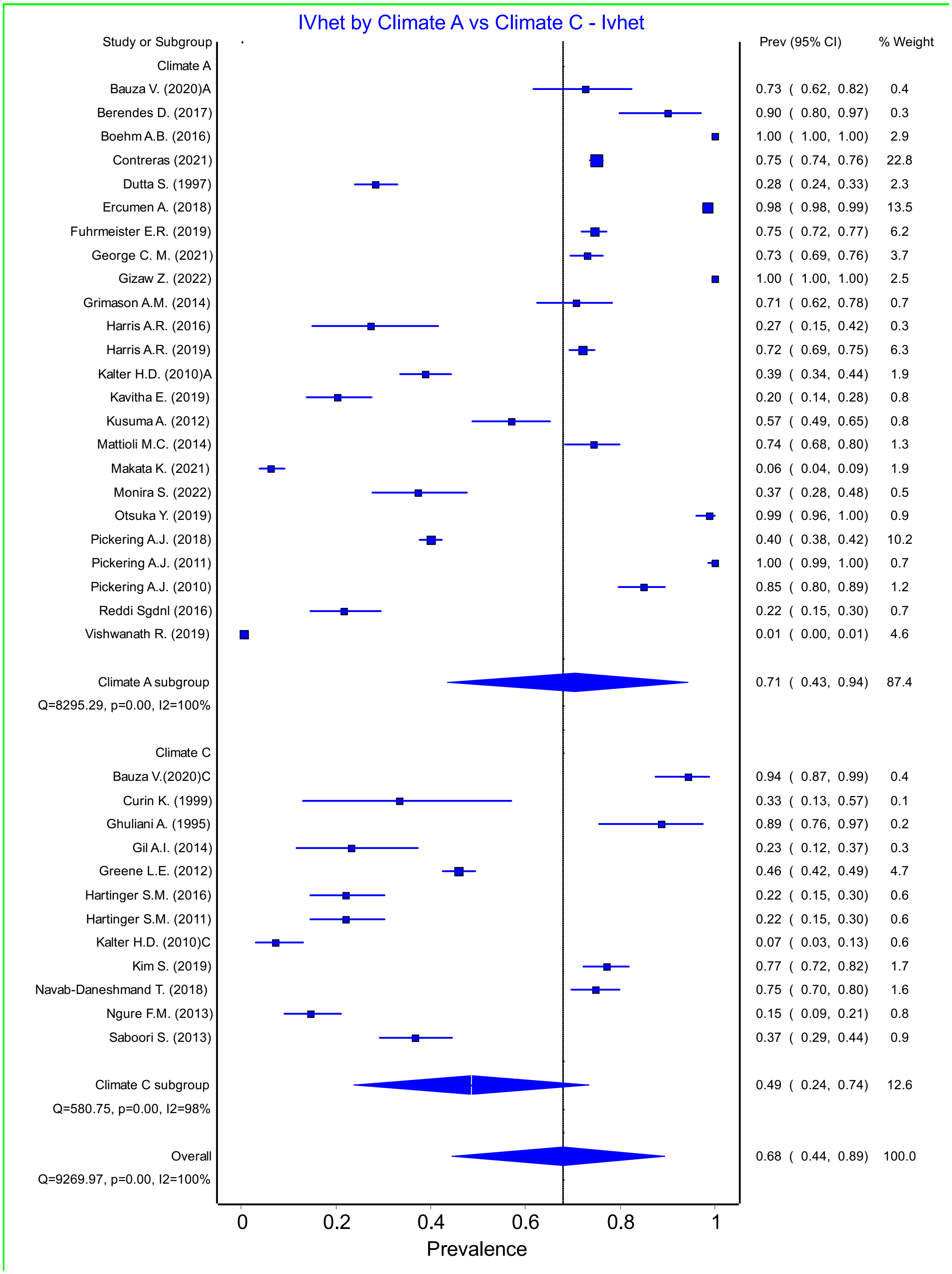


Figure S19: *E. coli* Prevalence Climate Group A vs Group C – low and lower-middle income group

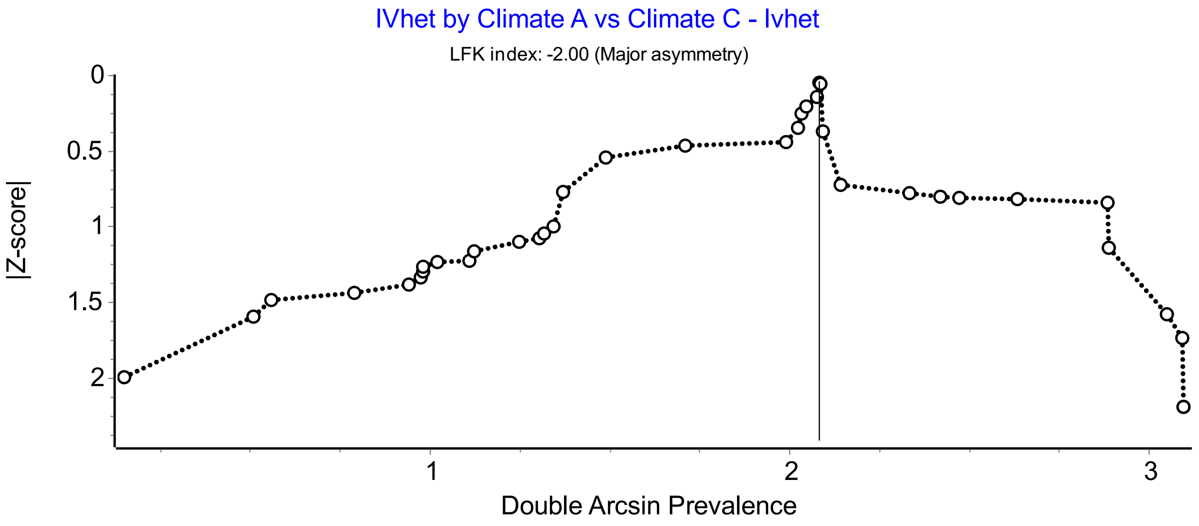


Figure S20: Doi plot of *E. coli* Prevalence Climate Group A vs Group C – low and lower-middle income group

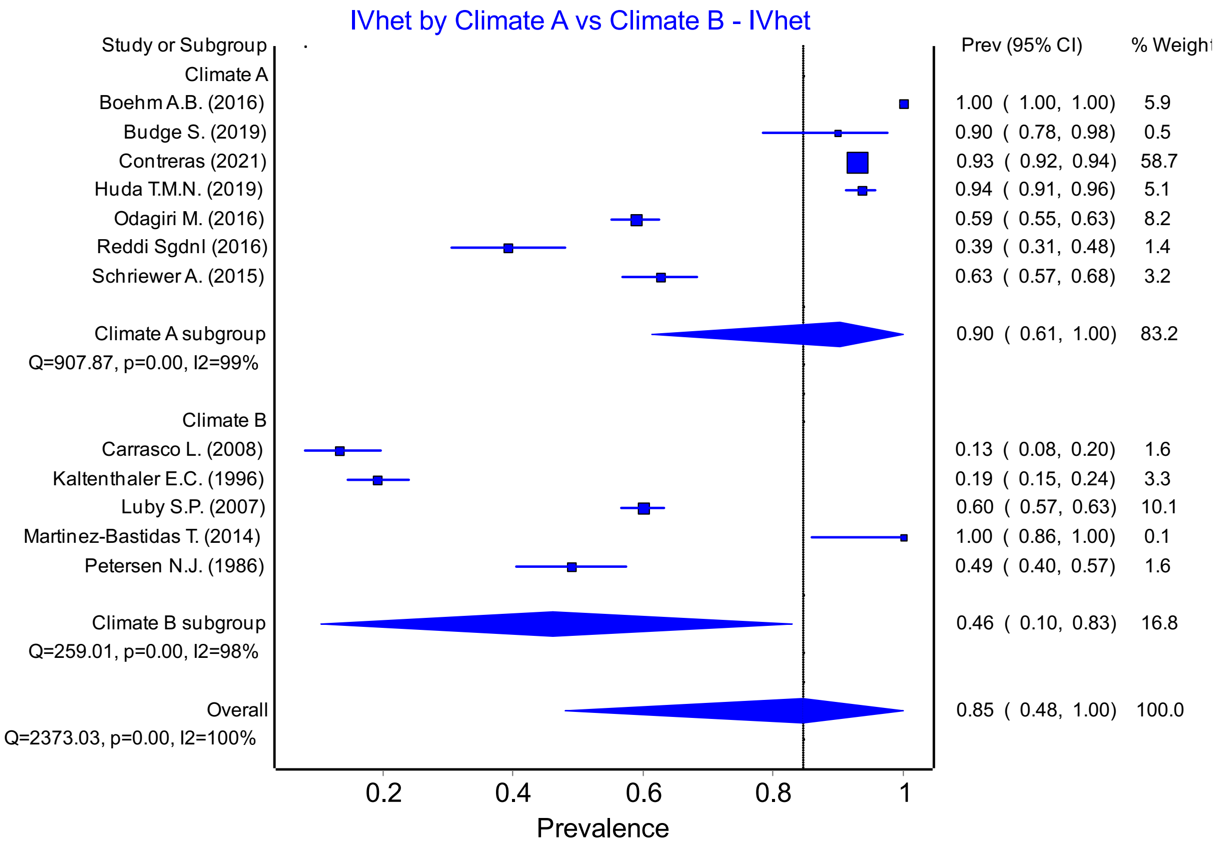


Figure S21: Fecal Coliform Prevalence Climate Group A vs Group B

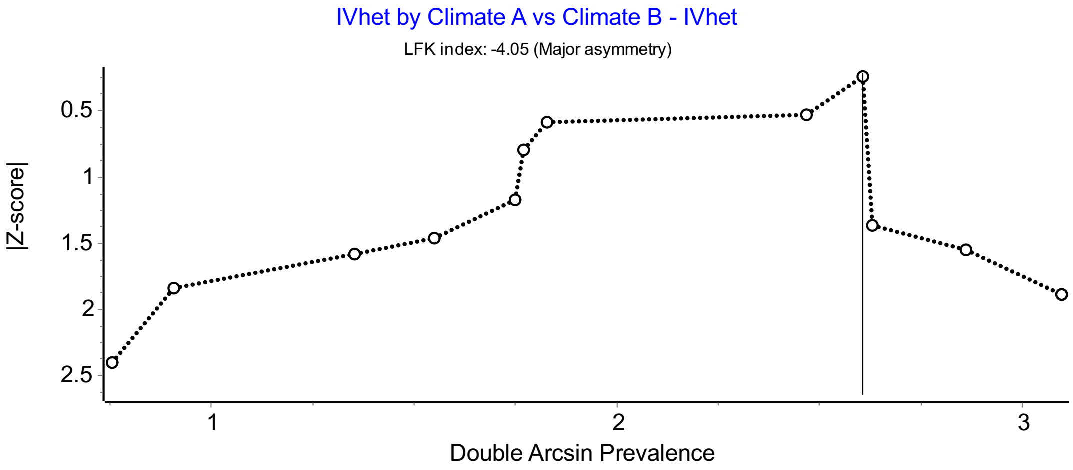


Figure S22: Doi plot of Fecal Coliform Prevalence Climate Group A vs Group B

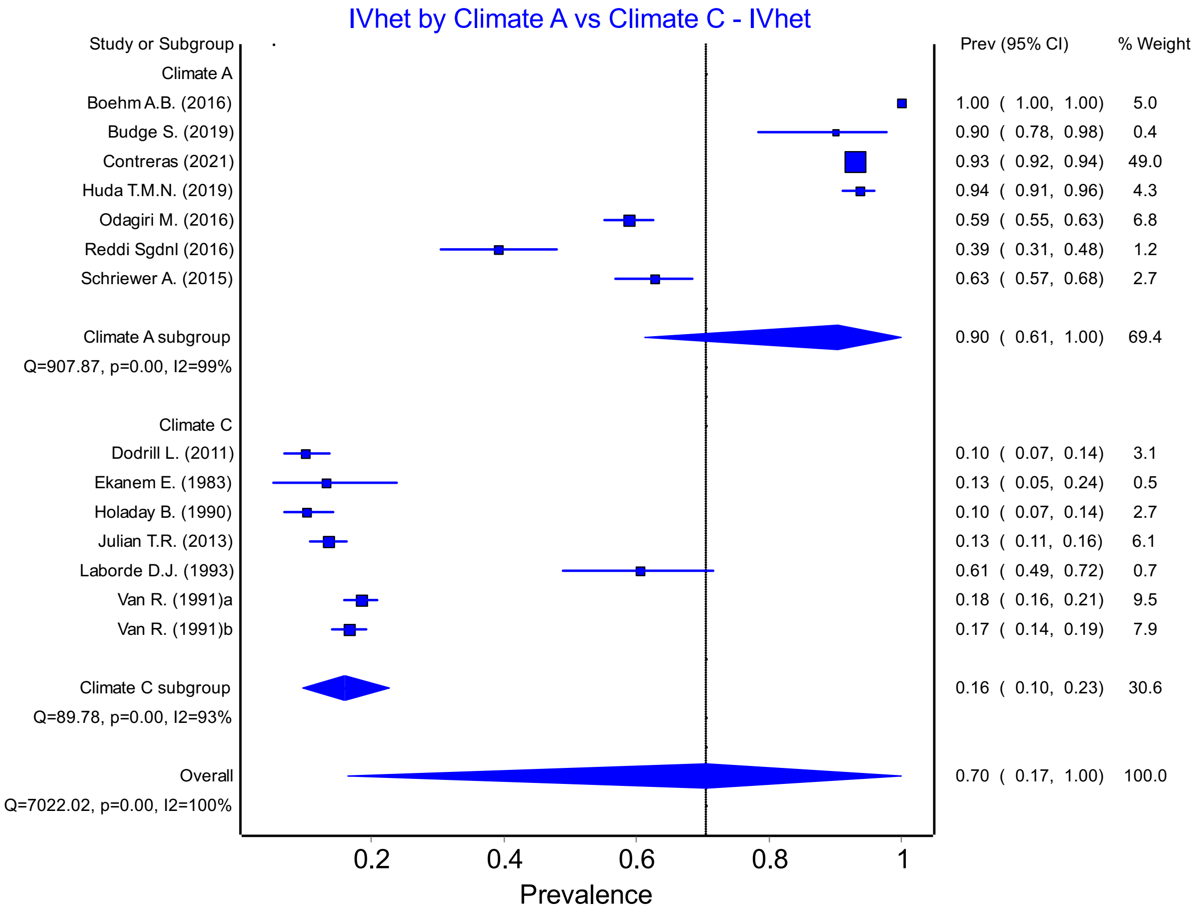


Figure S23: Fecal Coliform Prevalence Climate Group A vs Group C

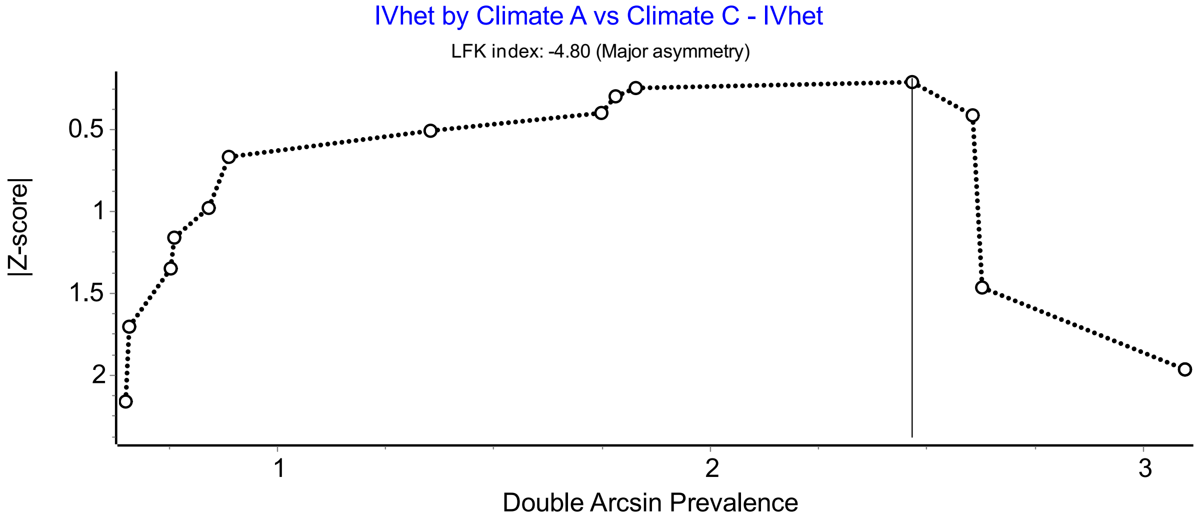


Figure S24: Doi plot of Fecal Coliform Prevalence Climate Group A vs Group C

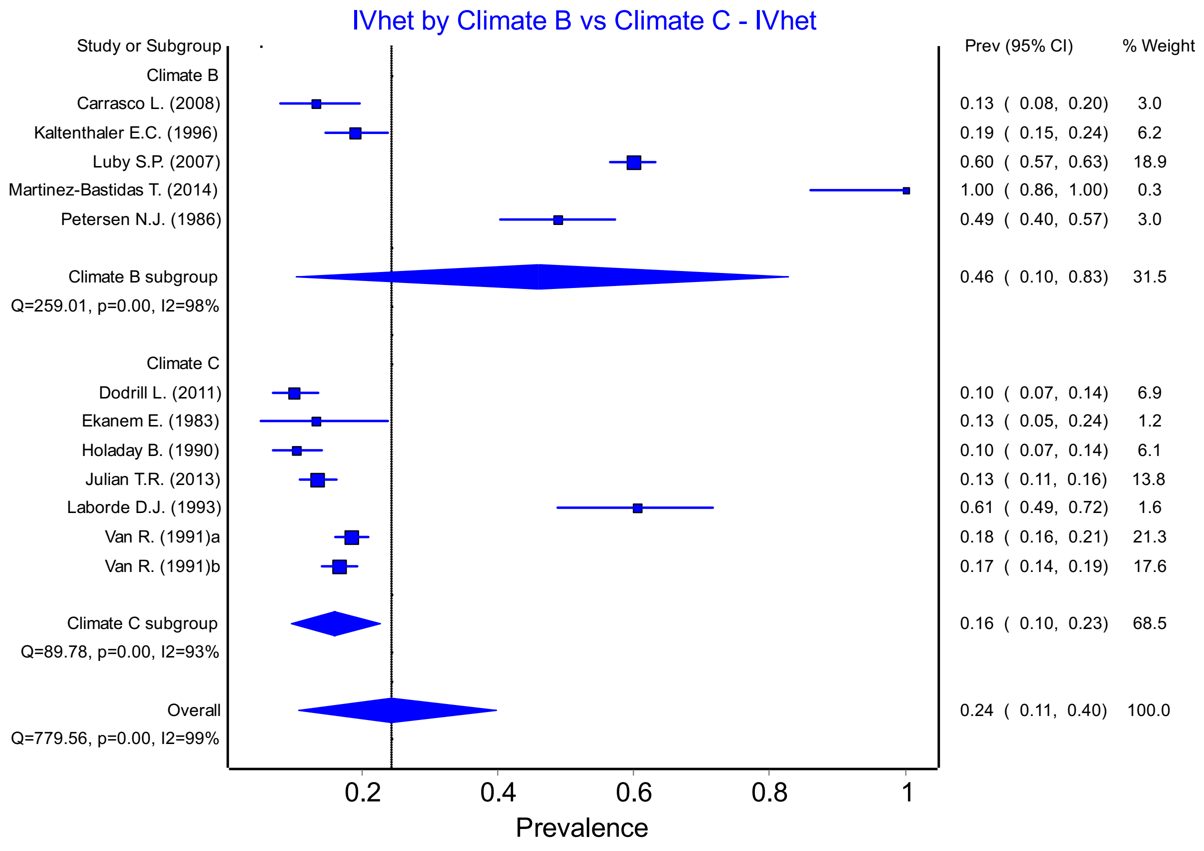


Figure S25: Fecal Coliform Prevalence Climate Group B vs Group C

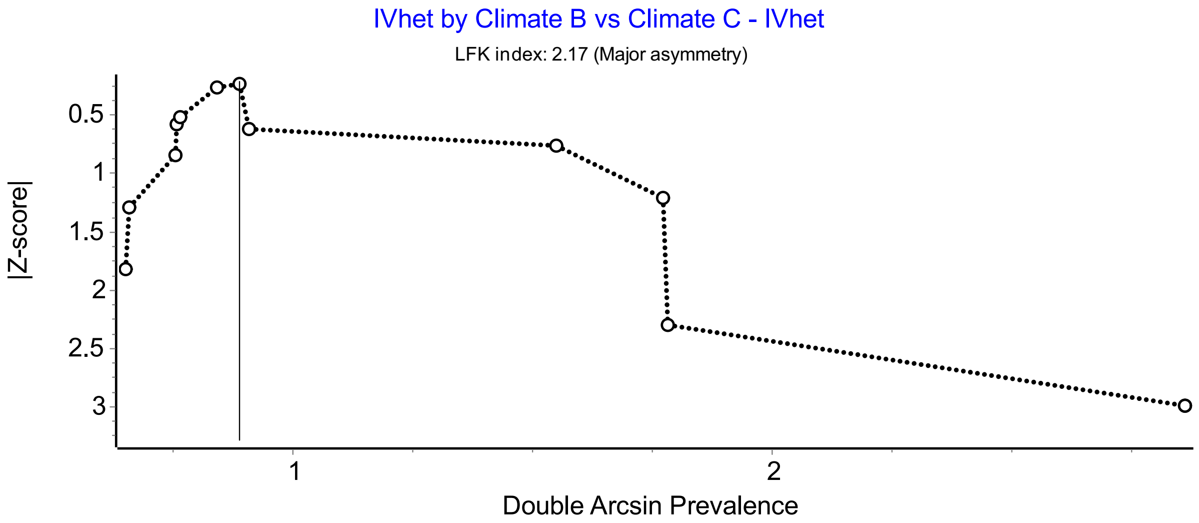


Figure S26: Doi plot for Fecal Coliform Prevalence Climate Group B vs Group C

### Adults vs. Children

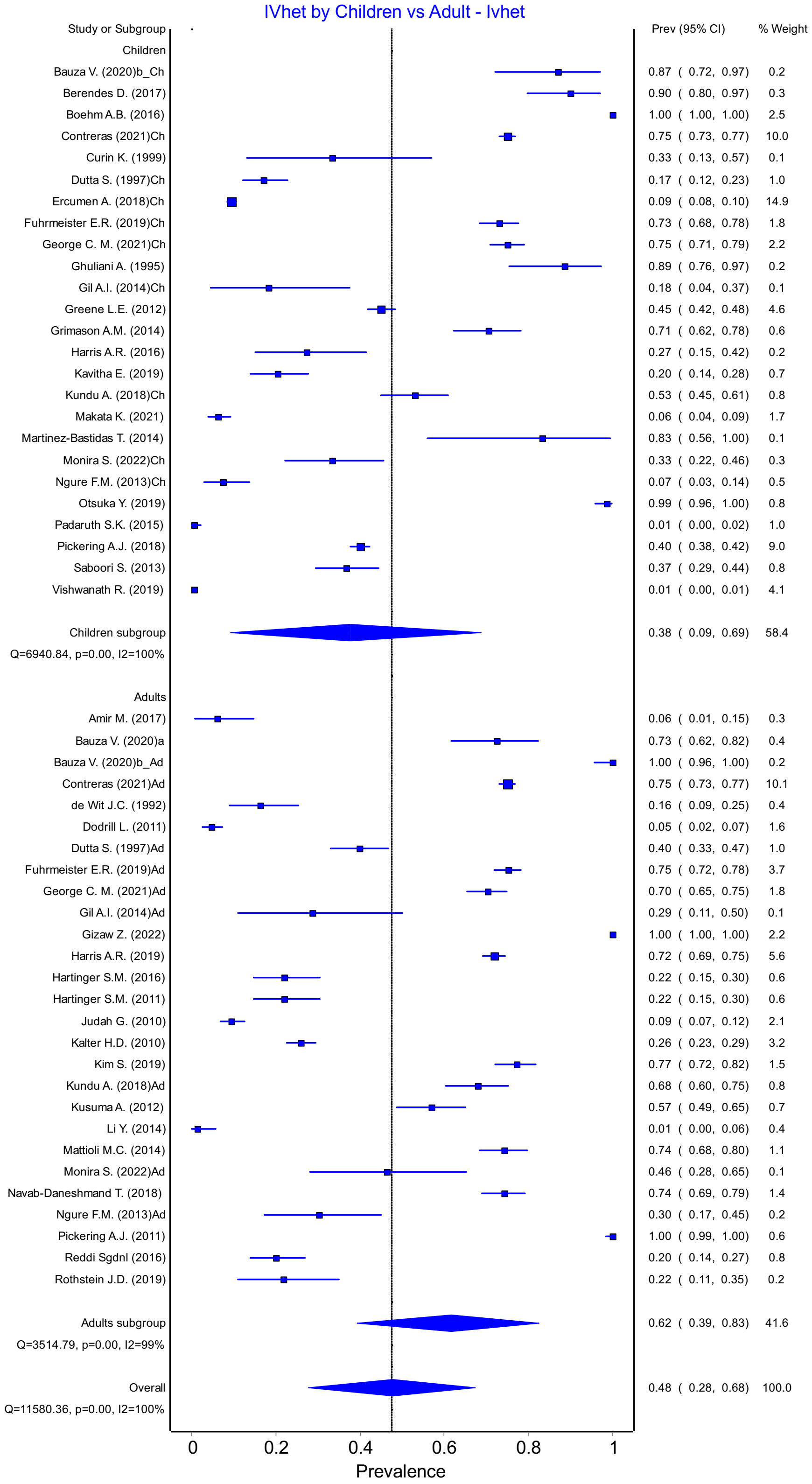


Figure S27: *E. Coli* Prevalence for Adults vs. Children

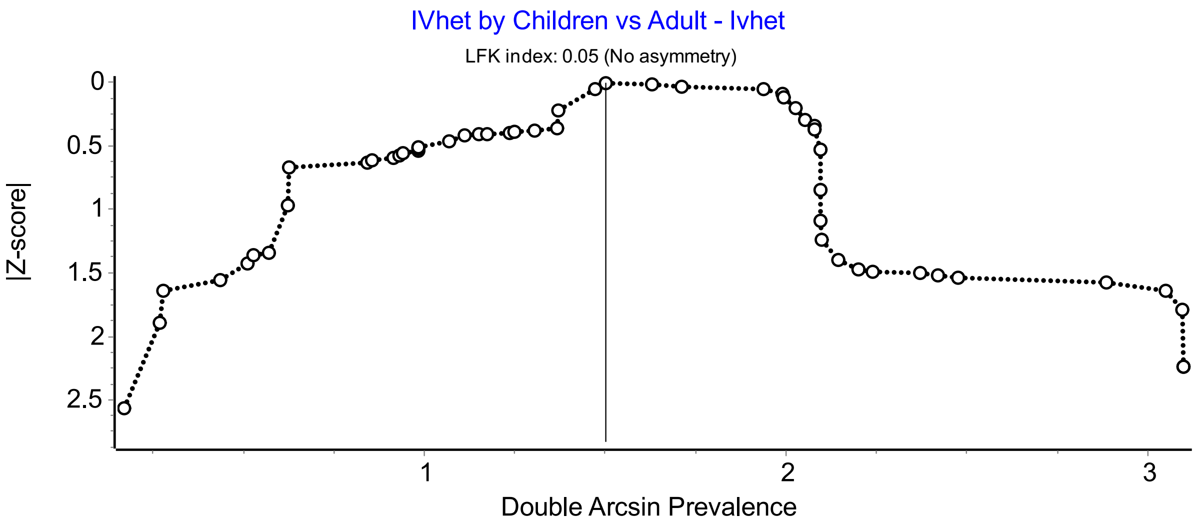


S28: Doi plot of *E. Coli* Prevalence for Adults vs. Children

The adults vs. children subgroup comparison of *E. Coli* prevalence was also broken into country income groups. There were not enough studies in upper-middle/high income countries that reported *E. Coli* prevalence for analysis to be completed in this group.

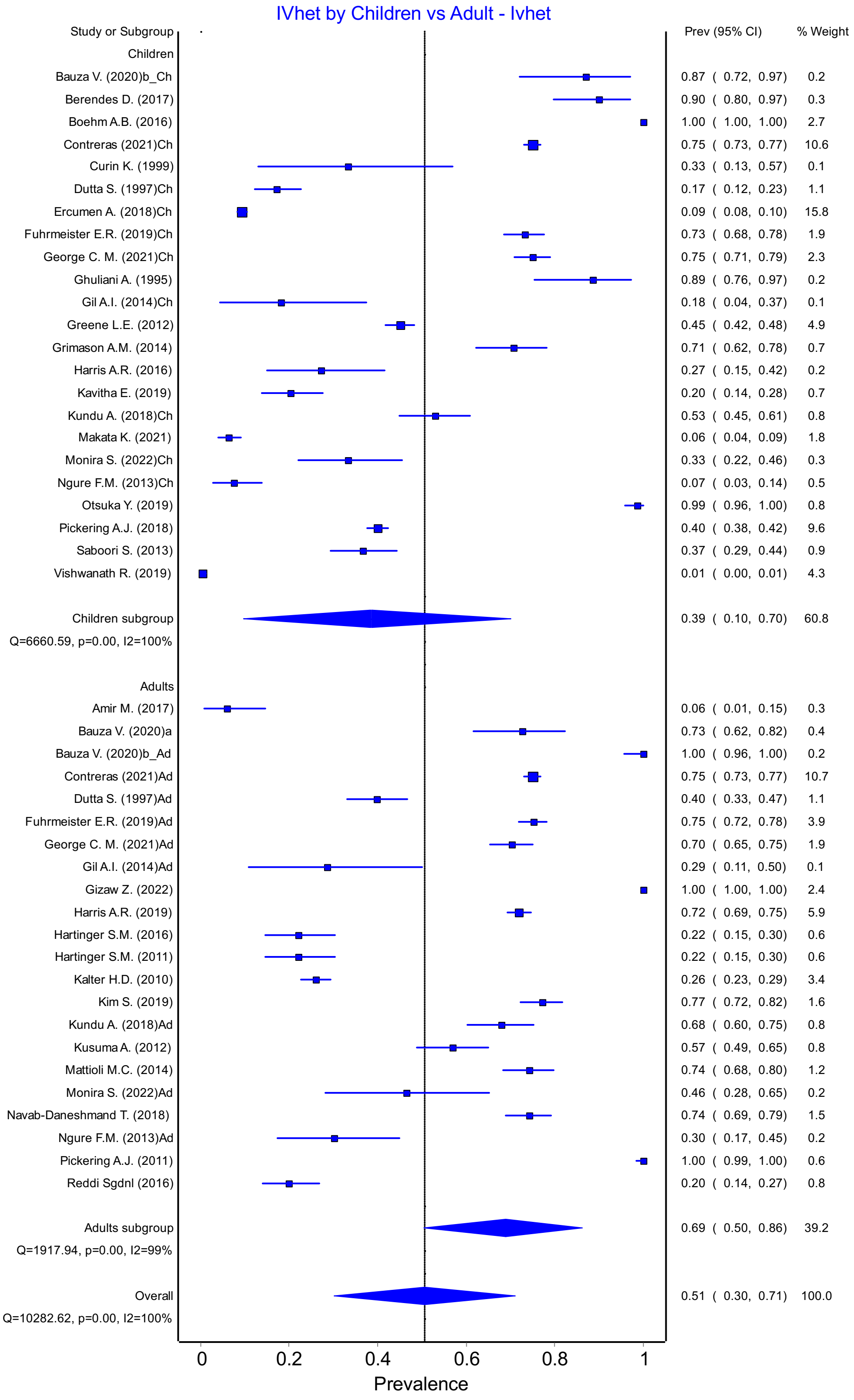

Figure S29: *E. Coli* Prevalence Adults vs. Children in low/lower-middle income countries

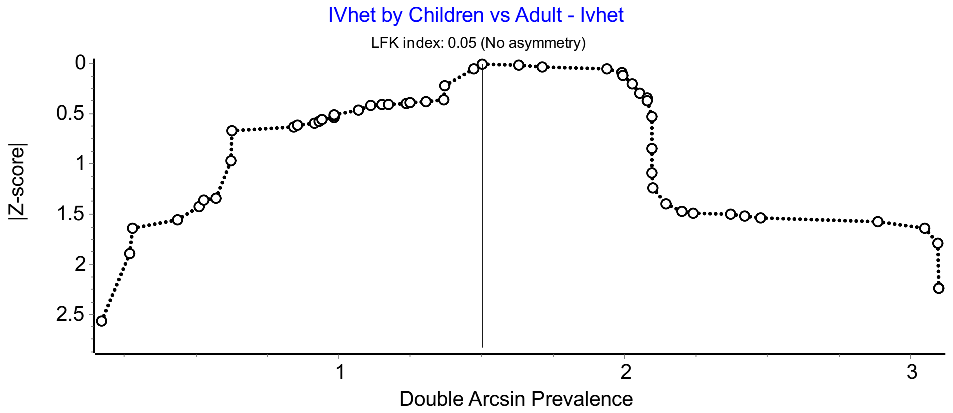

Figure S30: Doi plot of *E. Coli* Prevalence Adults vs. Children in low/lower-middle income countries

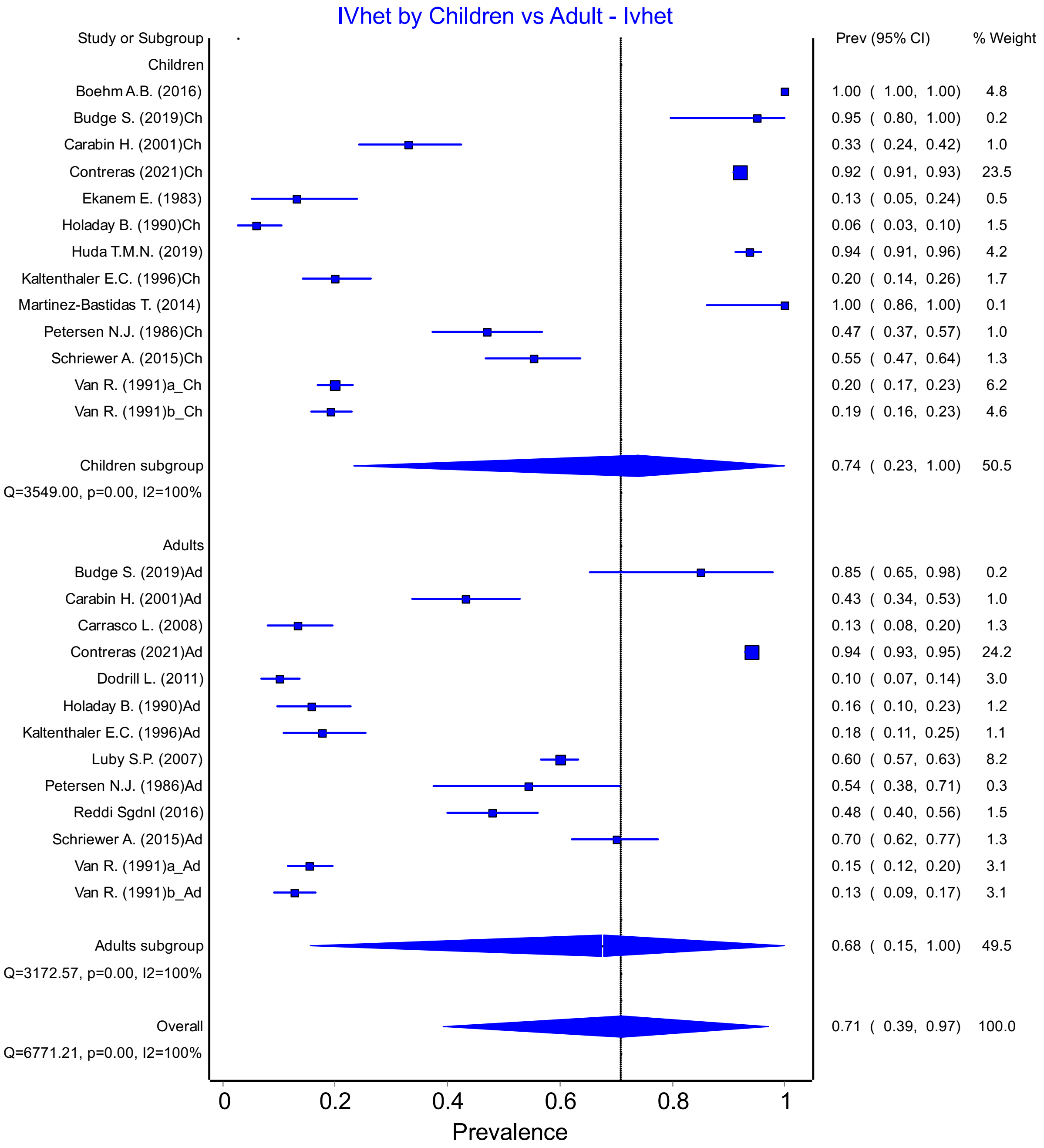


Figure S31: Fecal Coliform Prevalence for Adults vs. Children

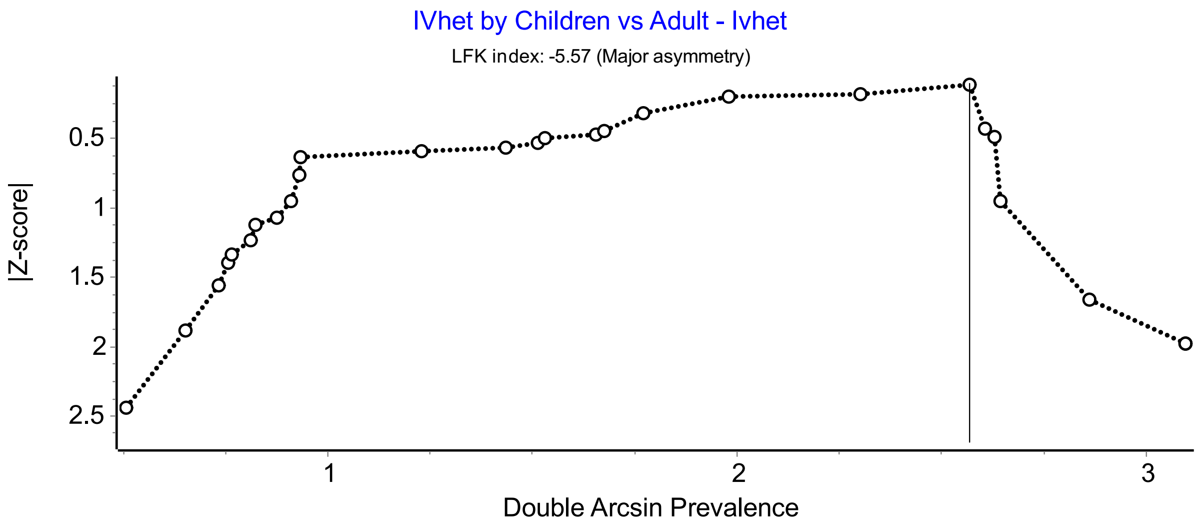


Figure S32: Doi plot of Fecal Coliform Prevalence for Adults vs. Children

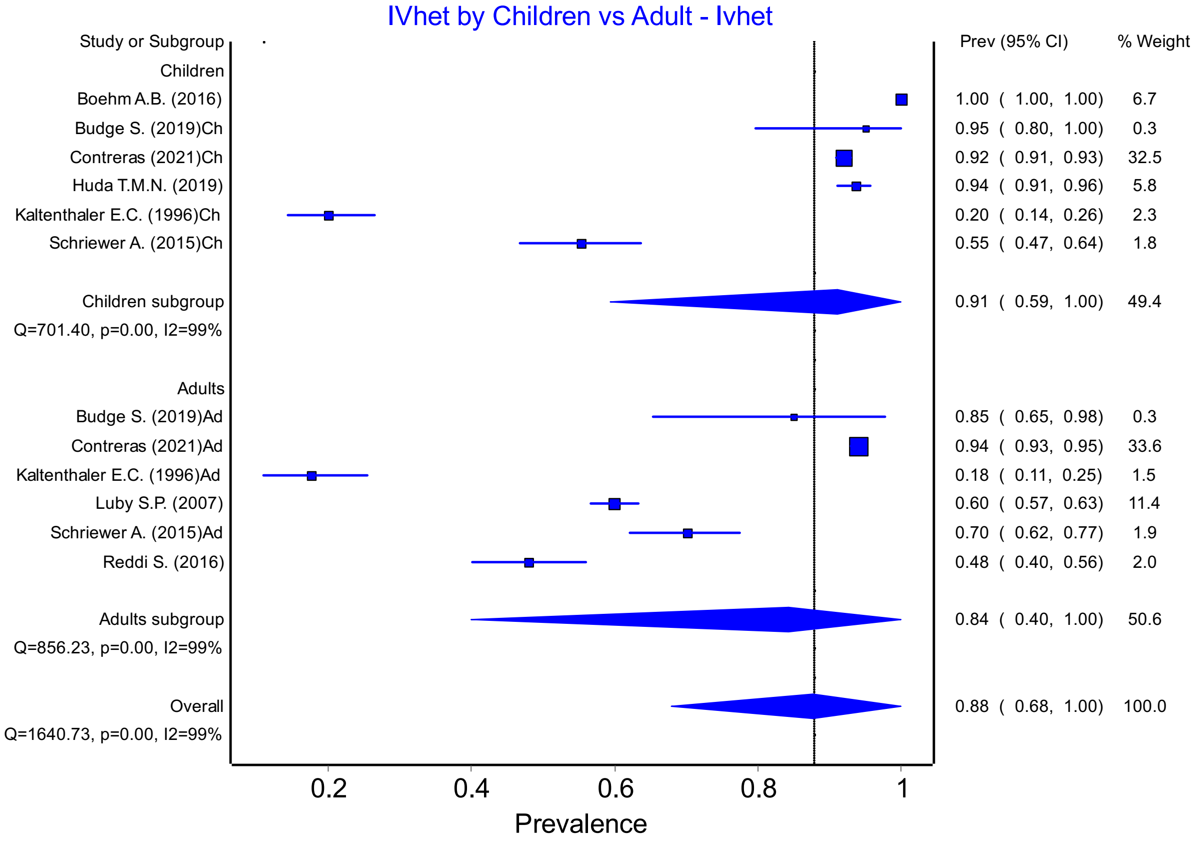


Figure S33: Fecal Coliform Prevalence Adults vs. Children in low/lower-middle income countries

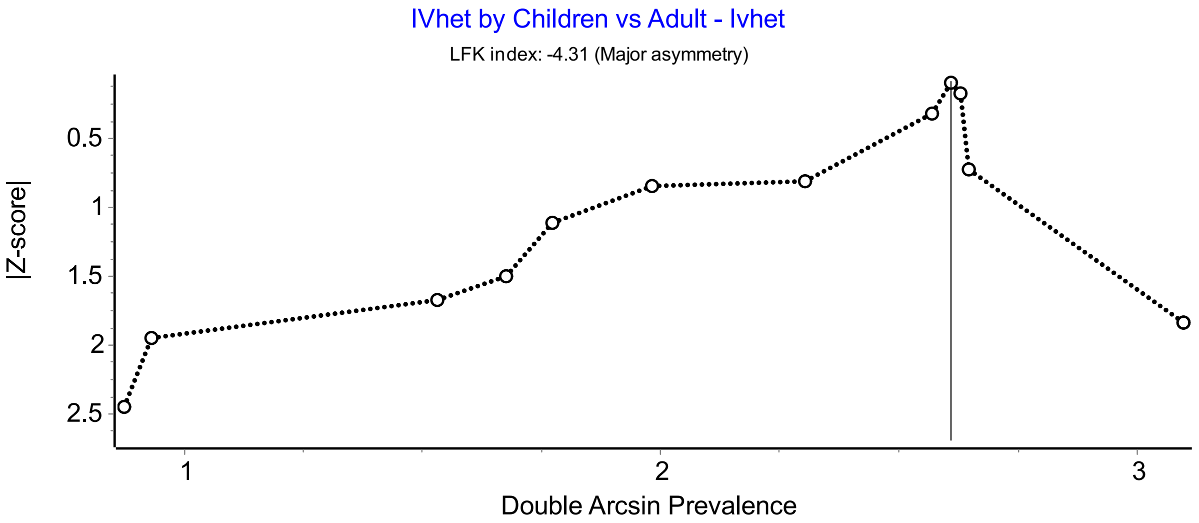


Figure S34: Doi plot of Fecal Coliform Prevalence Adults vs. Children in low/lower-middle income countries

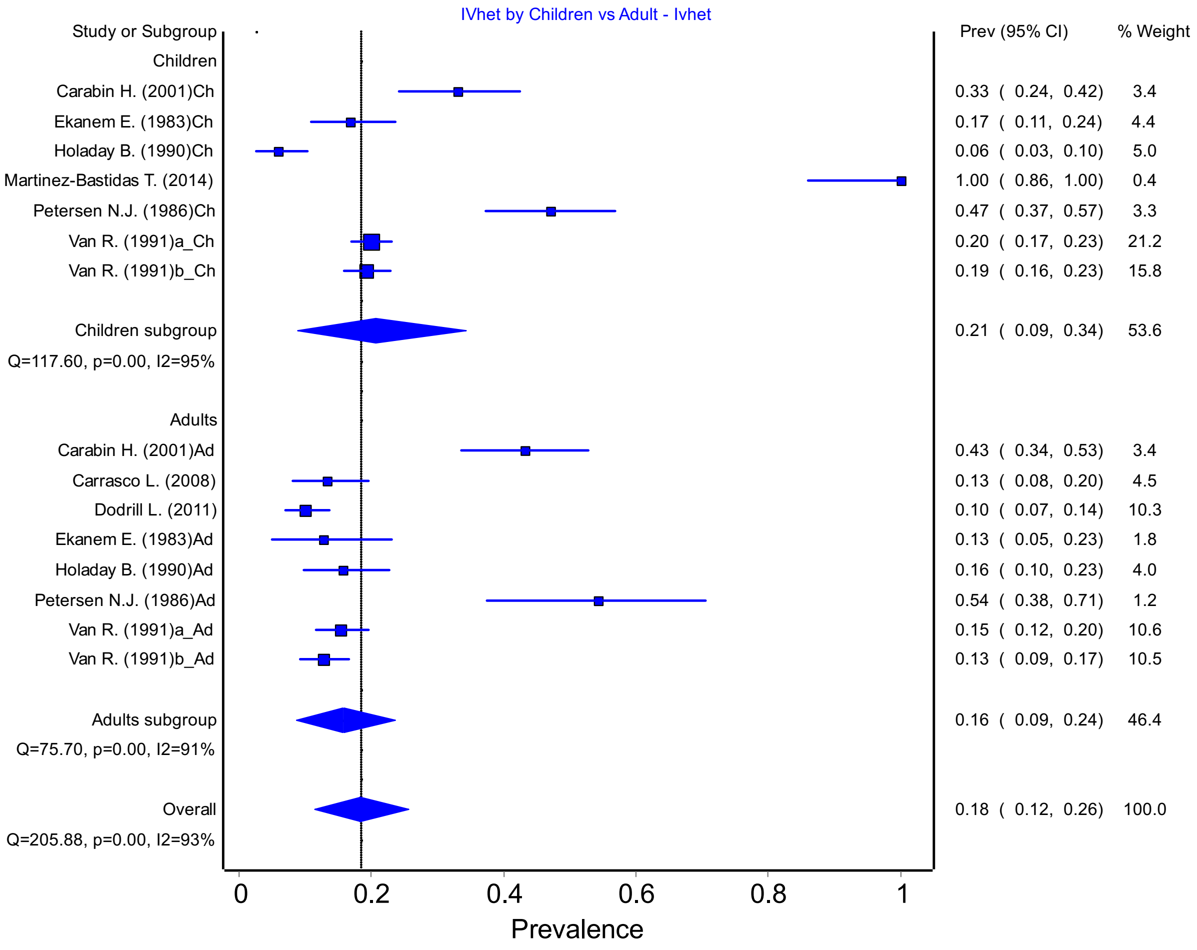


Figure S35: Fecal Coliform Prevalence Adults vs. Children in middle-high/high income countries

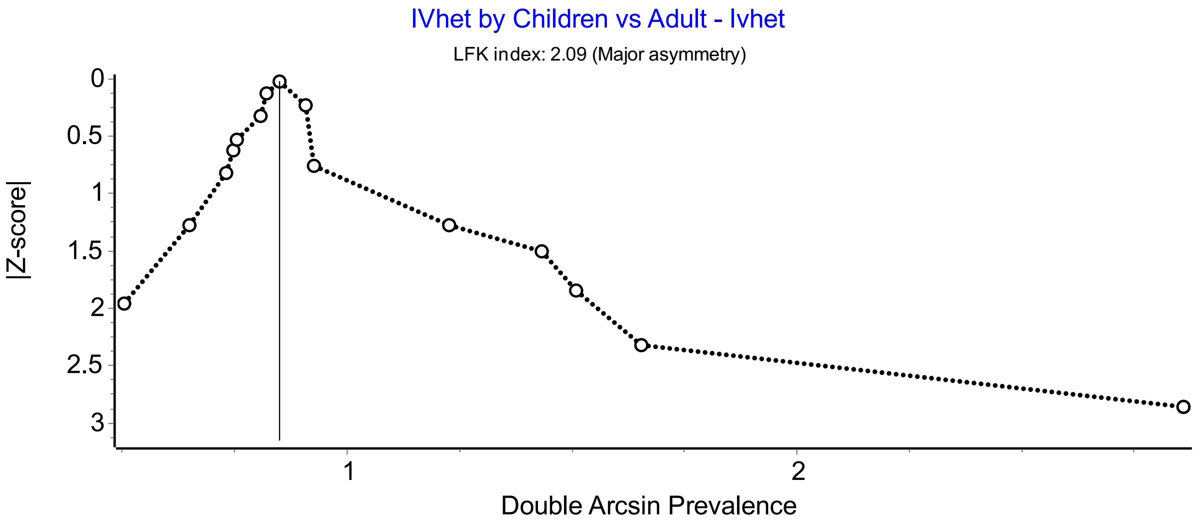


Figure S36: Doi plot of Fecal Coliform Prevalence Adults vs. Children in middle-high/high income countries

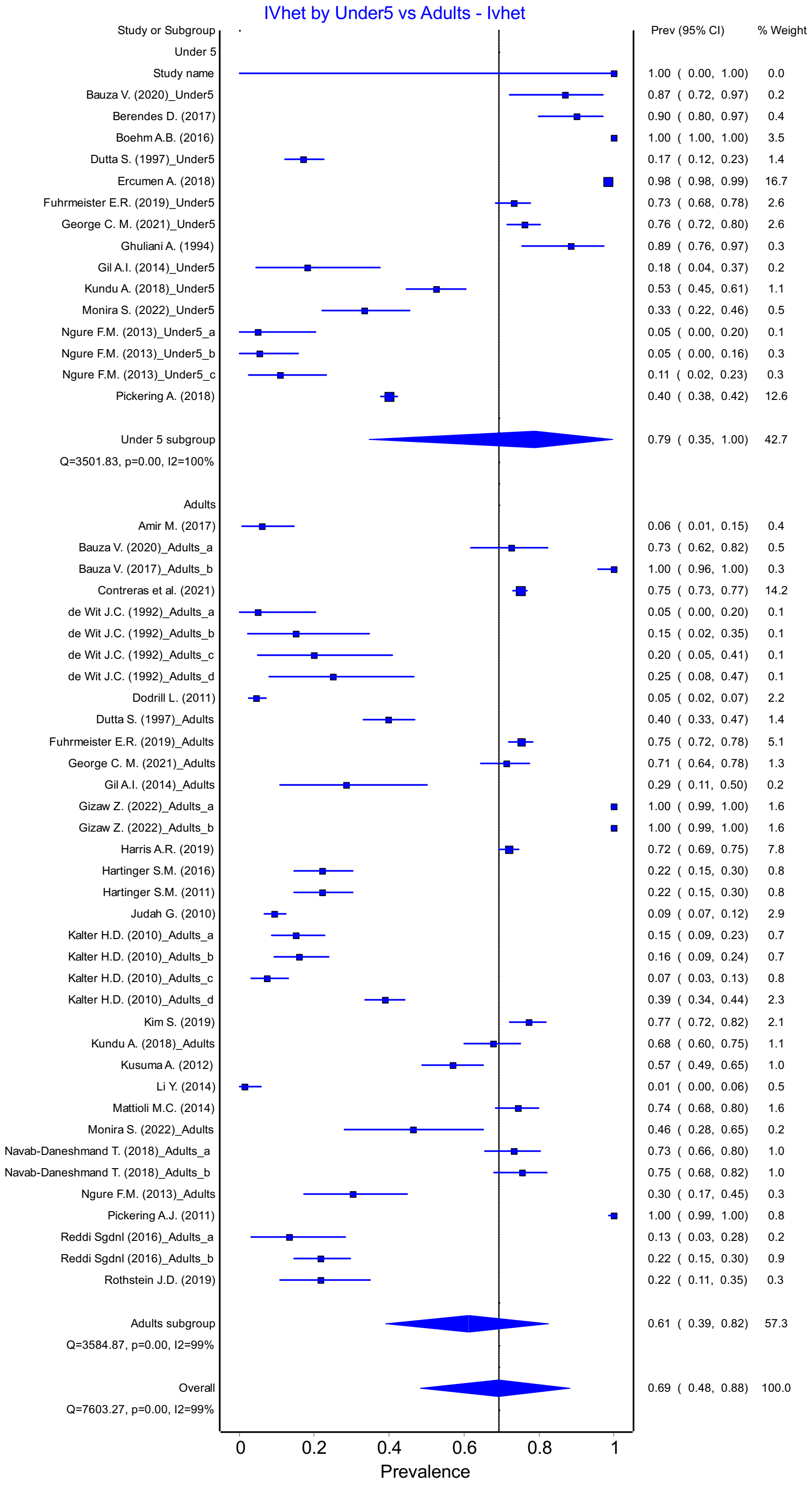


Figure S37: *E. Coli* Prevalence Adults vs. Children Under 5

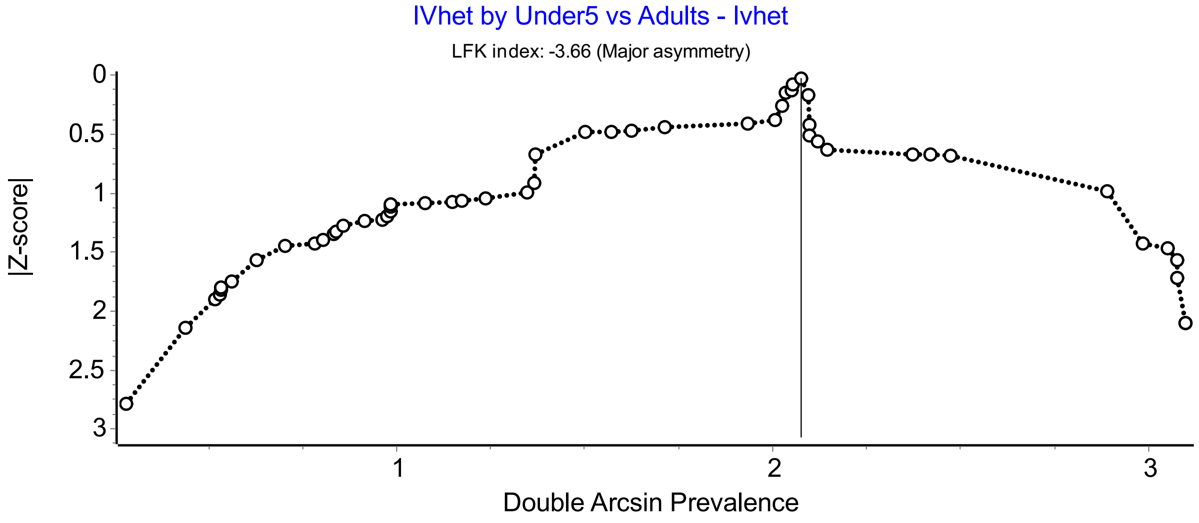


Figure S38: Doi plot of *E. Coli* Prevalence Adults vs. Children Under 5

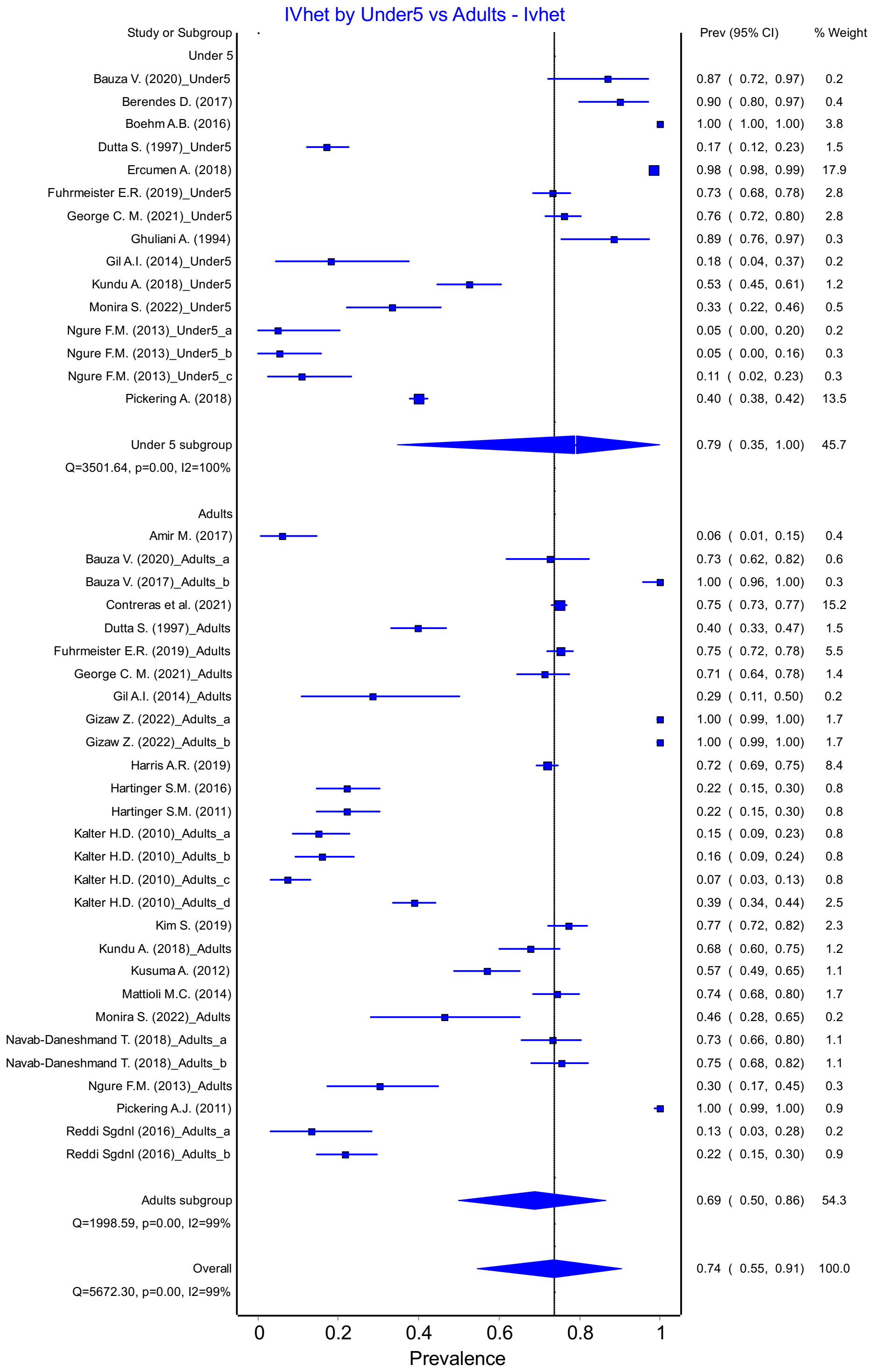


Figure S39: *E. Coli* Prevalence Adults vs. Children Under 5 in low/lower-middle income countries

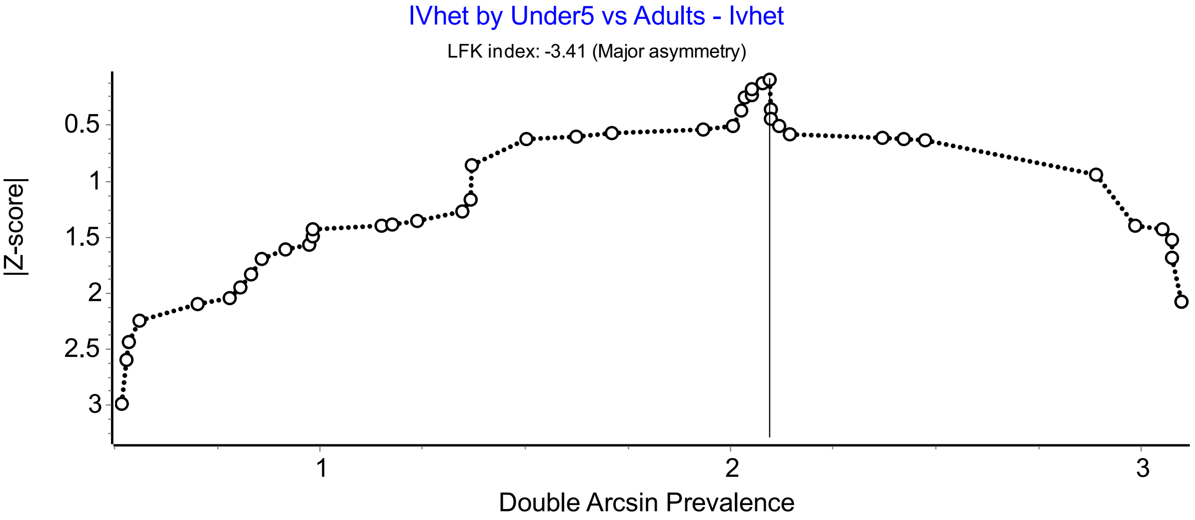


Figure S40: Doi plot of *E. Coli* Prevalence Adults vs. Children Under 5 in low/lower-middle income countries

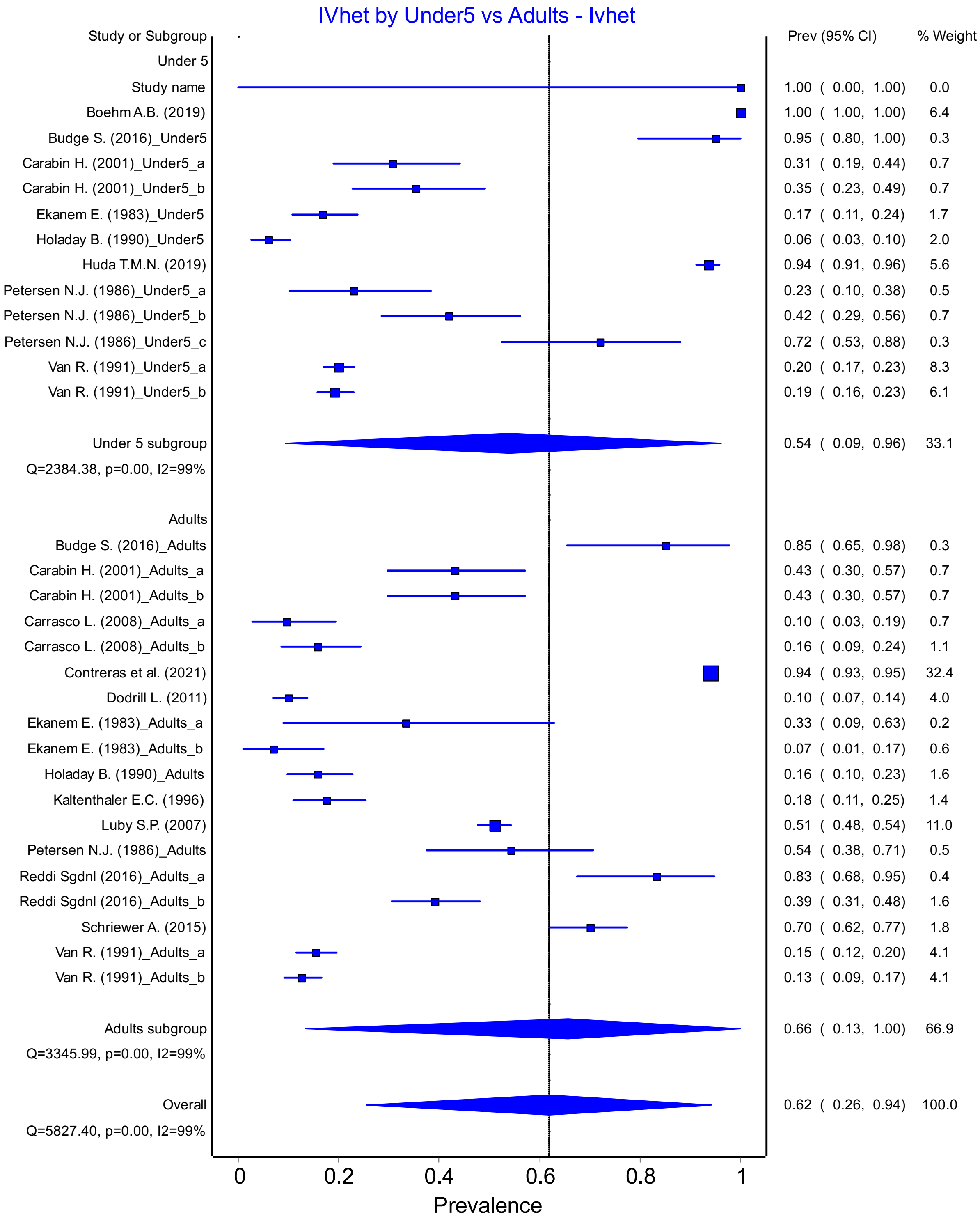


Figure S41: Fecal Coliform Prevalence Adults vs. Children Under 5

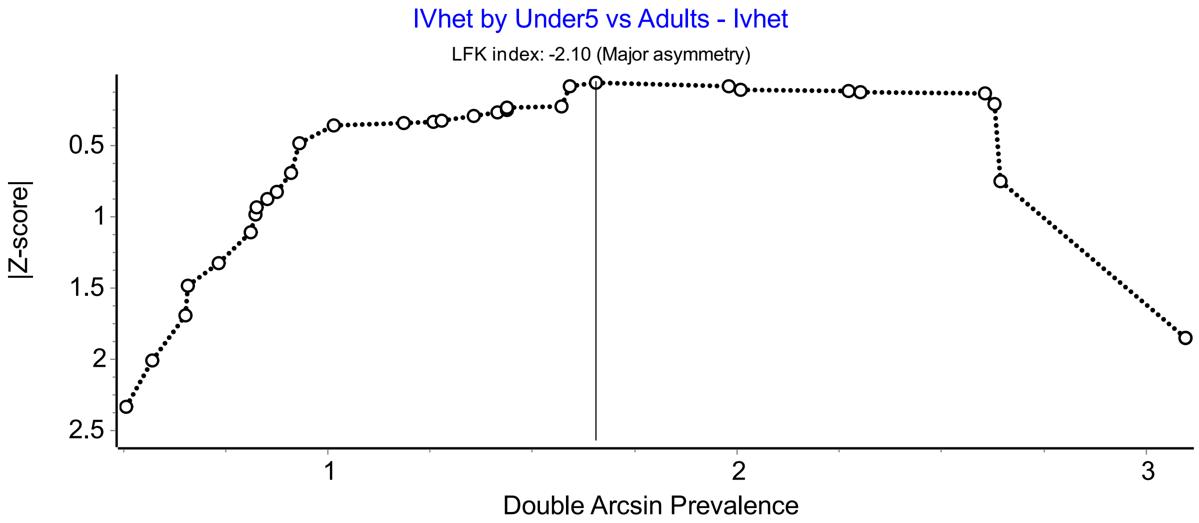


Figure S42: Doi plot of Fecal Coliform Prevalence Adults vs. Children Under 5

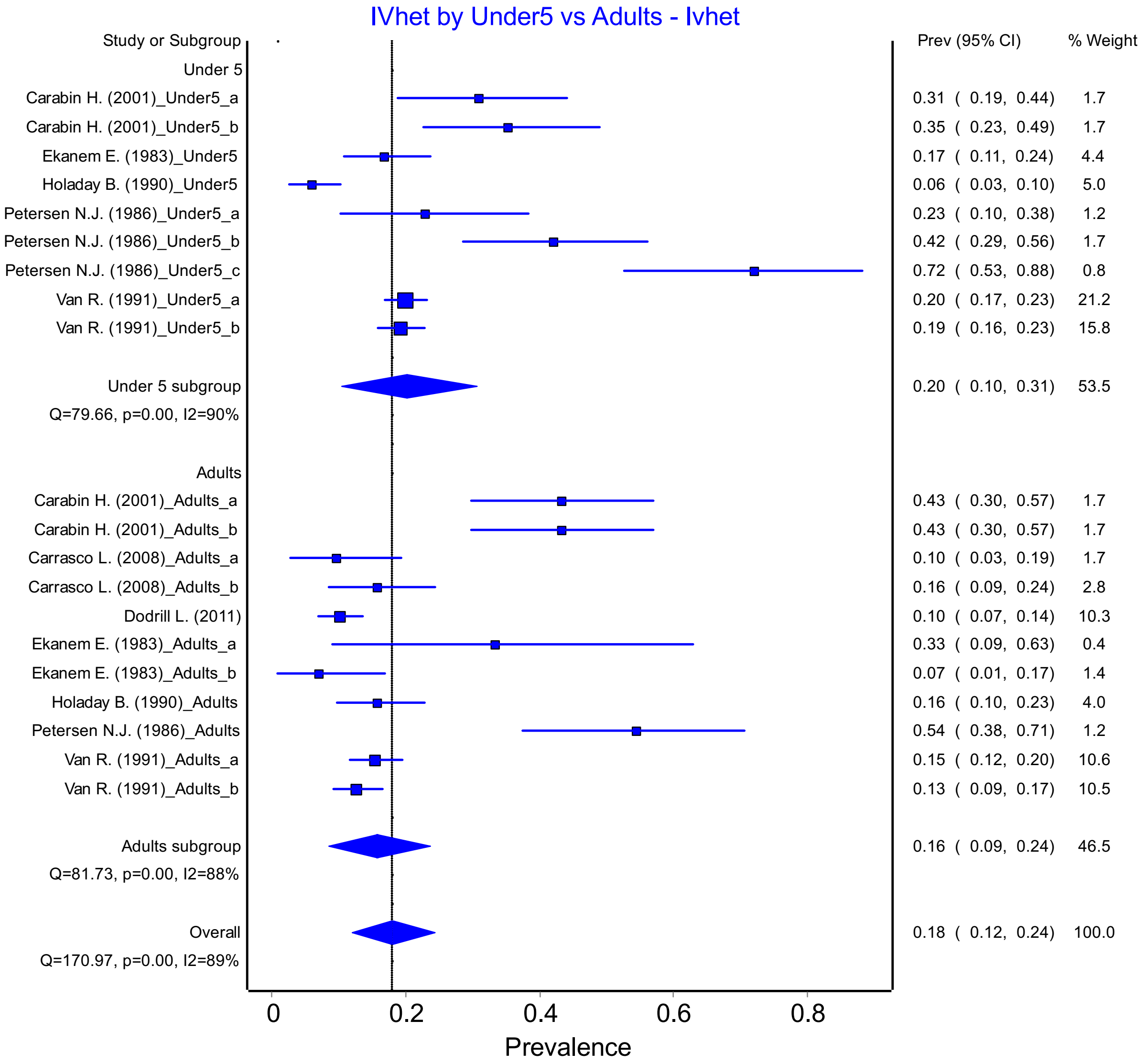


Figure S43: Fecal Coliform Prevalence Adults vs. Children Under 5 in upper-middle/high income countries

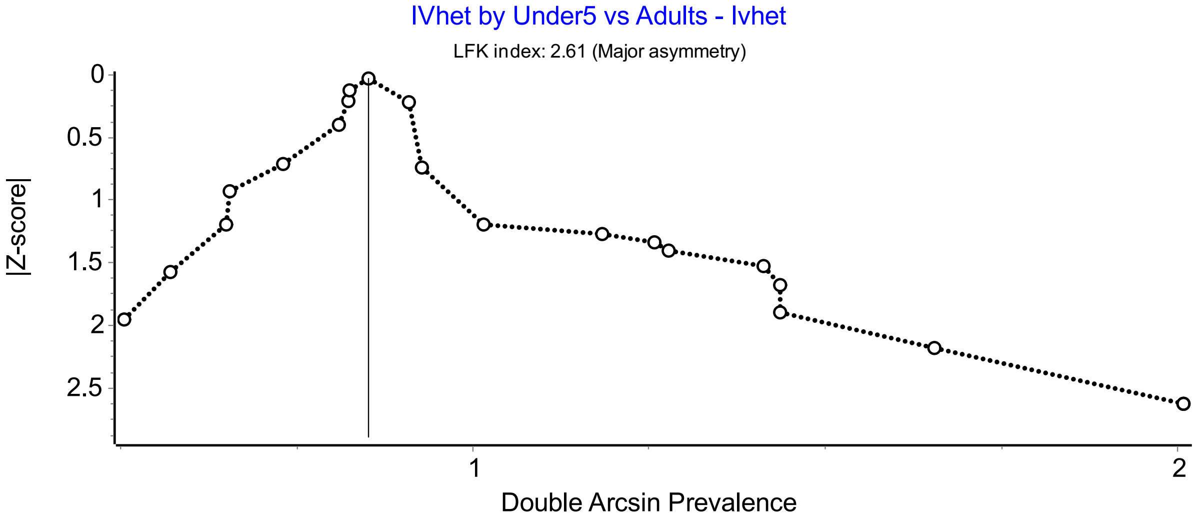


Figure S44: Doi plot of Fecal Coliform Prevalence Adults vs. Children Under 5 in upper-middle/high income countries

**Methods**

Only one study that reported *E. Coli* used the impression method, so we could only do analysis on comparing rinse and swab methods for *E. Coli* prevalence.

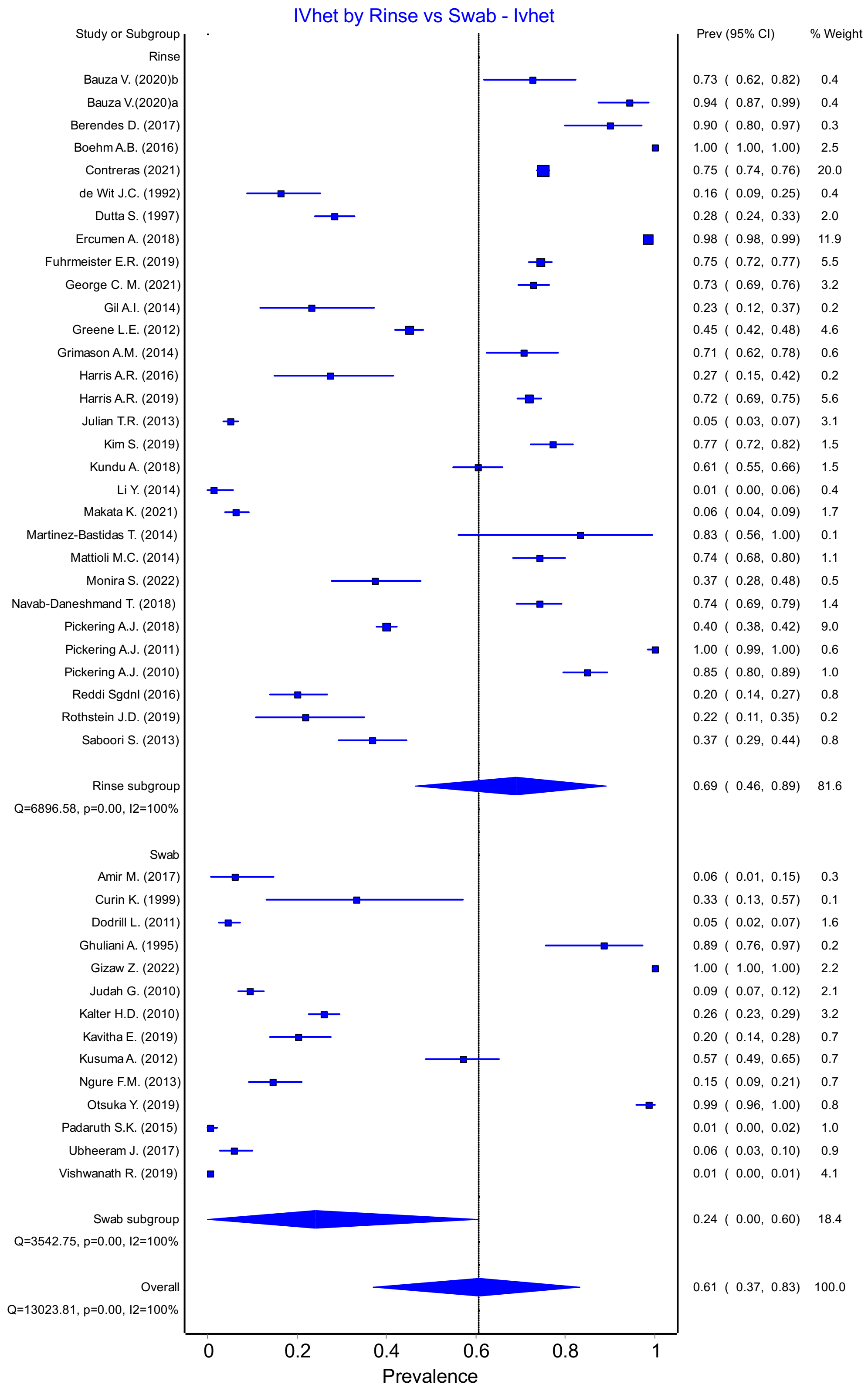


Figure S45: *E. Coli* prevalence in rinse vs swab methods

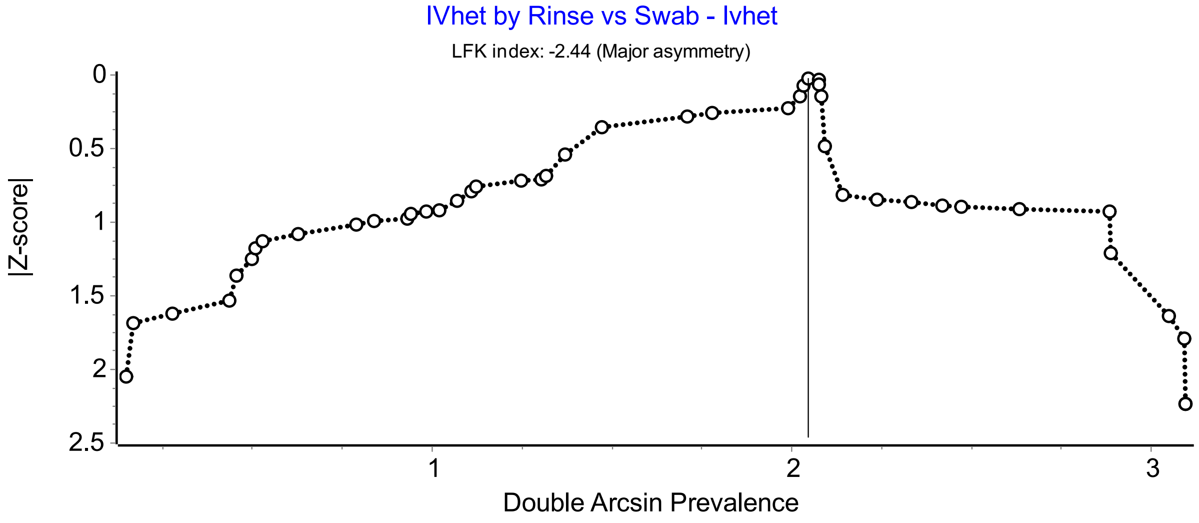


Figure S46: Doi plot of *E. Coli* prevalence in rinse vs swab methods

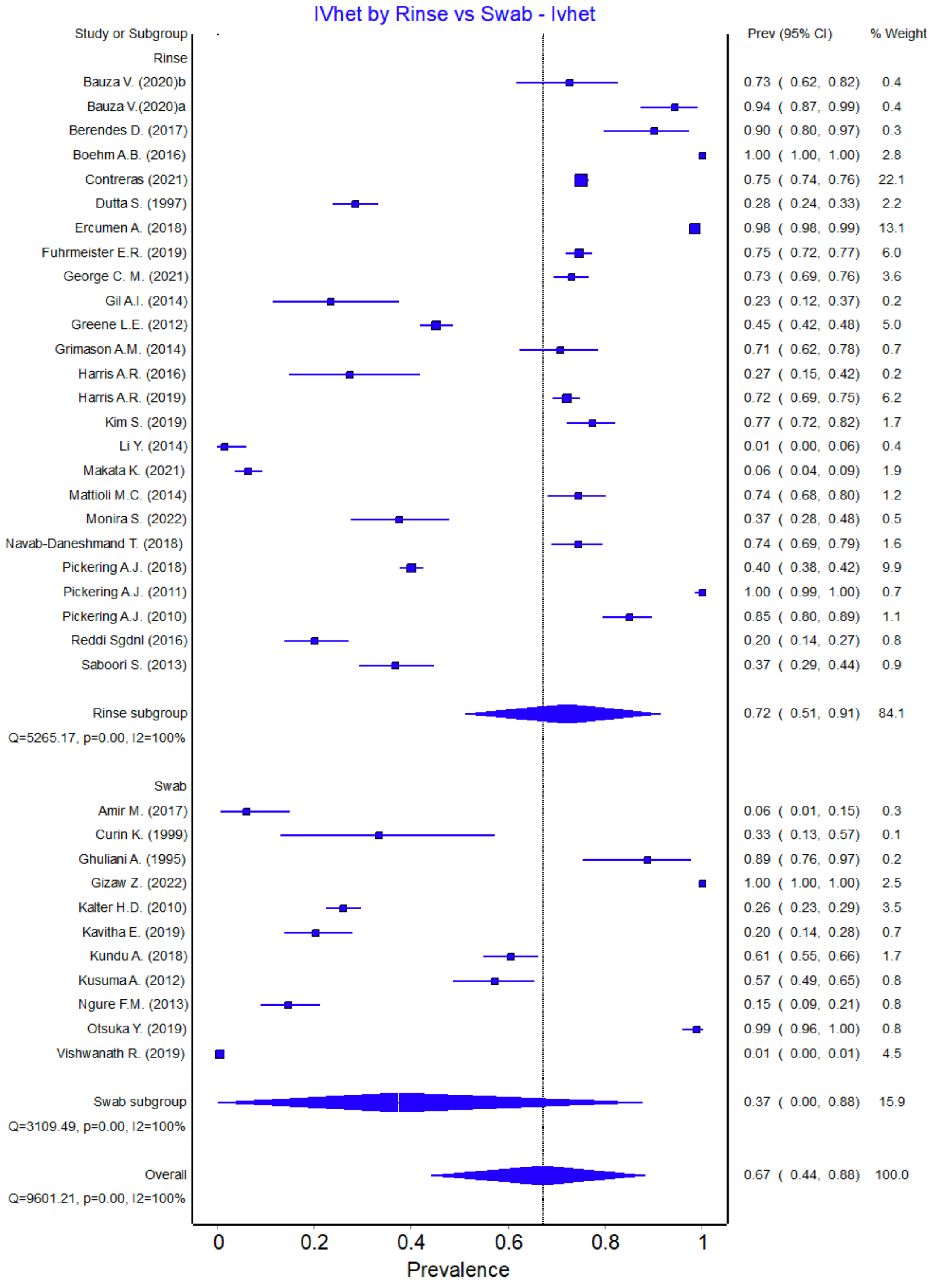


Figure S47: *E. Coli* prevalence in rinse vs swab methods in low/lower-middle income countries methods

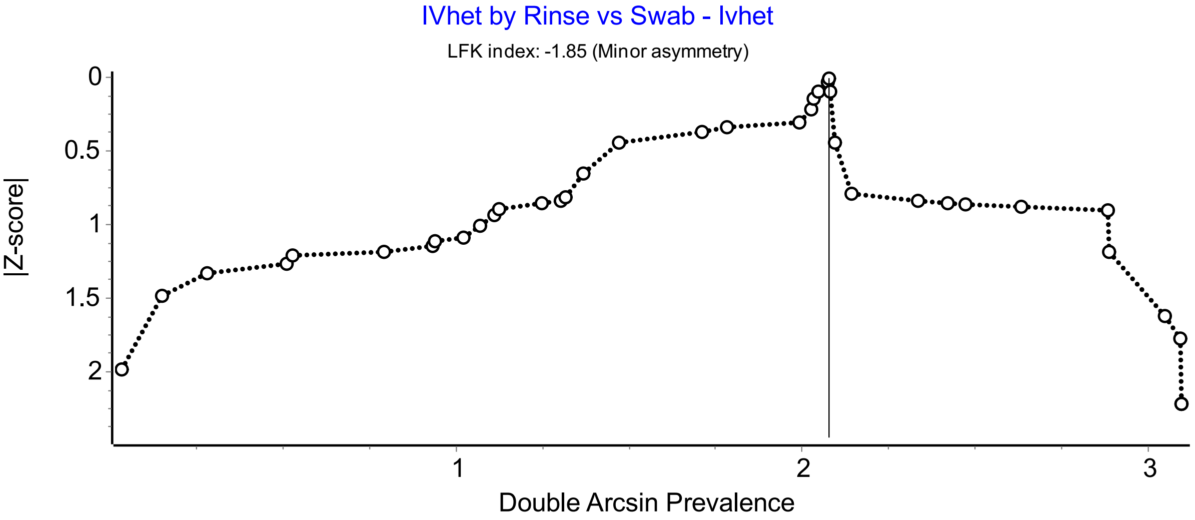


Figure S48: Doi plot of *E. Coli* prevalence in rinse vs swab methods in low/lower-middle income countries

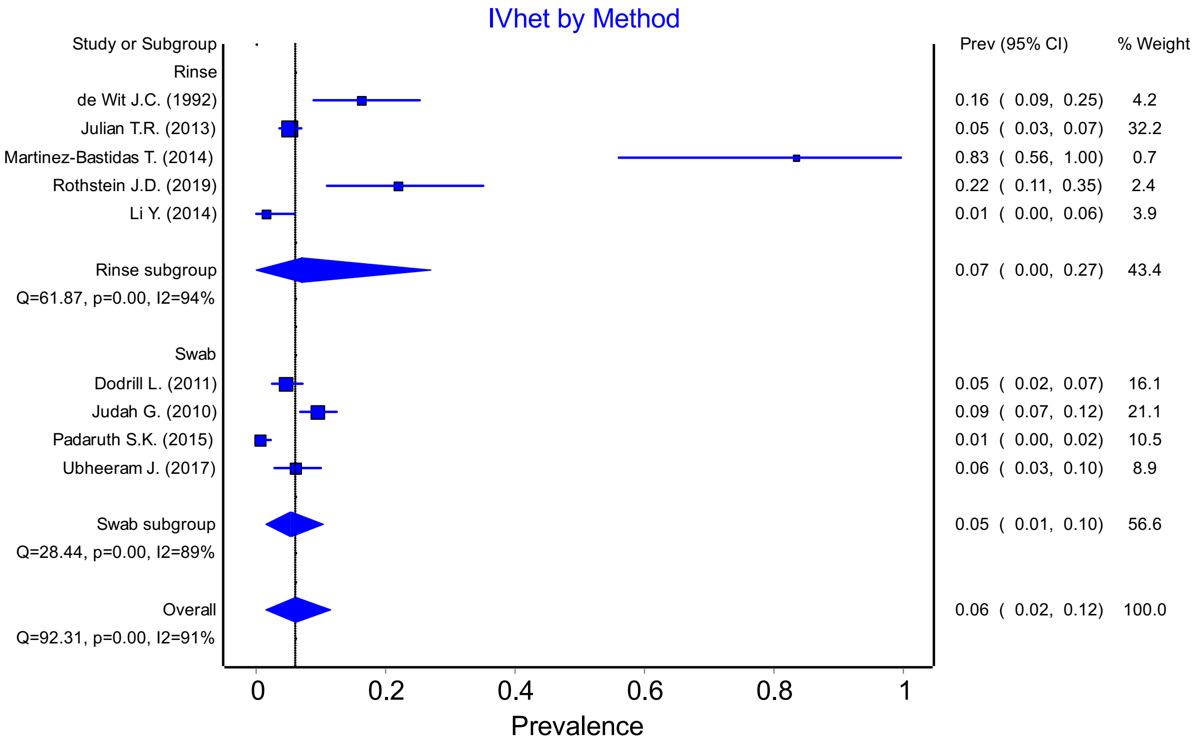


Figure S49: *E. Coli* prevalence in rinse vs swab methods in high/upper-middle income countries

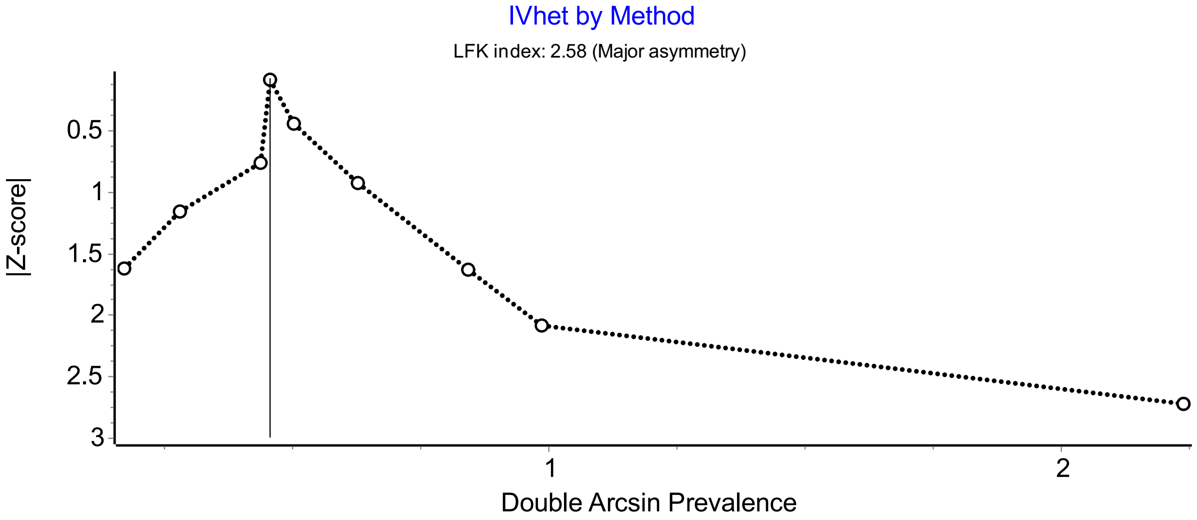


Figure S50: Doi plot of *E. Coli* prevalence in rinse vs swab methods in high/upper-middle income countries

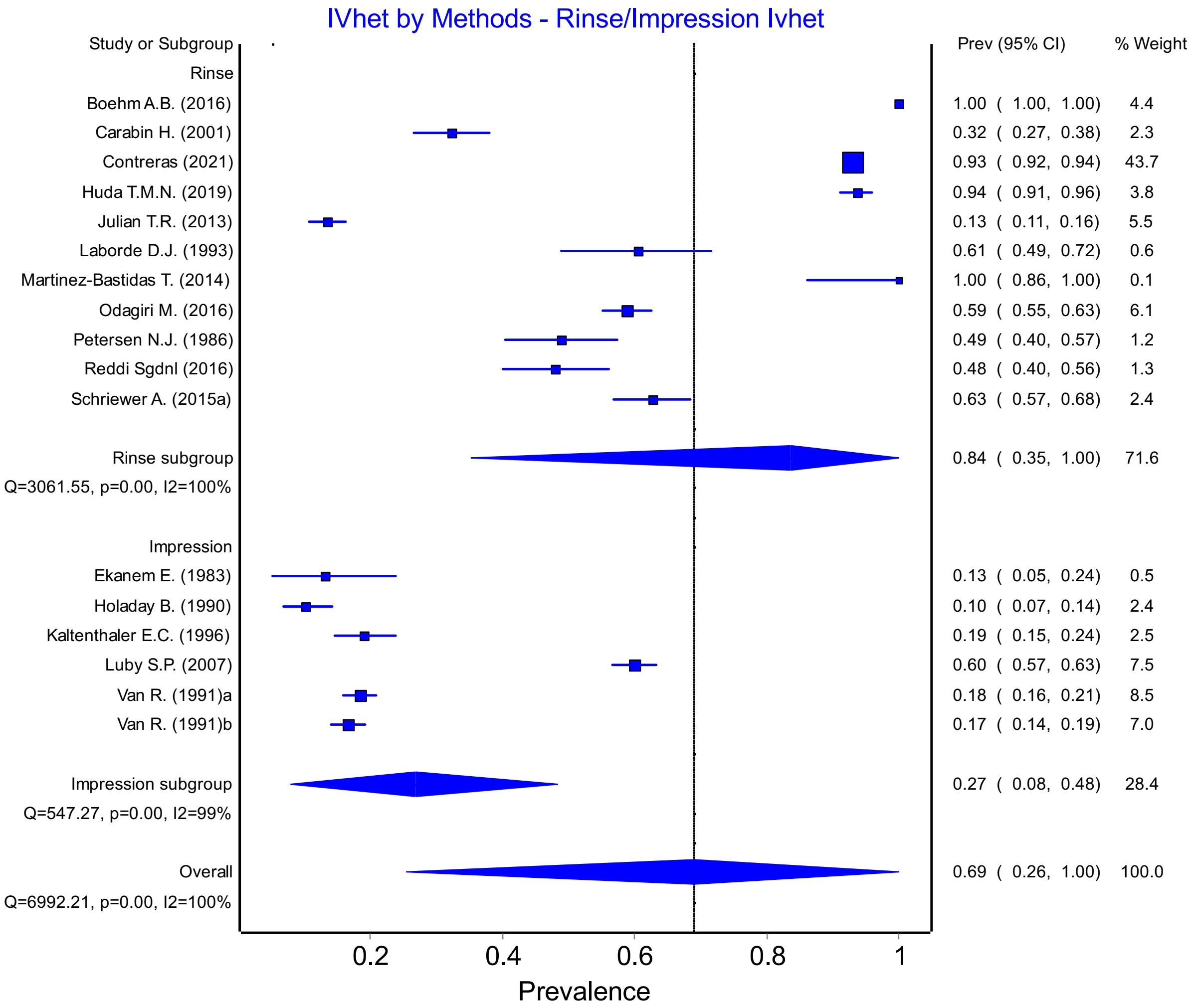


Figure S51: Fecal Coliform prevalence in rinse vs impression methods

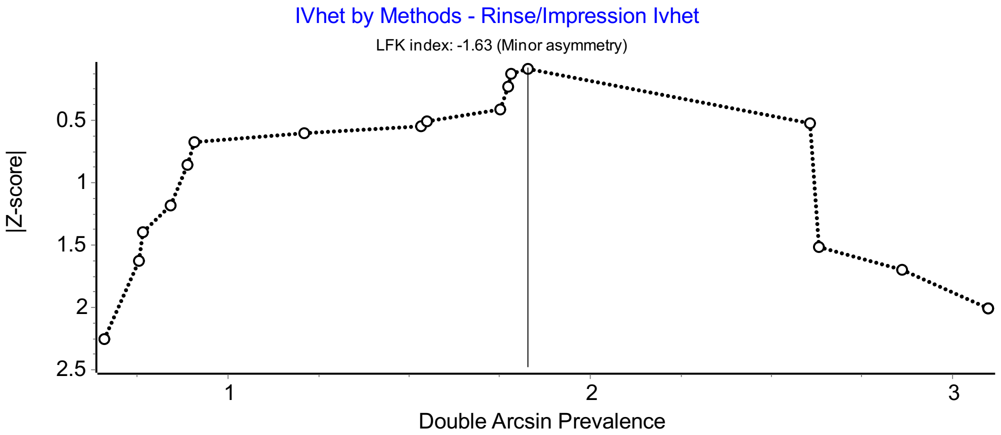


Figure S52: Doi plot of Fecal Coliform Prevalence in rinse vs impression methods

PRISMA-P Checklist for Environmental Contamination

Title: **Hands are frequently contaminated with fecal bacteria and enteric pathogens globally: A systematic review and meta-analysis**

Research team: **Tim Julian (Eawag), Amy Pickering (Stanford/Tufts/UC Berkeley)**

**Co-Authors:** Lou Curchod (Hands Term Paper), Rahel Scheidegger (Review Articles), Molly Cantrell (Review Articles), Émile Sylvestre (Data Analysis), Hannah Wharton (Review Articles)

Systematic Review (PRISMA-P 2015 - checklist)

RATIONALE:

**Enteric pathogen transmission occurs through contamination of, and subsequent exposure to, environmental reservoirs. The environmental reservoirs demonstrated to be of concern include food, water, flies, surfaces (or fomites), hands, and fields (or soils). Enteric pathogens may also be spread as aerosols, though there is limited data on infectivity of airborne exposures.**

**Despite awareness of the importance of multiple potential reservoirs, previous research focuses on transmission of enteric pathogens through water and food. This review aims to compile evidence on enteric pathogen transmission through the understudied environmental pathways of hands. The review will characterize contamination on hands, provide insight on child exposure, and highlight which pathways should be targeted for intervention.**

**The review is targeted to two audiences: practitioners and researchers. By highlighting the potential importance of neglected pathways, the review could spur interest in understudied or underappreciated interventions.**

OBJECTIVE:

**To describe the prevalence and level of contamination with fecal indicator bacteria and enteric pathogens in understudied environmental transmission pathway of hands.**

**Secondary objectives are to compare contamination levels between: 1) developed and developing countries, 2) children and adults, and 3) quantification/detection methods.**

Methods

ELIGIBILITY CRITERIA:
*Specify the study characteristics (such as PICO, study design, setting, time frame) and report characteristics (such as years considered, language, publication status) to be used as criteria for eligibility for the review*

**We will include peer reviewed, published study designs that measured fecal indicators in non-occupational settings, such as households and communities, including both developed and developing countries.**

INFORMATION SOURCES:

*Describe all intended information sources (such as electronic databases, contact with study authors, trial registers or other grey literature sources) with planned dates of coverage*

**All data will come from peer reviewed literature published before November 2022.**

SEARCH STRATEGY:
*Present draft of search strategy to be used for at least one electronic database, including planned limits, such that it could be repeated*

**We will search the following databases: PubMed, EMBASE, Web of Science**

**We will use the following generic search string, adapted for the databases listed above: (((fecal OR pathogenic OR enteric ) AND bacteria) OR e. coli OR enterococci OR helminth OR protozoa OR virus OR phage) AND hand AND contamination**[2]

DATA MANAGEMENT:
*Describe the mechanism(s) that will be used to manage records and data throughout the review*

**Databases will be searched directly and records downloaded into Endnote. Endnote software will be used first to remove duplicates, and then to create a master file for importing into Covidence (www.covidence.com www.covidence.org). Covidence is commercial software providing a collaborative environment for screening and managing records.**

SELECTION PROCESS:
*State the process that will be used for selecting studies (such as two independent reviewers) through each phase of the review (that is, screening, eligibility and inclusion in meta-analysis)*

**Titles and abstracts are initially screened to exclude irrelevant studies. Studies are excluded if they: 1) are about microorganisms that are not enteric pathogens or fecal indicators, 2) do not present primary data (e.g. reviews), or 3) suggest sampling of only food or water.**

**Only studies dealing with natural hand contamination are eligible, studies in hospitals, laboratories, or using artificial hand contamination were excluded. Also, studies dealing with food and animal contamination were excluded, even if they included work data from food handler's hands.**

**Two independent reviewers screen records using titles and abstracts.**

DATA COLLECTION PROCESS:
*Describe planned method of extracting data from reports (such as piloting forms, done independently, in duplicate), any processes for obtaining and confirming data from investigators*

**A standardized form will be used to extract data. Two or more reviewers will extract data from 3 studies to pilot the standardized form. Reviewers will continue to extract data with 10% record duplication (verified periodically) to ensure consistency.**

DATA ITEMS:
*List and define all variables for which data will be sought (such as PICO items, funding sources), any pre-planned data assumptions and simplifications*

**Specific data from each study we are interested in, for every combination of pathogen and environmental reservoir:**

- **First Author**
- **Year**
- **Study Link (ncbi or PDF)**
- **Microbiological Indicator or Pathogen (e.g. Pathogen or Bacteria)**
- **Target Population (i.e., Children, Female Caretakers, Heads of Households, Farm Workers)**
- **Adults vs. Children**
- **Country**
- **Country Income Level (High, Medium, Low)**
- **City/Locale**
- **Urban v. Rural**
- **Number of Samples**
- **Number of Samples Positive (above limit of detection)**
- **Concentration (Mean)**
- **Does Concentration include Non-Positives?**
- **Standard Deviation**
- **Concentration (Log10 Mean)**
- **Standard Deviation (Log10 Mean)**
- **Sample Collection Method Described (Details on Method of Collection)**
- **Sample Collection Method Simplified (Impression, Rinse, Swab, Other, Unknown)**
- **Sample Analysis Method (assay used)**
- **Limit of Detection (if reported)**
- **Additional Sample Descriptions (Open)**
- **Study Conclusion or Main Findings of Note**

OUTCOMES AND PRIORITIZATION:
*List and define all outcomes for which data will be sought, including prioritization of main and additional outcomes,with rationale*

**Presence or quantity of fecal indicator bacteria and enteric pathogens (including viruses, bacteria, helminths, phage, and protozoa) on hands.**

RISK OF BIAS IN INDIVIDUAL STUDIES:
*Describe anticipated methods for assessing risk of bias of individual studies, including whether this will be done at the outcome or study level, or both; state how this information will be used in data synthesis*

**We will include a discussion in the manuscript of the risk of bias in individual studies related to each indicator.**

DATA SYNTHESIS:

**We will extract the mean prevalence/concentration and standard deviation for each indicator available for hands. We will estimate overall mean prevalence and mean concentration, and analyze the data with respect to study site and sample population.**[3]

META-BIAS:

*Specify any planned assessment of meta-bias(es) (such as publication bias across studies, selective reporting within studies)*

**This is not applicable to our objective considering we are not reviewing an intervention.**

CONFIDENCE IN CUMULATIVE EVIDENCE:
*Describe how the strength of the body of evidence will be assessed (such as GRADE)*

**We will include a qualitative discussion of the strength of the data summarized in the review.**

**Table S3**: Papers included in the systematic review

| | First Author | Year | Title | Countries *(country income classification)* | Population |
|---|---|---|---|---|---|
| 1 | Ahoyo T.A. | 2011 | Impact of water quality and environmental sanitation on the health of schoolchildren in a suburban area of Benin findings in the Savalou-Bante and Dassa-Glazoue sanitary districts (French) | Benin *(low income)* | Children in primary school |
| 2 | Amir M. | 2017 | Impact of unhygienic conditions during slaughtering and processing on spread of antibiotic resistant *Escherichia coli* from poultry | Pakistan *(lower-middle income)* | Consumers before entering a poultry shop |
| 3 | Bauza V. | 2020 | Child feces management practices and fecal contamination: A cross-sectional study in rural Odisha, India | India *(lower-middle income)* | Caregivers |
| 4 | Bauza V. | 2020 | Enteric pathogens from water, hands, surface, soil, drainage ditch, and stream exposure points in low-income neighborhood of Nairobi, Kenya | Kenya *(low income)* | Children and caregivers |
| 5 | Berendes D. | 2017 | The influence of household- and community-level sanitation and fecal sludge management on urban fecal contamination in households and drains and enteric infection in children | India *(lower-middle income)* | Children |
| 6 | Boehm A.B. | 2016 | Occurrence of Host-Associated Fecal Markers on Child Hands, Household Soil, and Drinking Water in Rural Bangladeshi Households | Bangladesh *(low income)* | Children |
| 7 | Budge S. | 2019 | Do domestic animals contribute to bacterial contamination of infant transmission pathways? Formative evidence from Ethiopia | Ethiopia *(low income)* | Children and caregivers |
| 8 | Byrne D. | 2021 | Navigating Data Uncertainty and Modeling Assumptions in | Uganda *(low income)* | Children and caregivers |

| | | | | | |
|---|---|---|---|---|---|
| | | | Quantitative Microbial Risk Assessment in an Informal Settlement in Kampala, Uganda | | |
| 9 | Carabin H. | 2001 | Comparison of methods to analyse imprecise faecal coliform count data from environmental samples | Canada *(high income)* | Children and educators in daycare |
| 10 | Carabin H. | 1999 | Effectiveness of a training program in reducing infections in toddlers attending day care centers | Canada *(high income)* | Children and educators in daycare |
| 11 | Carrasco L. | 2008 | Occurrence of faecal contamination in households along the US–Mexico border | US, Mexico *(high, upper-middle income)* | Caregivers |
| 12 | Contreras J. | 2021 | Longitudinal Effects of a Sanitation Intervention on Environmental Fecal Contamination in a Cluster-Randomized Controlled Trial in Rural Bangladesh | Bangladesh *(low income)* | Mothers and children |
| 13 | Cranston I.* | 2015 | Transmission of *Enterobius vermicularis* eggs through hands of school children in rural South Africa | South Africa *(upper-middle income)* | Children in school |
| 14 | Curin K. | 1999 | Hygienic conditions in elementary and secondary schools in the county of Split-Dalmatia | Croatia *(lower-middle income)* | Children in school |
| 15 | Davis J. | 2011 | The effects of informational interventions on household water management, hygiene behaviors stored drinking water quality and hand contamination in Peri-Urban Tanzania | Tanzania *(low income)* | Female heads of household |
| 16 | de Wit J.C. | 1992 | Faecal micro-organisms on the hands of carriers: *Escherichia coli* as model salmonella | Netherlands *(high income)* | Students and staff at University |
| 17 | Devamani C. | 2014 | A Simple Microbiological Tool to Evaluate the Effect of Environmental Health Interventions on Hand Contamination | India, Mozambique *(lower-middle, low income)* | Men, mothers of young children, grandmothers |
| 18 | Dodrill L. | 2011 | Male commuters in north and south England: risk factors for the presence of faecal bacteria on hands | Great Britain *(high income)* | Male commuters |

| | | | | | |
|---|---|---|---|---|---|
| 19 | Dutta S. | 1997 | Isolation of *Escherichia coli* to detect faecal contamination of infants and their mothers in west Bengal | India *(low income)* | Infants and mothers |
| 20 | Ekanem E. | 1983 | Transmission dynamics of enteric bacteria in day-care centers | US *(high income)* | Children in daycare |
| 21 | Ercumen A. | 2018 | Effects of single and combined water, sanitation and handwashing interventions on fecal contamination in the domestic environment: a cluster-randomized controlled trial in rural Bangladesh | Bangladesh *(low income)* | Children |
| 22 | Friedrich M.N. | 2017 | Handwashing, but how? Microbial effectiveness of existing handwashing practices in high-density suburbs of Harare, Zimbabwe | Zimbabwe *(low income)* | Caregiver in household |
| 23 | Fuhrmeister E.R. | 2019 | Predictors of enteric pathogens in the domestic environment from human and animal sources in rural Bangladesh | Bangladesh *(low income)* | Children and Mothers in household |
| 24 | George C. | 2021 | Child hand contamination is associated with subsequent pediatric diarrhea in rural Democratic Republic of the Congo (REDUCE Program) | Democratic Republic of Congo *(low income)* | Children and caregivers |
| 25 | Ghuliani A. | 1995 | Contamination of weaning foods and transmission of *E. coli* in causation of infantile diarrhea in low income group in Chandigarh | India *(low income)* | Children (low income) |
| 26 | Gil A.I. | 2014 | Fecal contamination of food, water, hands, and kitchen utensils at the household level in rural areas of Peru | Peru *(lower-middle income)* | Children and caregivers in household |
| 27 | Gizaw Z. | 2022 | Effects of local handwashing agents on microbial contamination of the hands in a rural setting in Northwest Ethiopia: a cluster randomised controlled trial | Ethiopia *(low income)* | Mothers and caregivers of children under 5 |
| 28 | Gorman R. | 2002 | A study of cross-contamination of food-borne pathogens in the | Ireland *(high income)* | Adults in household |

| | | | | | |
|---|---|---|---|---|---|
| | | | domestic kitchen in the Republic of Ireland | | |
| 29 | Greene L.E. | 2012 | Impact of a school-based hygiene promotion and sanitation intervention on pupil hand contamination in western Kenya: a cluster randomized trial | Kenya *(low income)* | Children in school |
| 30 | Grimason A.M. | 2014 | Knowledge, awareness and practice of the importance of hand-washing amongst children attending state run primary schools in rural Malawi | Malawi *(low income)* | Children in school |
| 31 | Harris A.R. | 2016 | Ruminants contribute fecal contamination to the urban household environment in Dhaka, Bangladesh | Bangladesh *(low income)* | Children in household |
| 32 | Harris A.R. | 2019 | Comparison of analytical techniques to explain variability in stored drinking water quality and microbial hand contamination of female caregivers in Tanzania | Tanzania *(low income)* | Female caregivers |
| 33 | Hartinger S.M. | 2016 | Improving household air, drinking water in Peru | Peru *(lower-middle income)* | Mothers |
| 34 | Hartinger S.M. | 2011 | A community randomised controlled trial evaluating a home-based environmental intervention package of improved stoves, solar water disinfection and kitchen sinks in rural Peru: Rationale, trial design and baseline findings | Peru *(lower-middle income)* | Mothers |
| 35 | Holaday B. | 1990 | Patterns of fecal coliform contamination in day-care centers | US *(high income)* | Children and staff in daycare |
| 36 | Holaday B. | 1995 | Diaper type and fecal contamination in child day care | US *(high income)* | Children and staff at daycare |
| 37 | Huda T.M.N. | 2019 | Effect of neighborhood sanitation coverage on fecal contamination of the household environment in rural Bangladesh | Bangladesh *(low income)* | Children under 2 yrs |
| 38 | Islam M.S. | 2016 | Faecal contamination of commuters’ hands in main vehicle stations in Dhaka city, Bangladesh | Bangladesh *(low income)* | Adult commuters |

| | | | | | |
|---|---|---|---|---|---|
| 39 | Judah G. | 2010 | Dirty hands bacteria of faecal origin on commuters' hands | Great Britain *(high income)* | Adult commuters |
| 40 | Julian T.R. | 2015 | A pilot study on integrating videography and environmental microbial sampling to model fecal bacterial exposures in peri-urban Tanzania | Tanzania *(low income)* | Female adults |
| 41 | Julian T.R. | 2013 | Enterococcus spp on fomites and hands indicate increased risk of respiratory illness in childcare centers | US *(high income)* | Children and staff at daycare |
| 42 | Kaltenthaler E.C. | 1996 | The use of microbiology in the study of hygiene behaviour | Botswana *(lower-middle income)* | Children under 6 yrs and caregivers |
| 43 | Kaltenthaler E.C. | 1995 | Faecal contamination on children's hands and environmental surfaces in primary schools in Leeds | Great Britain *(high income)* | Children in school |
| 44 | Kalter H.D. | 2010 | Risk factors for antibiotic-resistant *Escherichia coli* carriage in young children in Peru: Community-based cross-sectional prevalence study | Peru *(lower-middle income)* | Mothers |
| 45 | Kavitha E. | 2019 | Bacteriological profile and perception on hand hygiene in school going children | India *(lower-middle income)* | Children in school |
| 46 | Kellogg D.S. | 2012 | High fecal hand contamination among wilderness hikers | US *(high income)* | Hikers |
| 47 | Keswick B.H. | 1983 | Survival and detection of rotaviruses on environmental surfaces in day care centers | US *(high income)* | Teacher in childcare center |
| 48 | Kim S. | 2019 | Evaluation of the impact of antimicrobial hand towels on hand contamination with E. coli among mothers in Kisumu County Kenya, 2011-2012 | Kenya *(low income)* | Mothers with child under 5 |
| 49 | Kundu A. | 2018 | Drinking water safety role of hand hygiene sanitation facility and water system in semi-urban areas of India | India *(lower-middle income)* | Mothers and children under 5 yrs |
| 50 | Kusuma A. | 2012 | *Escherichia coli* contamination of babies' food-serving utensils in a district of West Sumatra, Indonesia | Indonesia *(lower-middle income)* | People who feed babies |

| | | | | | |
|---|---|---|---|---|---|
| 51 | Kyriacou A. | 2009 | Screening for faecal contamination in primary schools in Crete, Greece | Greece *(high income)* | Children in school |
| 52 | Laborde D.J. | 1993 | Effect of fecal contamination on diarrheal illness rates in day-care centers | US *(high income)* | Children and staff at day cares |
| 53 | Li Y. | 2014 | Microbiological analysis of environmental samples collected from childcare facilities in North and South Carolina | US *(high income)* | Staff at day cares |
| 54 | Luby S.P. | 2007 | Field trial of a low cost method to evaluate hand cleanliness | Pakistan *(low income)* | Mothers of children under 5 yrs |
| 55 | Makata K. | 2021 | Hand hygiene intervention to optimise soil-transmitted helminth infection control among primary school children: the Mikono Safi cluster randomised controlled trial in northwestern Tanzania | Tanzania *(low income)* | Children (school age) |
| 56 | Martinez-Bastidas T. | 2014 | Detection of pathogenic micro-organisms on children's hands and toys during play | Mexico *(upper-middle income)* | Children |
| 57 | Mason M. | 2014 | Prevalence, Characteristics, and Epidemiology of Microbial Hand Contamination Among Minnesota State Fair Attendees | United States *(high income)* | State fair attendees (adults and children) |
| 58 | Mattioli M. | 2015 | Quantification of human norovirus GII on hands of mothers with children under the age of five years in Bagamoyo, Tanzania | Tanzania *(low income)* | Female care givers |
| 59 | Mattioli M. | 2013 | Hands and water as vectors of diarrheal pathogens in Bagamoyo, Tanzania | Tanzania *(low income)* | Female head of household with child under 5 yrs |
| 60 | Mattioli M. | 2014 | Enteric pathogens in stored drinking water and on caregiver's hands in Tanzanian households with and without reported cases of child diarrhea | Tanzania *(low income)* | Adult female caregiver |
| 61 | Monira S. | 2022 | Fecal Sampling of Soil, Food, Hand, and Surface Samples from Households in Urban Slums of Dhaka, Bangladesh: An Evidence-Based Development of Baby | Bangladesh *(low income)* | Children and caregivers |

| | | | | | |
|---|---|---|---|---|---|
| | | | Water, Sanitation, and Hygiene Interventions | | |
| 62 | Navab-Daneshman T. | 2018 | *Escherichia coli* contamination across multiple environmental compartments (soil, hands, drinking water, and handwashing water) in urban Harare: Correlations and risk factors | Zimbabwe *(low income)* | Caregiver |
| 63 | Ngure F.M. | 2013 | Formative research on hygiene behaviors and geophagy among infants and young children and implications of exposure to fecal bacteria.pdf | Zimbabwe *(low income)* | Children and caregivers |
| 64 | Odagiri M. | 2016 | Human fecal and pathogen exposure pathways in rural Indian villages and the effect of increased latrine coverage | India *(lower-middle income)* | Mothers and youngest child under 5 yrs |
| 65 | Oristo S. | 2017 | Contamination by Norovirus and Adenovirus on Environmental Surfaces and in Hands of Conscripts in Two Finnish Garrisons | Finland *(high income)* | Soldiers |
| 66 | Otsuka Y. | 2019 | Comprehensive assessment of handwashing and faecal contamination among elementary school children in an urban slum of Indonesia | Indonesia *(lower-middle income)* | Children in school |
| 67 | Padaruth S.K. | 2015 | Hygiene practices and faecal contamination of the hands of children attending primary school in Mauritius | Mauritius *(upper-middle income)* | Children in school |
| 68 | Petersen N.J. | 1986 | Design and modification of the day care environment | US *(high income)* | Children in daycare |
| 69 | Pickering A. | 2018 | Fecal indicator bacteria along multiple environmental transmission pathways (waters, hands, food, soil, flies) and subsequent child diarrhea in rural Bangladesh | Bangladesh *(low income)* | Children (youngest in household) |
| 70 | Pickering A.J. | 2010 | Hands, water, and health: Fecal contamination in Tanzanian communities with improved, non-networked water supplies | Tanzania *(low income)* | Children and mothers in household |

| | | | | | |
|---|---|---|---|---|---|
| 71 | Pickering A.J. | 2010 | Efficacy of Waterless Hand Hygiene Compared with Handwashing with Soap: A Field Study in Dar es Salaam, Tanzania | Tanzania *(low income)* | Students and adults in school |
| 72 | Pickering A.J. | 2011 | Bacterial hand contamination among Tanzanian mothers varies temporally and following household activities | Tanzania *(low income)* | Female care takers in household with at least one child under 5 yrs |
| 73 | Pinfold J.V. | 1990 | Faecal contamination of water and fingertip-rinses as a method for evaluating the effect of low-cost water supply and sanitation activities on faeco-oral disease transmission. I. A case study in rural north-east Thailand | Thailand *(lower-middle income)* | Female head of household |
| 74 | Ram P.K. | 2011 | Variability in hand contamination based on serial measurements: Implications for assessment of hand-cleansing behavior and disease risk | Bangladesh *(low income)* | Mothers of children under 2 yrs |
| 75 | Reddi Sgdnl | 2016 | Bacteriological quality of drinking water at point of use and hand hygiene of primary food preparers implications for household food safety | India *(lower-middle income)* | Primary food preparers in household |
| 76 | Rothstein J.D. | 2019 | Household contamination of baby bottles and opportunities to improve bottle hygiene in Peri Urban Lima Peru | Peru *(upper-middle income)* | Caregivers |
| 77 | Saboori S. | 2013 | Impact of regular soap provision to primary schools on hand washing and E. coli hand contamination among pupils in Nyanza province, Kenya: A cluster-randomized trial | Kenya *(low income)* | Children in school |
| 78 | Schriewer A. | 2015 | Human and Animal Fecal Contamination of Community Water Sources, Stored Drinking Water and Hands in Rural India Measured with Validated Microbial Source Tracking Assays | India *(lower-middle income)* | Mothers and youngest child under 5 yrs |
| 79 | Steinbaum L.* | 2017 | Following the Worms: Detection of Soil-Transmitted Helminth | Kenya | Mothers |

| | | | | | |
|---|---|---|---|---|---|
| | | | Eggs on Mothers' Hands and Household Produce in Rural Kenya | *(lower-middle income)* | |
| 80 | Ubheeram J. | 2017 | Effectiveness of hand hygiene education among a random sample of women from the community | Mauritius *(upper-middle income)* | Female volunteers in the community |
| 81 | Van R. | 1991 | Environmental contamination in child day-care centers | US *(high income)* | Caregivers and children under 2 yrs |
| 82 | Van R. | 1991 | The effect of diaper type and overclothing on fecal contamination in day-care centers | US *(high income)* | Children and caregivers in daycare |
| 83 | Vishwanath R. | 2019 | Detection of bacterial pathogens in the hands of rural school children across different age groups and emphasizing the importance of hand wash | India *(lower-middle income)* | Children in school |
| 84 | Winter J. | 2021 | The impact of on-premises piped water supply on fecal contamination pathways in rural Zambia | Zambia *(low income)* | Caregivers |

*Additional papers that were not found using search criteria, but were known by authors
